# Infection and transmission risks in schools and contribution to the COVID-19 pandemic in Germany – a retrospective observational study using nation-wide and regional health and education agency notification data

**DOI:** 10.1101/2022.01.18.22269200

**Authors:** Torben Heinsohn, Berit Lange, Patrizio Vanella, Isti Rodiah, Stephan Glöckner, Alexander Joachim, Dennis Becker, Tobias Brändle, Stefan Dhein, Stefan Ehehalt, Mira Fries, Annette Galante-Gottschalk, Stefanie Jehnichen, Sarah Kolkmann, Annelene Kossow, Martin Hellmich, Jörg Dötsch, Gérard Krause

## Abstract

**Introduction:** Currently, information on infection and transmission risks of students and teachers in schools, the effect of infection control measures for schools as well as the contribution of schools to the overall population transmission of SARS-CoV-2 in Germany are limited to regional data sets restricted to short phases of the pandemic.

**Methods:** We used German federal state (NUTS-2) and county (NUTS-3) data from national and regional public health and education agencies to assess infection risk and secondary attack rates (SARs) from March 2020 to October 2021 in Germany. We used multiple regression analysis and infection dynamic modelling, accounting for urbanity, socioeconomic factors, local population infection dynamics and age-specific underdetection to investigate the effects of infection control measures.

**Results:** We included (1) nation-wide NUTS-2 level data from calendar weeks (W) 46-50/2020 and W08-40/2021 with 304676 infections in students and 32992 in teachers; (2) NUTS-3 level data from W09-25/2021 with 85788 student and 9427 teacher infections and (3) detailed data from 5 regions covering W09/2020 to W27/2021 with 12814 infections, 43238 contacts and 4165 secondary cases for students (for teachers 14801, 5893 and 472 respectively).

In counties with mandatory surgical mask wearing during class in all schools infection risk of students and teachers was reduced by 56/100.000 persons per 14 days and by 30% and 24% relative to the population respectively. Overall contribution to population infections of contacts in school settings was 2-13%. It was lowest during school closures and vacation and highest during normal presence class intervals. Infection risk for students increased with age and was similar to or lower than the population risk during second and third waves in Germany and higher in summer 2021. Infection risk of teachers was higher than the population during the second wave and similar or lower thereafter with stricter measures in place. SARs for students and staff were below 5% in schools throughout the study period. SARs in households more than doubled from 14% W21-39/2020 to 29-33% in W08-23/2021. Most contacts were reported for schools, yet most secondary cases originated in households. In schools, staff predominantly infected staff and students predominantly infected students.

**Conclusion:** Open schools under hygiene measures and testing strategies contribute up to 13% of nation-wide infections in Germany and as little as 2% during vacations/school closures. Tighter infection control measures stabilised school SARs whilst household SARs more than doubled as more transmissible variants became prevalent in Germany. Mandatory mask wearing during class in all school types effectively reduces secondary transmission in schools, as do reduced attendance class models.

## INTRODUCTION

Schools have been a key target domain for nonpharmaceutical interventions (NPIs) in the SARS-CoV-2 pandemic. However, infection dynamics in schools, their impact on the wider population and the effect of NPIs remain ill-defined. Measures include isolation and quarantine, masking and testing, hygiene measures, ventilation and social distancing, including reduced attendance class models and school closures(1-3).

In Germany, the 16 federal states hold sovereignty over education policy and in large parts health system management. During the SARS-CoV-2 pandemic in Germany, schools were first closed on March 17, 2020, nationally and gradually reopened in May 2020. Subsequently, federal states instituted diverse NPIs around social distancing, cloth masking and reduced class sizes. Full school closures were implemented again nationally on December 16, 2020, until reopening under federal state-specific NPIs in February/March 2021. In February 2021 the German Association of Scientific Medical Societies published an evidence-based guideline on NPIs to reduce transmission of SARS-CoV-2 in schools(4). The variability of measures between federal states diminished but remained in some parts such as masking of primary school students or masking of students during class. Different testing strategies (e.g. antigen testing, pooled PCR testing, mandatory/voluntary) for students and teachers were implemented in spring 2021. For a third time school closures, now depending on local infection dynamics, were instituted in April 2021 along with mandatory testing in schools nationally. All federal states returned to face-to-face classes and reduced stringency of measures prior to the summer vacation 2021. In September 2021 the adult population in Germany surpassed a vaccination coverage of 60%(5). The National Advisory Committee for Immunisations (Ständige Impfkommission, STIKO) issued a vaccine recommendation for adolescents from 12-17 years of age and for children at risk in June 2021 and a general recommendation for all adolescents in August 2021(6).

Whereas school closures have proven effective in containing SARS-CoV-2 transmission(7-9), they are associated with adverse effects on the physical and mental health of students and exacerbate ethnic and socioeconomic disparities(10-14). Legislators implemented NPIs and testing in schools to reduce setting specific transmission(15) and avoid closures. Evidence of effects of measures was initially derived from modelling studies or indirectly from non-school settings(3). The sovereignty of federal states over education policy resulted in 16 different and parallel approaches. This allows for comparative evaluation of infection dynamics and the effects of NPIs in schools.

The risk of infection and transmission in schools relative to the population remain contentious. In a systematic review(16) we showed that during phases with low population incidence, there is no increased risk of infection in students or school staff, however with increasing infection dynamics in the population, the risk of infection in students and staff increases. In Germany and Europe, studies on school transmission risk and contribution to overall transmission in the population are based on geographically and temporally limited data (3, 17-20), with few exceptions(21).

In Germany, data on infection dynamics is collected at the county level by over 400 public health agencies and reported to the national Robert Koch Institute (RKI). Detailed data on (school) setting-specific infection dynamics, contacts, and their risk of infection are not reported to the RKI and can only be obtained individually from the 400+ public health and education agencies. Thus, studies investigating school setting transmission in Germany are limited to small geographic units and short time periods.

In addition, most current studies remain descriptive and do not account for age- and setting-specific infection dynamics and underdetection of cases(18-20, 22). Both, setting specific transmission risk as well as setting specific contribution to overall infections is crucial to inform pandemic policy on effects of NPIs for schools.

In this study we obtained federal state and county level data for school settings as well as detailed regional data obtained from individual public health and educational agencies to assess infection risk, secondary attack rates, the contribution of schools to the overall transmission as well as the effect of NPIs in schools during the first 18 months of the pandemic in Germany.

## METHODOLOGY

### Study design and data sources

First, we performed a retrospective observational study of prospectively collected SARS-CoV-2 infection case report data from health and educational authorities in Germany (Ethical approval N°9609_BO_K_2021, Hannover) from 5 regions (S1 Table, S2 Table, S3 Text). Public health authorities report case data according to the Infection Protection Act (IfSG). Cases were defined by direct viral detection via nasopharyngeal swabs using PCR or by cultural isolation of the pathogen. In addition to notified cases, some regional data included secondary cases as well as contacts per case, allowing to compute secondary attack rates (S1 Table).

Secondly, we used data collected by the Standing Conference of Ministers of Education and Cultural Affairs in Germany (KMK) (23). Federal state agencies collected data weekly in a structured format from all schools within their jurisdiction and included the number of students, classes and staff infected or absent due to quarantine and school closures. Federal state-level data is publicly available from calendar week (W) 46/2020 onwards, excluding school closures. In addition, we received unpublished county-level data, collected in the same manner on the same parameters (S1 Table)(24).

Thirdly, we sourced population infection and incidence data from the Robert Koch Institute (RKI) SURVSTAT tool, described as incidence per 100000 inhabitants(25). We also sourced time and county-specific data on hygiene and testing measures in school from government sources. Further information on data sources and collection is in the supplement (S1-S3).

### Data analysis

We performed a descriptive analysis of infection risk for students and teachers as cumulative risks and crude risk ratios for specified periods of time with 95% Confidence Intervals (CI). We calculated secondary attack rates (SAR) as percentages with 95% CI of those being identified as notified infections in all known contacts. We adapted the pandemic phases as proposed by the RKI(26) for school policy intervals (Table 1). An overview of population incidences across phases is in the supplement (S4 Figure).

**Table 1.** Overview of the Pandemic Phases and Outline of School Policies

| Phase | Duration | Infection environment | School policy environment |
| --- | --- | --- | --- |
| 1 | W10-W20/2020 | First Covid-19 wave in Germany | - W12-17 school closures<br>- W18+ phased reopening with hygiene concepts |
| 2 | W21-39/2020 | Summer Plateau 2020 | - W21+ phased reopening with hygiene concepts<br>- Summer vacation<br>- Normal face-to-face class with sporadic cloth mask rules introduced |
| 3 | W40/2020 – 08/2021 | Second Covid-19 wave in Germany | - W40-51 face-to-face classes with tightening cloth mask rules<br>- W52-02 Christmas vacation<br>- W02-08 school closures |
| 4 | W09-23/2021 | Third Covid-19 wave in Germany | - W09+ reduced attendance class models with surgical mask rules, gradual introduction of testing, teachers given priority for vaccinations<br>- W16+ *Bundesnotbremse* with incidence-dependent school closures, mandatory testing<br>- W21+ phased return to face-to-face classes |
| 5 | W24/2021+ | Summer Plateau 2021 | - Face-to-face class with some lifting of mask and testing mandates<br>- Summer vacation<br>- W30 vaccination coverage in the total population reaches 50%<br>- W33 vaccinations started for adolescents age 12 to 17 years |
| **Adaptations made for this study** |  |  |  |
| 3a | W40-51/2020 | Second Covid-19 wave in Germany | - W40-51 face-to-face classes with tightening cloth mask rules |
| 3b | W52/2020 – W07/2021 | Second Covid-19 wave in Germany | - W52-02 Christmas vacation<br>- W02-08 school closures |
| Phase 3/4 transition | W07-08/2021 |  | - School reopening from W08/2021 |

Based on Schilling et al, 2021 (26), own illustration and additions.

We investigated infection control measures and other factors influencing infection risk of teachers or students in a multiple linear regression model. We used an ordinary least squares (OLS) model to test for statistical associations of the official active cases per 100000 students and teachers as reported by the KMK for the county level on the two-week-lags of the attendance rates (by educational level), the stringency of mask mandates (Table 1 in S3 Text) and the mandate and stringency of testing in schools. We controlled for the two-week incidence per 100000 inhabitants of the corresponding county, the percentage of fully vaccinated persons in the corresponding federal state, the socioeconomic status and geography of the county (S3 Text, S11 Text). Furthermore, we tested whether the school specific NPIs had an over proportional effect on the school population compared to the overall population. We proceeded similarly to the first approach, yet did not include a control group as an explanatory variable but instead divided the active cases among the students or teachers, respectively, by the 14-day incidence of the total population of the district. This way we estimated the effect of the NPIs on cases in schools relative to cases in the population, thus estimating whether the NPIs in schools have a significantly different effect on the overall population as on the school population. Further information on the regression analyses are in the supplement (S5 Tables, S3 Text).

The infection dynamics in schools were estimated using a SEIRS (Susceptible-Exposed-Infectious-Recovered-Susceptible) model previously described(27). This distinguishes between healthy but susceptible individuals, those infected but not yet infectious (exposed), and symptomatic and asymptomatic patients. In addition, we included compartments for hospitalisations, patients entering intensive care units (ICUs) and persons with long-COVID, i.e. those who continue to have sequelae after recovery. In the final state, the patients are recovered or dead. Furthermore, we assume a reinfection process. We split the recovery compartment into a compartment for those recovered from COVID and a long-COVID compartment, since we assume that both have a different reinfection rate. Fig.1 illustrates the model as a flow chart.

**Figure 1.**
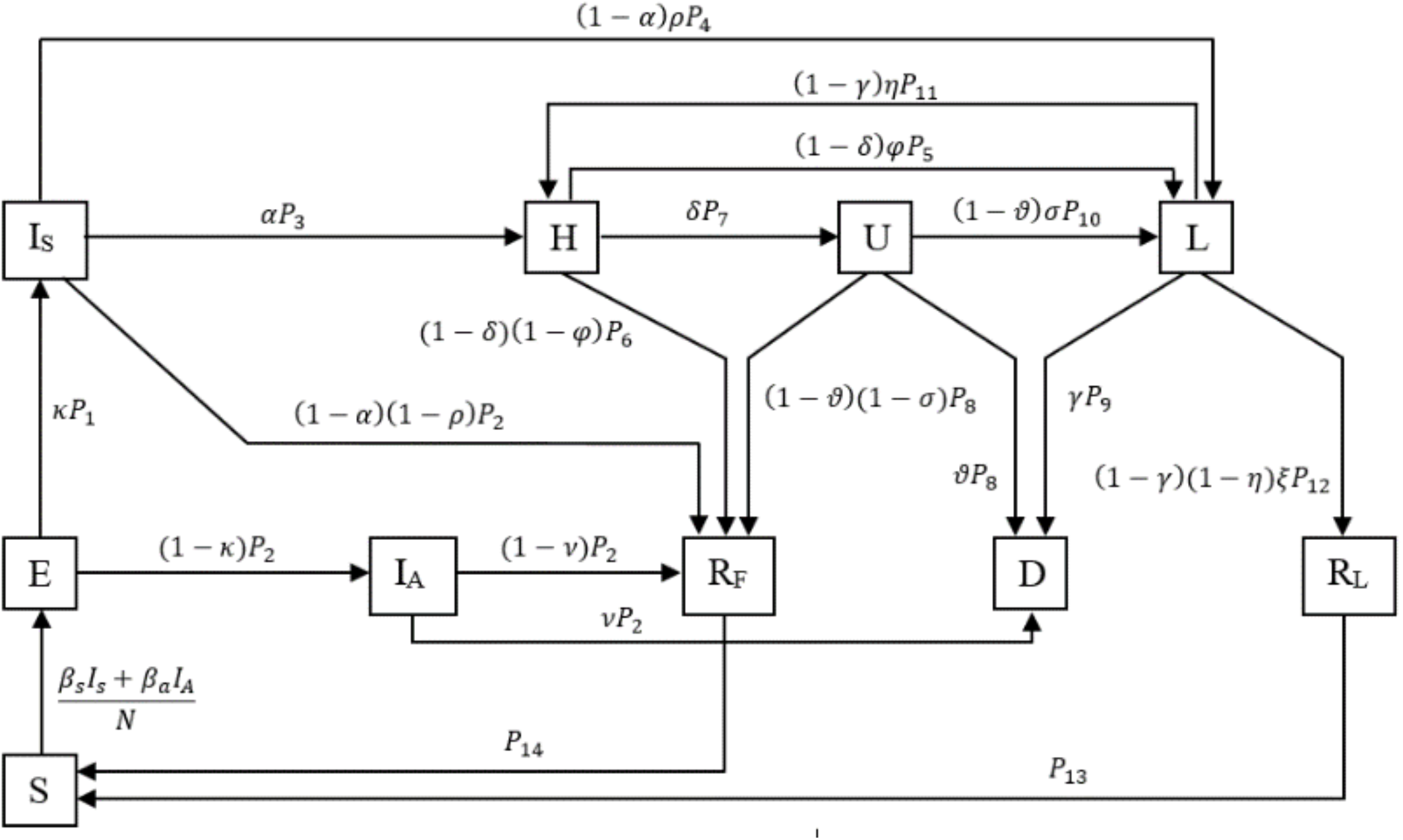
Compartmental SEIR model. An individual in age group *i* is classified either as susceptible (S), exposed (E), asymptomatically infected (IA), symptomatically infected (IS), hospitalized (H), in intensive care (U), suffering under long-COVID (L), fully recovered (RF), recovered from long-COVID (R_L_), and dead (D). *P*_1_ − *P*_8_ are health state transition rates. The Greek letters quantify transition probabilities between the statuses.

The remaining parameters represent time periods of transitions between the different statuses as well as probabilities for the transitions and are estimated from international data and literature research. The data for the model is composed of reports from KMK(23), RKI (25) and the DIVI Intensive Care Register(28). We accounted for age-specific underdetection taking pandemic period- and age-specific underdetection ratios from a large seroprevalence study in Germany(29).

## RESULTS

### Risk of infection for students and teachers in Germany according to nation-wide data

Data obtained from KMK was collected on the federal state level for W46-50/2020 and W08-40/2021 (at time of analysis), covering 304676 student and 32992 teacher cases. Cumulative risk of notified SARS-CoV-2 infection for students was 0.96% W46-50/2020, 1.22% in phase 4, 1.16% in phase 5 and for teachers 1.77%, 1.53% and 0.47% respectively. At the county level in W09-25, infection risk was 1.68% for students and 1.51% for teachers. Quarantine risk of students and teachers over federal states varied widely from 1-21% during all phases of the pandemic (Table 2, Table 1 in S7 Tables).

**Table 2.**
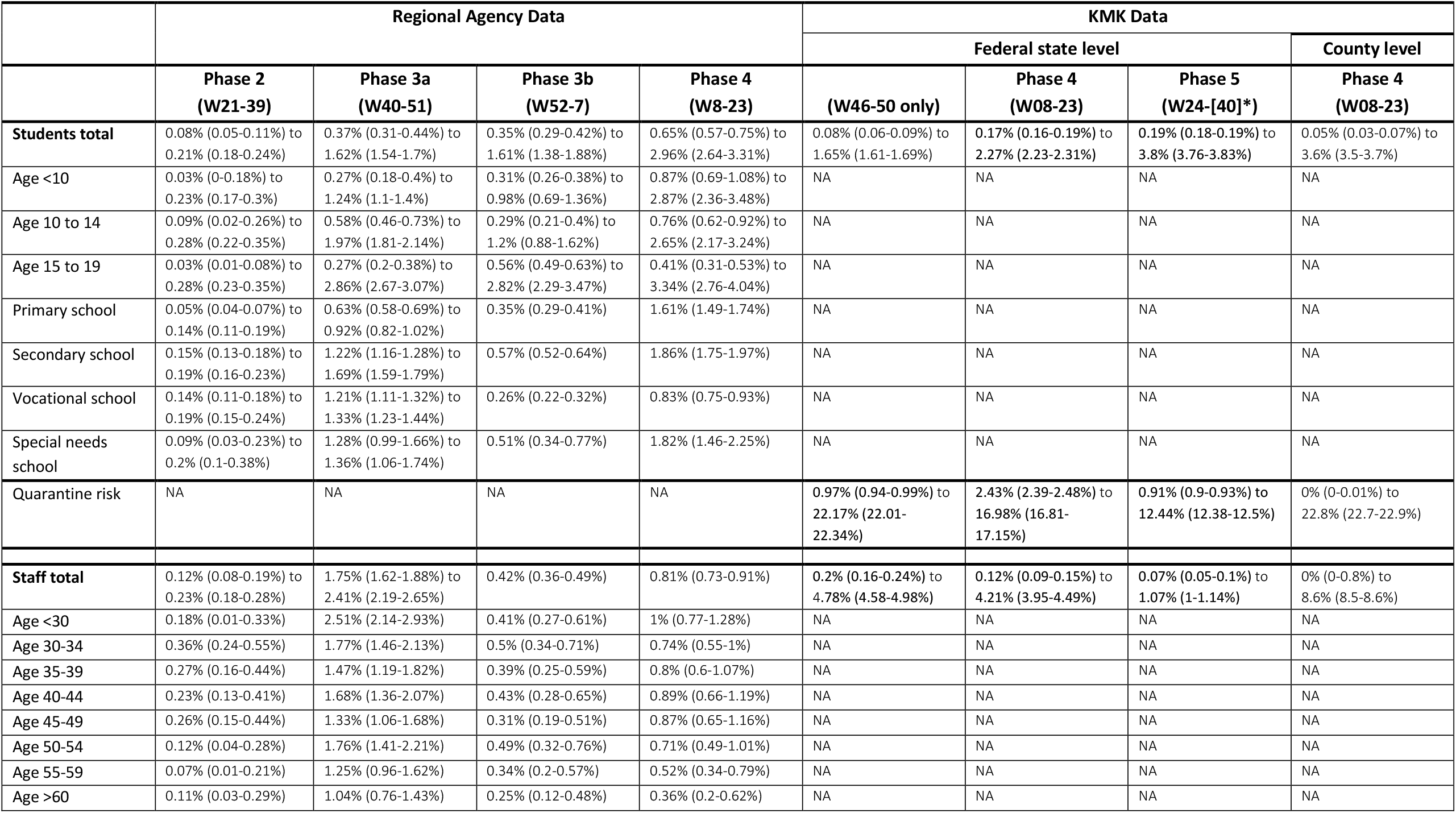
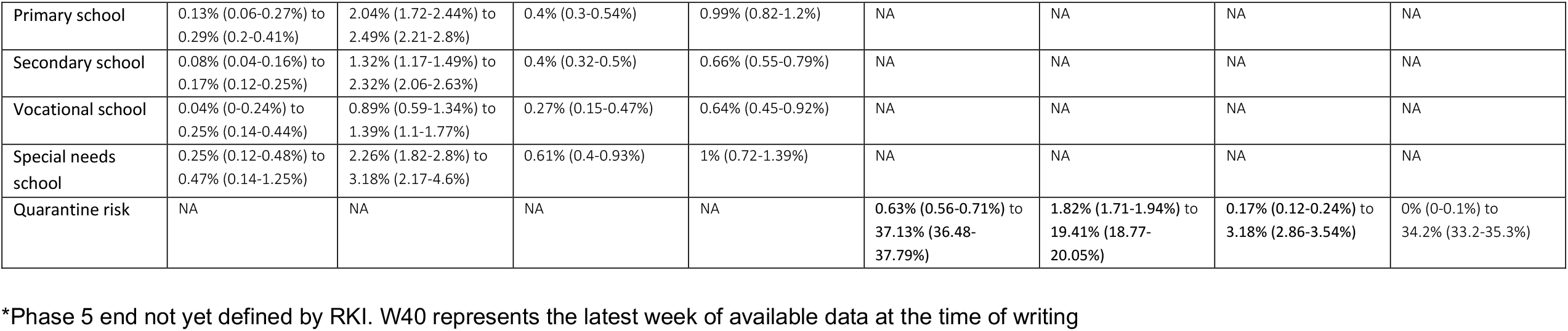
Risk of infection over pandemic phases with mimima and maxima risk per federal state/ county

|  | Regional Agency Data |  |  |  | KMK Data |  |  |  |
| --- | --- | --- | --- | --- | --- | --- | --- | --- |
|  |  |  |  |  | Federal state level |  |  | County level |
|  | Phase 2<br>(W21-39) | Phase 3a<br>(W40-51) | Phase 3b<br>(W52-7) | Phase 4<br>(W8-23) | (W46-50 only) | Phase 4<br>(W08-23) | Phase 5<br>(W24-[40]*) | Phase 4<br>(W08-23) |
| **Students total** | 0.08% (0.05-0.11%) to<br>0.21% (0.18-0.24%) | 0.37% (0.31-0.44%) to<br>1.62% (1.54-1.7%) | 0.35% (0.29-0.42%) to<br>1.61% (1.38-1.88%) | 0.65% (0.57-0.75%) to<br>2.96% (2.64-3.31%) | 0.08% (0.06-0.09%) to<br>1.65% (1.61-1.69%) | 0.17% (0.16-0.19%) to<br>2.27% (2.23-2.31%) | 0.19% (0.18-0.19%) to<br>3.8% (3.76-3.83%) | 0.05% (0.03-0.07%) to<br>3.6% (3.5-3.7%) |
| Age <10 | 0.03% (0-0.18%) to<br>0.23% (0.17-0.3%) | 0.27% (0.18-0.4%) to<br>1.24% (1.1-1.4%) | 0.31% (0.26-0.38%) to<br>0.98% (0.69-1.36%) | 0.87% (0.69-1.08%) to<br>2.87% (2.36-3.48%) | NA | NA | NA | NA |
| Age 10 to 14 | 0.09% (0.02-0.26%) to<br>0.28% (0.22-0.35%) | 0.58% (0.46-0.73%) to<br>1.97% (1.81-2.14%) | 0.29% (0.21-0.4%) to<br>1.2% (0.88-1.62%) | 0.76% (0.62-0.92%) to<br>2.65% (2.17-3.24%) | NA | NA | NA | NA |
| Age 15 to 19 | 0.03% (0.01-0.08%) to<br>0.28% (0.23-0.35%) | 0.27% (0.2-0.38%) to<br>2.86% (2.67-3.07%) | 0.56% (0.49-0.63%) to<br>2.82% (2.29-3.47%) | 0.41% (0.31-0.53%) to<br>3.34% (2.76-4.04%) | NA | NA | NA | NA |
| Primary school | 0.05% (0.04-0.07%) to<br>0.14% (0.11-0.19%) | 0.63% (0.58-0.69%) to<br>0.92% (0.82-1.02%) | 0.35% (0.29-0.41%) | 1.61% (1.49-1.74%) | NA | NA | NA | NA |
| Secondary school | 0.15% (0.13-0.18%) to<br>0.19% (0.16-0.23%) | 1.22% (1.16-1.28%) to<br>1.69% (1.59-1.79%) | 0.57% (0.52-0.64%) | 1.86% (1.75-1.97%) | NA | NA | NA | NA |
| Vocational school | 0.14% (0.11-0.18%) to<br>0.19% (0.15-0.24%) | 1.21% (1.11-1.32%) to<br>1.33% (1.23-1.44%) | 0.26% (0.22-0.32%) | 0.83% (0.75-0.93%) | NA | NA | NA | NA |
| Special needs<br>school | 0.09% (0.03-0.23%) to<br>0.2% (0.1-0.38%) | 1.28% (0.99-1.66%) to<br>1.36% (1.06-1.74%) | 0.51% (0.34-0.77%) | 1.82% (1.46-2.25%) | NA | NA | NA | NA |
| Quarantine risk | NA | NA | NA | NA | 0.97% (0.94-0.99%) to<br>22.17% (22.01-22.34%) | 2.43% (2.39-2.48%) to<br>16.98% (16.81-17.15%) | 0.91% (0.9-0.93%) to<br>12.44% (12.38-12.5%) | 0% (0-0.01%) to<br>22.8% (22.7-22.9%) |
| **Staff total** | 0.12% (0.08-0.19%) to<br>0.23% (0.18-0.28%) | 1.75% (1.62-1.88%) to<br>2.41% (2.19-2.65%) | 0.42% (0.36-0.49%) | 0.81% (0.73-0.91%) | 0.2% (0.16-0.24%) to<br>4.78% (4.58-4.98%) | 0.12% (0.09-0.15%) to<br>4.21% (3.95-4.49%) | 0.07% (0.05-0.1%) to<br>1.07% (1-1.14%) | 0% (0-0.8%) to<br>8.6% (8.5-8.6%) |
| Age <30 | 0.18% (0.01-0.33%) | 2.51% (2.14-2.93%) | 0.41% (0.27-0.61%) | 1% (0.77-1.28%) | NA | NA | NA | NA |
| Age 30-34 | 0.36% (0.24-0.55%) | 1.77% (1.46-2.13%) | 0.5% (0.34-0.71%) | 0.74% (0.55-1%) | NA | NA | NA | NA |
| Age 35-39 | 0.27% (0.16-0.44%) | 1.47% (1.19-1.82%) | 0.39% (0.25-0.59%) | 0.8% (0.6-1.07%) | NA | NA | NA | NA |
| Age 40-44 | 0.23% (0.13-0.41%) | 1.68% (1.36-2.07%) | 0.43% (0.28-0.65%) | 0.89% (0.66-1.19%) | NA | NA | NA | NA |
| Age 45-49 | 0.26% (0.15-0.44%) | 1.33% (1.06-1.68%) | 0.31% (0.19-0.51%) | 0.87% (0.65-1.16%) | NA | NA | NA | NA |
| Age 50-54 | 0.12% (0.04-0.28%) | 1.76% (1.41-2.21%) | 0.49% (0.32-0.76%) | 0.71% (0.49-1.01%) | NA | NA | NA | NA |
| Age 55-59 | 0.07% (0.01-0.21%) | 1.25% (0.96-1.62%) | 0.34% (0.2-0.57%) | 0.52% (0.34-0.79%) | NA | NA | NA | NA |
| Age >60 | 0.11% (0.03-0.29%) | 1.04% (0.76-1.43%) | 0.25% (0.12-0.48%) | 0.36% (0.2-0.62%) | NA | NA | NA | NA |
| Primary school | 0.13% (0.06-0.27%) to 0.29% (0.2-0.41%) | 2.04% (1.72-2.44%) to 2.49% (2.21-2.8%) | 0.4% (0.3-0.54%) | 0.99% (0.82-1.2%) | NA | NA | NA | NA |
| Secondary school | 0.08% (0.04-0.16%) to 0.17% (0.12-0.25%) | 1.32% (1.17-1.49%) to 2.32% (2.06-2.63%) | 0.4% (0.32-0.5%) | 0.66% (0.55-0.79%) | NA | NA | NA | NA |
| Vocational school | 0.04% (0-0.24%) to 0.25% (0.14-0.44%) | 0.89% (0.59-1.34%) to 1.39% (1.1-1.77%) | 0.27% (0.15-0.47%) | 0.64% (0.45-0.92%) | NA | NA | NA | NA |
| Special needs school | 0.25% (0.12-0.48%) to 0.47% (0.14-1.25%) | 2.26% (1.82-2.8%) to 3.18% (2.17-4.6%) | 0.61% (0.4-0.93%) | 1% (0.72-1.39%) | NA | NA | NA | NA |
| Quarantine risk | NA | NA | NA | NA | 0.63% (0.56-0.71%) to 37.13% (36.48-37.79%) | 1.82% (1.71-1.94%) to 19.41% (18.77-20.05%) | 0.17% (0.12-0.24%) to 3.18% (2.86-3.54%) | 0% (0-0.1%) to 34.2% (33.2-35.3%) |

\*Phase 5 end not yet defined by RKI. W40 represents the latest week of available data at the time of writing

On the federal state level, the crude infection risk ratio for students to the population showed an infection risk similar or lower to the general population in W46-50/2020. It was lower in phase 4 and highest in phase 5, where it exceeded that of the general population (Fig.2a). For teachers, the crude risk ratio is highest in W46-50 2020. It is lower in phase 4 and is similar or lower in phase 5 (Fig.2b). The crude infection risk ratio relative to the population is higher for teachers than students in W46-50/2020 and higher for students than teachers in phase 5.

**Figure 2.**
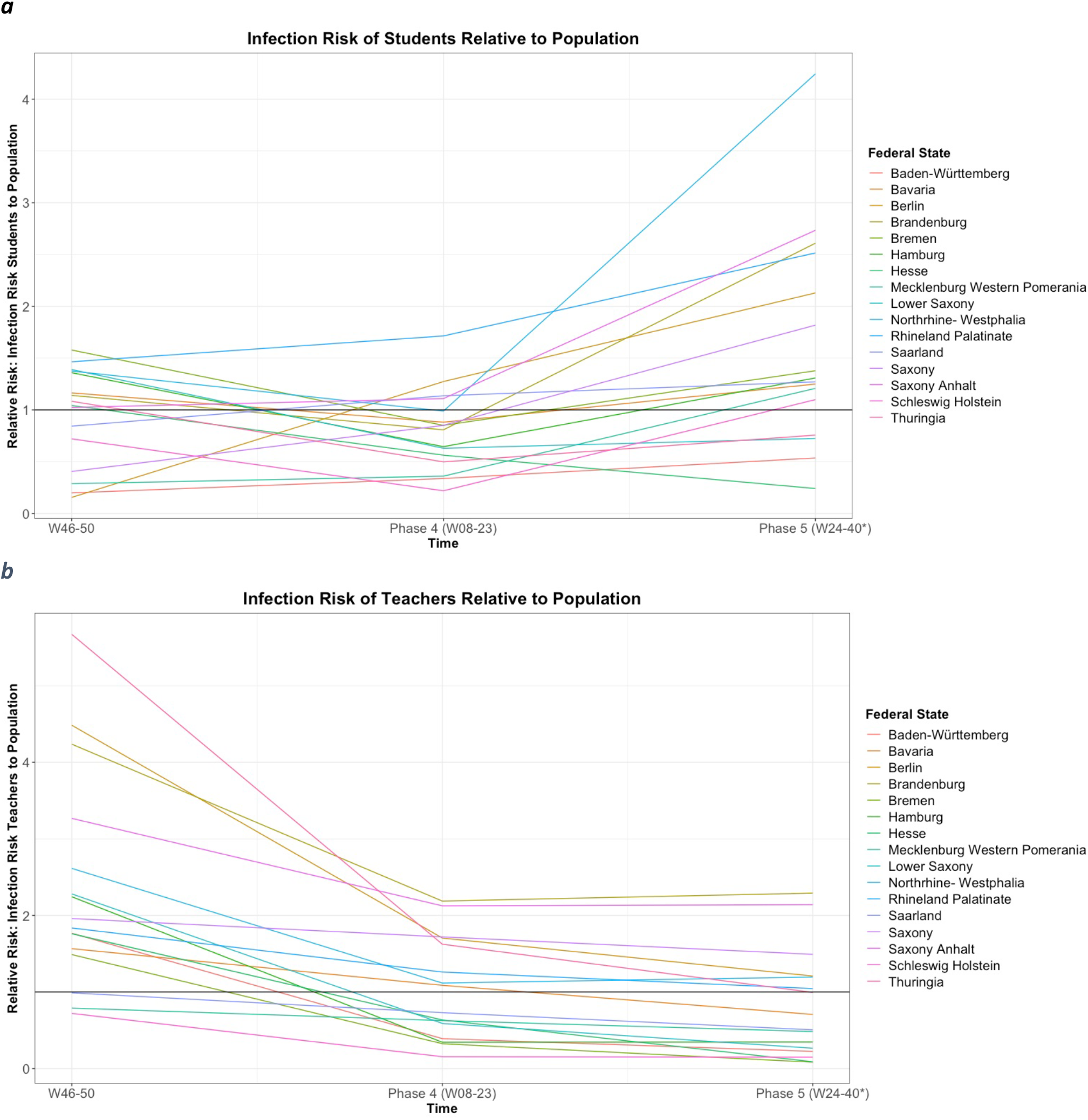
Crude risk ratio of (a) Students to Population and (b) Teachers to Population. Crude risk ratio of infection of students (a) and teachers (b) compared to the infection risk of the population in the respective federal state and time interval. A crude risk ratio of 1 indicates equal risk of infection.

### Risk of infection according to regional data

Data obtained from local authorities covered a total general population of 3970903, with 545409 (13.7%) students and 60401 (1.5%) teaching and other staff. We included 15433 index cases, 49131 contacts and 4637 secondary cases reported during the school year 2020/21 across 5 regions of Germany. Further information is in the supplement (Table S1).

Infection risk in phases 2-4 ranged from 2-7.6% for the general population, 1.3-5.8% for students and 2.4-3.2% for staff (Table 2). Infection risk was lower for students and staff than the general population during periods of school closure (Figures 1-7 in S6 Figures). Student infection risk increased with student age. The under 10-year-olds show a lower risk of infection than the 10-to-14 and 15-to-19-year-olds in most phases and regions (Fig. 3). Infection risk was higher for students in more advanced school forms with data available from 2 regions (S8 Text). Staff infection risk was similar across age groups. There was a trend towards higher infection risk in special needs and primary school staff and lower risk in secondary school staff (S8 Text). Further data is in the supplemental (Table 2 in S7 Tables, Tables 1-7 in S6 Figures, S8 Text).

**Figure 3.**
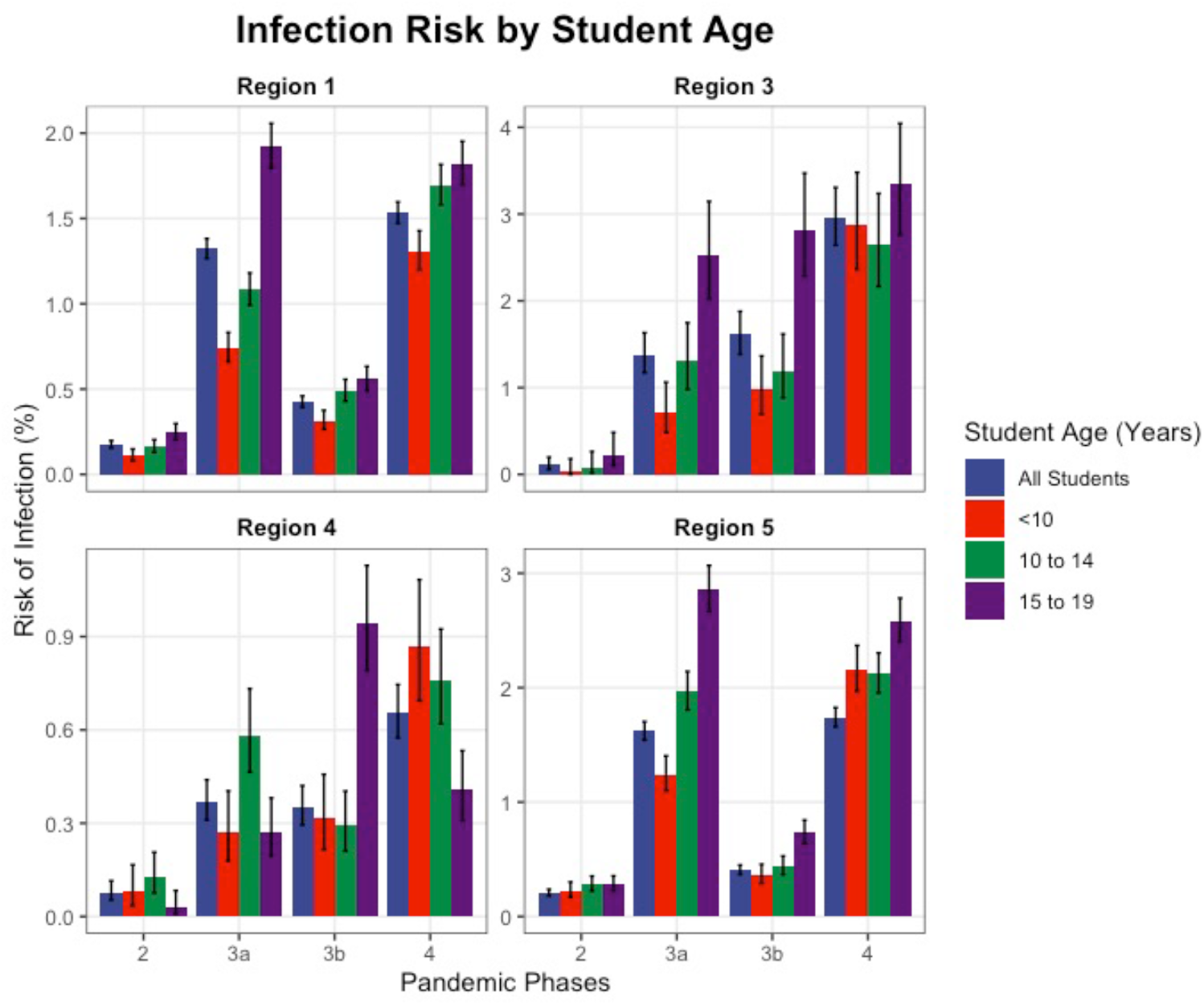
Infection risks of students by age in four regions

### Risk of infection after contact with a notified case (secondary attack rates)

Overall – irrespective of contact setting - risk of infection after contact with a notified case (SAR) for the whole observation period ranges from 4.6-12.8% across all included counties. For the different phases, ranges are 4.1-16.7% (phase 2), 4.9-9.1% (phase 3a), 5.3-19.4% (phase 3b) and 5.4-19% (phase 4). SAR increased with index as well as with contact age with data available for four regions (S9 Tables, S10 Text).

For all phases in all regions, 15-to-19-year-old student index cases had a higher SAR than under 10-year-olds and in several regions higher than 10-to-14-year-olds. SARs increased also with contact age and with more advanced school forms (S9 Tables, S10 Text).

Data from one region allows differentiation of SARs by contact settings. Whereas overall most contacts (64%) were recorded for schools as the contact area, most secondary cases (84%) were reported in household contacts (Fig. 4). Total SARs are 1.2% for contacts in schools and 23.2% for contacts in households, with 8.2% for all contact areas (S9 Tables). Regarding pandemic phases, household SARs in students are 1.5 to 2 times higher in phase 4 than phase 2 and phase 3a, whereas school specific SARs do not change significantly (Fig. 5).

**Figure 4.**
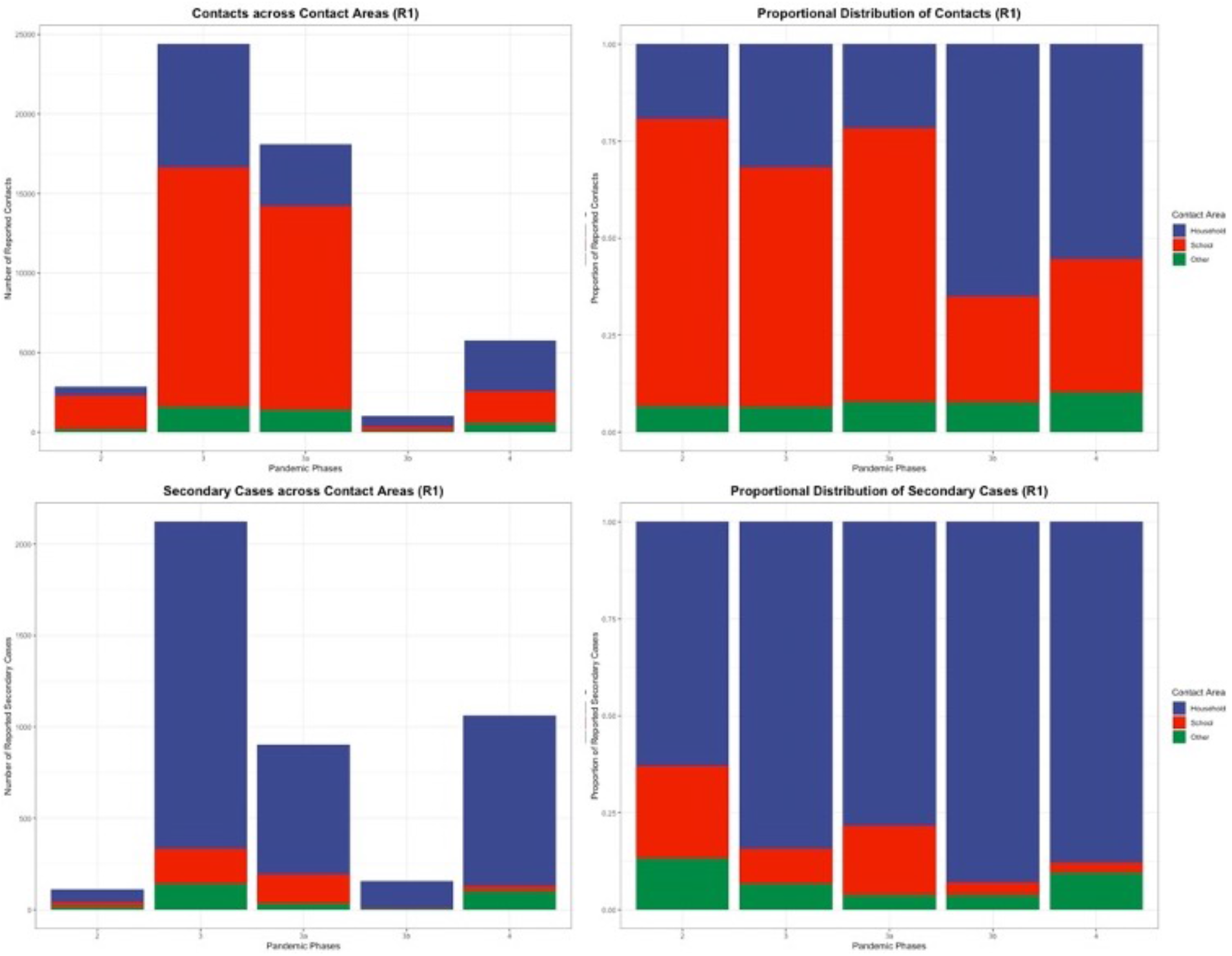
Contacts and Secondary Cases in Phases 2-4 (W21 2020 to W23 2021) (R1)

**Figure 5:**
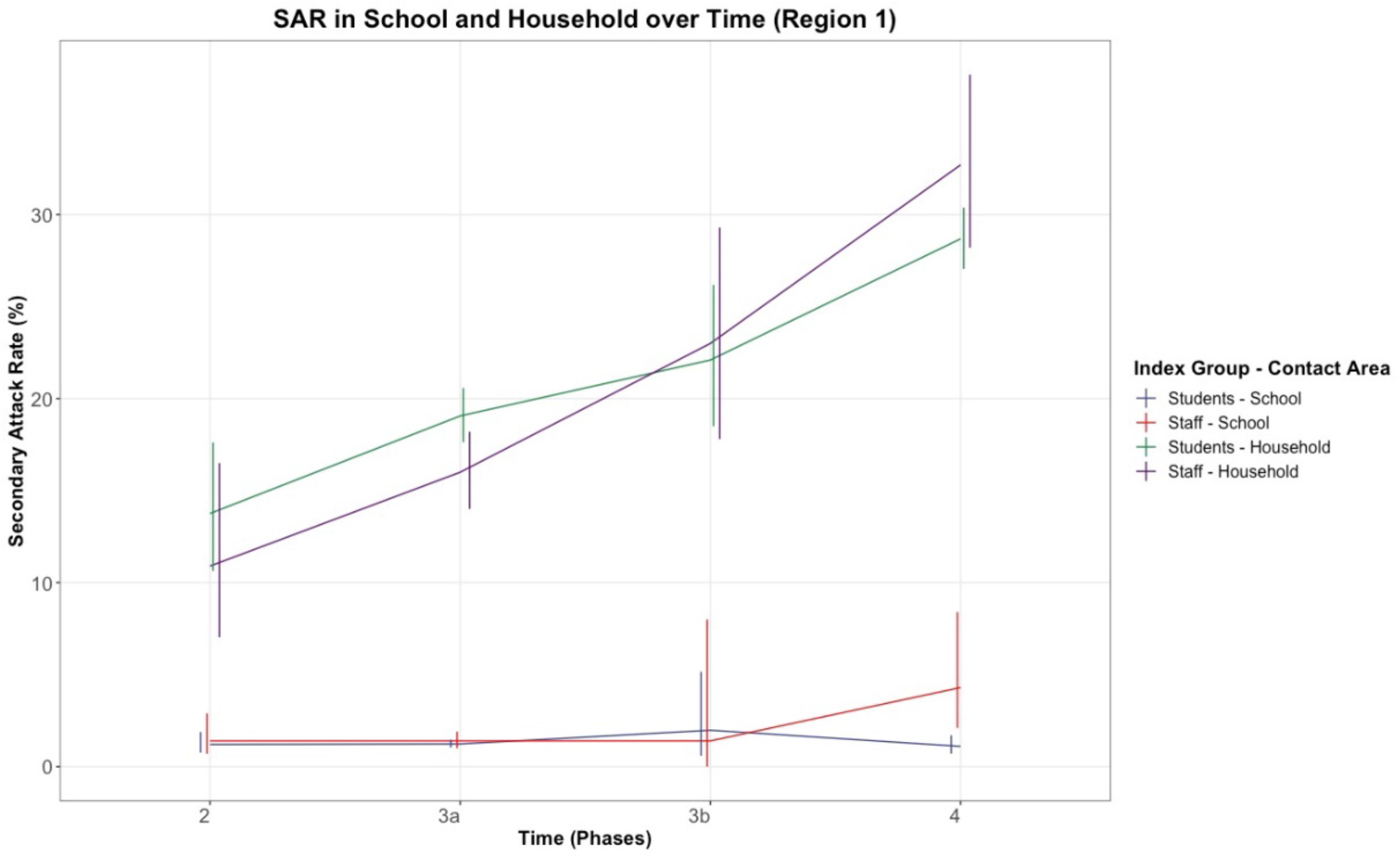
School and Household SAR in Students and Staff over Time

Absolute and proportional contacts across contact areas changed over time. Whereas in phase 3a the overall SAR was 4.9%, with schools at 1.2% SAR and 74.8% of contacts reported, and the household SAR at 19.1% with 20% of contacts, in phase 3b (school closure) the SAR changed to 16% overall as the share of household contacts was now 65.6%. Average reported contacts remained similar, with 10.6 (phase 3a) and 10.4 (phase 3b) in schools and 2.97 (phase 3a) and 2.77 (phase 3b) in households. In total, 2.2 times more contacts were reported in schools, whereas 8.8 times more secondary cases originated in households relative to schools (S9 Tables, Fig. 4 and 5).

Contact and secondary case data for staff was available from one region. Overall SAR for staff was 8%. Household SARs were 1.5 to 2 times higher in phase 4 than phase 3a and phase 2. School-specific SARs of staff members were higher for phase 4 at 4.3% compared to 1.4% in phase 3a and the average of 1.5% (Fig. 4).

Among school contacts of staff, 14% were over 18 years old. 57.7% (30 out of 52) of secondary cases of infected staff in schools occurred in this age group with a SAR of 4%, compared to 0.8% in the under 10-year-olds and 1% in the 10-to-18-year-olds.

### Effects of school-specific non-pharmaceutical measures on infection dynamics in schools during the third wave (week 09-25) using county-level data

The ordinary least squares (OLS) model for the official active cases per 100000 students and teachers as reported by the KMK showed that in counties with a mask mandate during class in all types of schools (i.e. primary schools as well as secondary schools) the average number of weekly cases was lower by 56 per 100000 cases for both the students and teachers (Table 3). The mandate was associated with a greater reduction of infection activity among students and teachers than the total population. The mask mandate in class for all types of schools was associated with a reduction of case numbers relative to the general population of 29.8% for students and 24.3% for teachers (S11 Text, Table 1). This result withstood sensitivity analysis of interaction with time periods of increasing infections with less measures or periods of decreasing infections and more measures implemented (S11 Text, Tables 2 and 3)

**Table 3.** Regression Results for Active Cases per 100k Students or Teachers, respectively, on the two-week-lags of school-specific NPIs

| Variable | Students<br>Estimate of reduction or<br>increase in notified<br>infections / 100.000<br>persons/14 days (95%-CI) | Teacher<br>Estimate of reduction or<br>increase in notified<br>infections / 100.000<br>persons/14 days (95%-CI) |
| --- | --- | --- |
| 2-week incidence per 100k inhabitants | 0.3 (0.28; 0.32) | 0.5 (0.47; 0.55) |
| Attendance in schools |  |  |
| Open schools | baseline | baseline |
| School vacation | - 8.8 (- 19.3; 1.6) | - 46.8 (- 68.5; - 25.2) |
| School closures | - 60.4 (- 73.4; -47.7) | - 91.6 (- 118.1; - 65) |
| Reduced presence in schools | - 19.3 (- 29.6; - 9.1) | - 50.3 (- 71.5; - 29.1) |
| Mask mandates |  |  |
| No mask mandate or voluntary masking | baseline | baseline |
| Partial mask mandate in some or all schools | - 17.8 (- 25.5; - 10) | 9.9 (- 5.2; 25.1) |
| Mandatory masks in all school classes | - 55.5 (- 63.4; - 47.7) | - 55.6 (- 71.6; - 39.6) |
| Testing |  |  |
| No testing in schools | baseline | baseline |
| Voluntary testing in schools | 45 (38.2; 51.9) | 17.8 (4; 41.6) |
| Mandatory testing in schools | 49.8 (41.2; 58.5) | 6.5 (-11.3; 24.3) |
| Percentage of completely vaccinated population in<br>corresponding federal state [cont.] | - 4.1 (- 4.9; - 3.2) | - 5.5 (- 7.2; - 3.8) |
| Urbanity† |  |  |
| Rural | baseline | baseline |
| Urban (RegioStar71/72) | 21.3 (11.9; 30.7) | - 44.2 (- 63; - 25.5) |
| Deprivation Index * |  |  |
| Deprivation Index 0-0.5 | baseline | baseline |
| Deprivation Index 0.51-1 | -7.8 (- 16; 0.33) | 81.5 (65.2; 97.7) |
| R² [as %] (sample size) | 37.7 (3809) | 32.7 (3979) |

see Supplementary Methods
Attendance in schools: Open Schools = 81-100% of students in presence, School vacation = 0%, School closures 1-20%, Reduced presence in schools = 21-80%
Partial mask mandate in some or all schools: mask mandates only for secondary schools, for all schools but not in class, or in class only for secondary schools
\*Deprivation Index: RKI German Index of Social Deprivation (30)
†Urbanity Index by Federal Ministry of Transport and Infrastructure; 71= metropolis, urban centre defined by infrastructure and service availability in the region; 72 = regiopolis, regional urban centre defined by infrastructure and service availability in region (31)

Lower attendance rates among students within regions were associated with reductions in infection risks of teachers and students. Vacation and reduced presence class models were associated with a greater reduction in the risk of infection for teachers than for students.

Higher vaccination coverage in the population was associated with a decrease in notified infections in students as well as teachers.

Mandatory testing in schools in our study period was associated with an average increase of 50 notifications per 100000 per 14 days among students. (Table 2, S11 Text).

Urban counties were associated with a higher number of notified infections in students and a lower number in teachers compared to rural counties (S11 Text). A high degree of social deprivation in a county was associated with higher numbers of infections in teacher (S11 Text).

### Contribution of contacts in schools to overall transmission in the population according to nation-wide data from educational agencies

We used the compartmental model described in Fig.1 and data from the KMK. Not accounting for underestimation of infections, we found a high variability in the contribution of contacts from school infections to the overall transmission during the third wave from 2-12%. Accounting for age-specific underestimation of notified infections in comparison to actual infections based on seroprevalence estimates, this range was 2-13% (Fig.6). The contribution was lowest during vacations or school closures. The peaks occurred with open schools. The W24 peak coincides with a 7-day average population incidence of 6 cases per 100000 inhabitants and the W40 peaks occurs at 65 per 100000 7-day population incidence. The local peak at W12 occurs with reduced presence class models and a population incidence of 130 per 100000 inhabitants.

**Figure 6.**
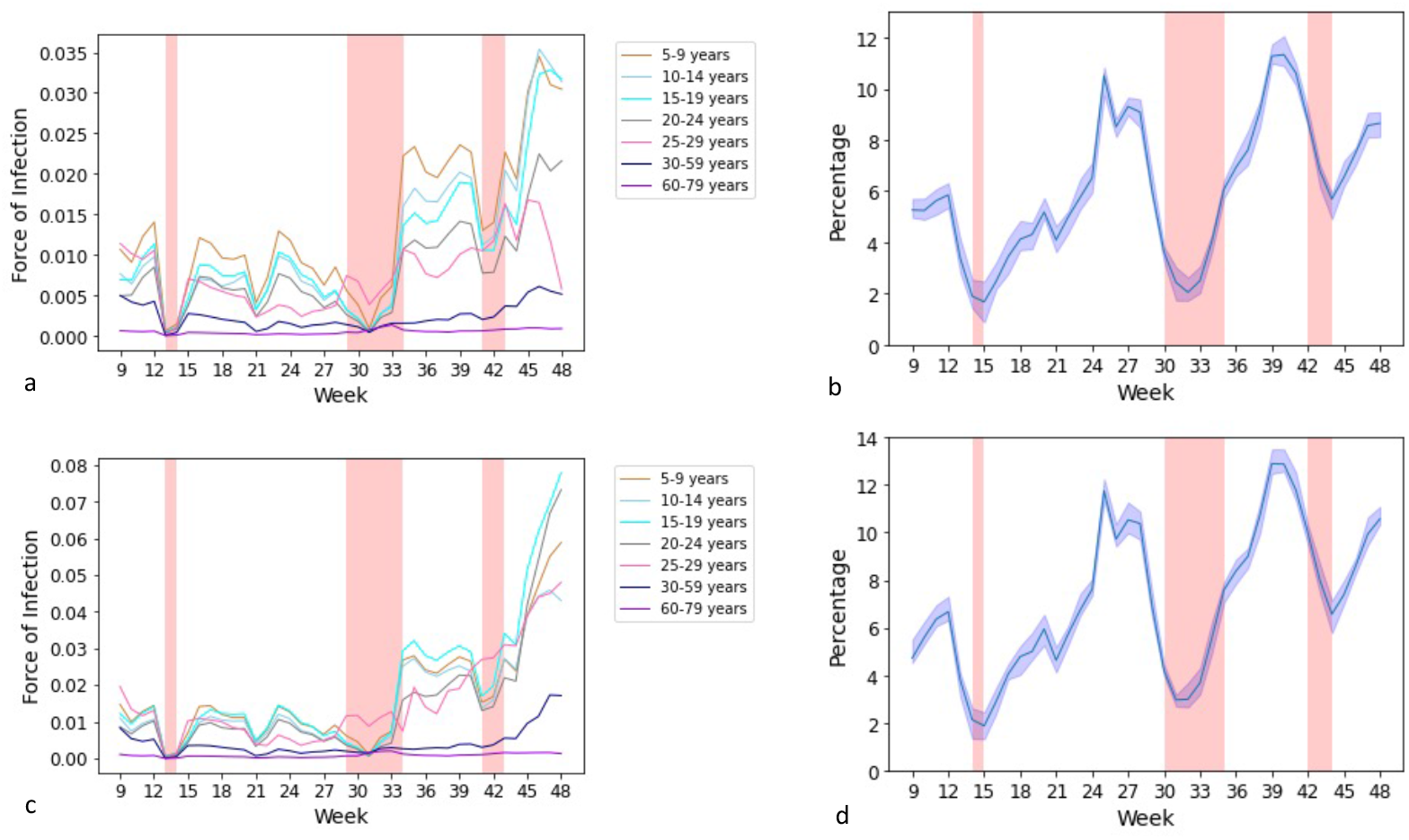
Estimated force of infection and contribution of contacts of infected persons in school to overall transmission in Germany week 9-39 2021, accounting (c&d) and not accounting for underdetection (a&b). Pink areas represent periods when the majority (at least 7) federal states have school vacations. Vacation periods in federal states in Germany differ by multiple weeks and will span periods prior and post to the areas marked. W16-21 include county-specific incidence-dependent school closures which affected less than 50% of counties in Germany

## DISCUSSION

In this study of infection and transmission risks during the pandemic in Germany, we could show that the contribution of infection dynamics in schools to that of the general population was up to 13% under hygiene and testing measures. Furthermore, NPIs such as strict mask mandates (in all schools during class) and reduced presence models are associated with an effective reduction in infection and transmission risks in schools and their contribution to population infections.

Most infections and transmission in school took place in phase 3a when schools operated in normal class models with some cloth mask rules and general hygiene concepts. Teachers showed a higher risk of infection than the general population. Infection risk for students increased with age, as previously shown(17,19). During winter 2020/21 school closures (phase 3b), total reported infections and transmission in schools were low and infection risk for both students and staff was lower than the general population. Meanwhile, more time spent in the household lead to longer exposure and thus a higher SAR in household members, yet fewer secondary cases in total.

In phase 4 schools reopened with stricter NPIs including reduced presence class models and stricter mask mandates with testing and incidence-dependent school closures introduced later on. Meanwhile, the prevalence of the more transmissible B.1.1.7 (alpha) variant rose in Germany. Infection risks for students and teachers were similar to that of the population. School-setting SAR of both students and teachers remained low (<5% during phases 2-4), whereas household SAR more than doubled. The stricter NPIs likely counteracted the higher transmissibility of the alpha variant in schools.

This hypothesis is supported by our regression analysis. We estimate a reduction by 24-30% of infection risk for both students and teachers relative to the population and by more than 55/100.000 per 14 days in absolute case numbers in those counties implementing mandatory surgical mask wearing during class in all schools. Similar observations have been made in Arizona, USA, where the odds of school-setting outbreaks of SARS-CoV-2 were 3.5 times higher in schools without a mask policy(32).

Furthermore, all interventions reducing attendance rate of students (rotating and distance classes as well as school closures and vacations) reduced the risk of infection of students and teachers. Concurrently, we see a higher risk of infection for students and teachers in counties with higher deprivation indices. This was known for Germany, though not specifically for schools(33, 34).

These findings are reflected in our analysis on the contribution of schools to the general population infection dynamics. School vacation and closures reduced this contribution to as low as 2%. Previous modelling for the first and second wave in Europe showed that school closures resulted in >30% and <15% reductions of transmission(8, 35). The peak at 12% contribution in W24 occurs with open schools, loosening population NPIs and loosening but still strict hygiene and testing measures in schools. Meanwhile, the lower peak at 7% early in phase 4 occurred during reduced attendance class models and stricter NPIs.

During phase 5 we observe a continuation but loosening of mask and testing mandates with full attendance classes in a low population infection environment. The crude infection risk ratio of teachers relative to the population in phase 5 remained steady while it was higher for students than the population. This is likely due to vaccination coverage rising in the adult population and mandatory testing in schools. Whereas teachers as a vocational group were vaccinated with priority, vaccination for students age 12-17 was not available until W33. Our regression analysis showed the protective effect of higher vaccination rates in the population as well as a higher number of notified cases among students with mandatory testing, likely through reducing underdetection. Summer vacations reduced the contribution to population infection dynamics to a trough of 3%. Peaks occurred at 13% during open schools in normal presence classes.

We further found that SARs are higher both with higher age of the of index cases as well as contacts. SARs have previously been reported only for early phases of the pandemic from German public health agency data, showing SARs below 2%(17). The household SARs observed are in line with previous findings for the general population(36). The majority of secondary cases of both students and staff happened in households. In schools, age-specific SARs show that transmission took place mostly from staff to staff and from student to student more than student to staff. This is in line with previous findings(16).

Interestingly, in sensitivity analyses (S11 Text) of the regression analysis, we find a decrease in infection risk of primary school students when attendance in school increased, whereas attendance is positively correlated with infection risk for all other student and staff groups. Previous studies showed a similar effect for the end of vacations(37). The infection risk in school for primary school students is possibly lower due to stricter adherence to the rules in schools and by parents and less time spent in households with a higher transmission risk.

Limitations of our work are inherent in the notification process to both public health and educational agencies as well as the gathering of aggregate county-specific data (S12 Text). Both notification data itself as well as contact data is an underestimate of the actual infection dynamic. We attempted to account for underdetection with age-specific estimates taken from seroprevalence studies. This could make our estimate of contribution of school-contacts to overall transmission a potential underestimate. Residual confounding in the regression analysis is a possibility as we use aggregate county or school measures and did not have full access to individual-level confounding factors, e.g., on distribution of parental professions or industrial make-up of the counties included. We tried to include the most important confounding factors on infection dynamics in the population as well as deprivation and urbanity of counties in the analysis. SARs are limited by several factors. Contact tracing is inherently limited by both interviewer capacity and ability and interviewee memory and honesty, however with schools and households as contained domains the error margin is limited. Among contacts, the data did not indicate whether these were still susceptible to infection. However, it can be assumed that biases are similar in each region as they are situated within the same country and timeline of events. A reduction in the proportion of the susceptible population through vaccination or infection is assumed relevant from March 2021, where it is included as a parameter in regression analysis. All considered, the SARs presented here described further transmission after cases have been detected in the school with limitations.

Despite these limitations we conclude that school setting contribution to overall transmission in the population is relevant, but variable over different time periods in the pandemic and responsive to NPIs. In Germany, school-related NPIs, in particular masking and reduced attendance, have been successful in mitigating the spread of the virus among both students and teachers.

## Data Availability

(1) All population infection data is publicly available from the RKI at: https://survstat.rki.de/Default.aspx (1) KMK federal state data is publicly available at: https://www.kmk.org/dokumentation-statistik/statistik/schulstatistik/schulstatistische-informationen-zur-covid-19-pandemie.html (2) KMK county-level data is not publicly available. Data can be obtained on request by contacting the KMK directly at: https://www.kmk.org/kmk/information-in-english/contact.html (3) Regional health and education authority data was obtained through data sharing agreements as specified in the approved ethics protocol directly from the multiple government agencies. Data is based on public health notification records and case-specific data collected in the notification and contact-tracing process. Data can be obtained on request. Interested parties may contact the corresponding author, who will provide points of contact for the relevant agencies. Signature of a data sharing agreement or data sharing addendum with Helmholtz Centre for Infection Research and/or the providing government agency will be required. Data availability is subject to approval by the providing agency. Agencies include the health agencies of Cologne https://www.stadtkoeln.de/service/adressen/00112/index.html Stuttgart https://welcome.stuttgart.de/item/show/547988/1/dept/1475?site=welcomecenter Altenburger Land https://www.altenburgerland.de/sixcms/detail.php?id=11267&_lang=de Konstanz https://www.lrakn.de/service-und-verwaltung/aemter/gesundheit+und+versorgung/gesundheitsamt Gutersloh https://service.kreis-guetersloh.de/detailansicht/-/egov-bis-detail/einrichtung/332/show Bautzen https://www.landkreis-bautzen.de/landratsamt/organisation/gesundheitsamt/46 and the education agency of Hamburg https://www.hamburg.de/bsb/

## ACKNOWLEDGMENTS

We would like to thank the education ministries in the federal states in Germany for gathering the relevant data from federal states and counties and of course all staff in schools and regional school agencies who collected data. In particular, we would like to thank Dr. Marco Mundelius and Dr. Paula Henselin from the statistical commission of the Standing Conference of Ministers of Education and Cultural Affairs (KMK) in Germany for support in data acquisition and explanation. We appreciate the help of Timon Hellwagner, who gave valuable advice on geographical measures used in our regression analysis. We thank Anna Wolff und Michael Buess from the DIKOMA Team and the whole team responsible for COVID Cases in Schools and Day Care Facilities of the Public Health Agency in Cologne. We thank the Authority for Education and Vocational Training (Behörde für Schule und Berufsbildung), Free and Hanseatic City of Hamburg – especially the COVID-19-taskforce – for data provision. We thank Uwe Melzer, Landrat, and Uwe Fischer, data protection officer, for their kind support. We would like to thank Paul Glaßner, Fabian Peters, Gabriel Huck of Public Health Department Konstanz for their support. We thank our colleagues Cordelia Fischer, Florian Hölzl and Kerstin Gronbach and their teams for collecting data and we thank Marion Silo, Erich Horntasch and Bettina Huber responsible for programming and data analyses in the Public Health Department Stuttgart.

## ETHICS STATEMENT

Ethics approval was obtained by the IRB of the Medical University Hannover (Ethical approval N°9609_BO_K_2021, Hannover). Data was collected by public agencies in all cases in line with public regulation on data privacy. Data was anonymised prior to transfer. Data was analysed anonymously. Data study complies with EU-GDPR as it is a data evaluation for research in the public interest (§ 89 EU-GDPR).

