## Supplement for "Infection and transmission risks in schools and contribution to the COVID-19 pandemic in Germany – a retrospective observational study using nation-wide and regional health and education agency notification data"

### SUPPLEMENTARY FILES

SUPPLEMENTARY FILES 1

**S1 Table. Data Sources** 2

**S2 Table. Overview of Regional Data Collection** 5

**S3 Text. Additional Methodology** 8

**S4 Figure. Overview of Incidences across RKI Phases of the Pandemic** 11

**S5 Tables. Infection Control Measures at the School-Level** 12
Table 1: Infection Control Measures for Schools in Region 1 12
Table 2: Infection Control Measures for Schools in Region 2 14
Table 3: Infection Control Measures for Schools in Region 3 16
Table 4: Infection Control Measures for Schools in Region 4 18
Table 5: Infection Control Measures for Schools in Region 5 20

**S6 Figures. Incidences of Population, Students and Staff across regions based on local agency data** 22
Figure 1: Incidences of Population, Students and Staff over time in Region 1 22
Figure 2: Incidences of Population, Students and Staff over time in Region 2 23
Figure 3: Incidences of Population and Students over time in Region 3 24
Figure 4: Incidences of Population and Students over time in Region 4 25
Figure 5: Incidences of Population and Students over time in Region 5 26
Figure 6: Incidences of Students (All Regions) 27
Figure 7: Incidences of Staff (All Regions) 28

**S7 Tables: Infection Risks Across Regions and Subgroups** 29
Table 1: Infection Risks, Crude Risk Ratios and Quarantine Risk Across Federal States (KMK Data) 29
Table 2: Infection Risk Across Regions and Subgroups (Local Agency Regional Data) 31

**S8 Text. Additional Results 1: Infection Risk** 33

**S9 Tables. Secondary Attack Rates** 34
Table 1: Secondary Attack Rates in Students by Region, Phase and Age (Local Agency Regional Data) 34

**S10 Text. Additional Results 2: Secondary Attack Rates** 39

**S11 Text. Additional Results 3: Effects of NPIs in Schools** 42

**S12 Text. Additional Limitations** 50

**S13 Text. Supplement References** 52

### S1 Table. Data Sources

*Table 1 Data obtained from public health agencies on SARS-CoV-2 infections in students and teachers and their contacts*

| | Infection Data provided by local authorities | | | | | | Standing Conference of Ministers of Education and Cultural Affairs | |
|---|---|---|---|---|---|---|---|---|
| | Total | Region 1 (R1) | Region 2 (R2) | Region 3 (R3) | Region 4 (R4) | Region 5 (R5) | NUTS-3 level data | Federal state level data |
| Location | | Urban | Urban | Rural | Rural | Urban | 275/401 NUTS-3 | 16/16 federal states |
| Population | 3970903 | 1088040 | 1899160 | 90118 | 285325 | 608260 | | |
| - students | 545409 | 149428 | 257216 | 10046 | 34369 | 94350 | 5092252 | 13382111 |
| - staff (teachers) | 60401 | 38314 | 36147 (22807) | NA | NA | NA | (624944) | (818409) |
| Data source | | Health agency | School agency | Health agency | Health agency | Health agency | School agency | School agency |
| Time from | 07.03.20 | 09.03.20 | 04.08.20 | 10.03.20 | 06.04.20 | 07.03.20 | 01.03.2021 | (1) 09.11.2020† (2) 22.02.2021 |
| Time until | 08.07.21 | 01.05.21 | 14.01.21 | 09.06.21 | 14.04.21 | 08.07.21 | 27.06.2021 | (1) 13.12.2020 (2) 11.10.2021 |
| Duration (d) | 1898 | 418 | 163 | 456 | 373 | 488 | 119 | (1) 35 (2) 224 |
| Notified infections | 15433 | 7248 | 3575 | 559* | 511 | 3540* | 95215 | (1) 113068 (2) 224600 |
| - students | 12814 | 5108 | 3096 | 559 | 511 | 3540 | 85788 | (1) 96970 (2) 207706 |
| - staff | 1719 | 1240 | 755 (479) | NA | NA | NA | 9427 | (1) 16098 (2) 16894 |
| **Contacts** | 49131 | 27508 | NA | 2820 | 2256 | 16547 | NA | NA |
| - of students | 43238 | 21615 | NA | 2820 | 2256 | 16547 | NA | NA |
| - of staff | 5893 | 5893 | NA | NA | NA | NA | NA | NA |

*Cases derived from age. Cases from 6- and 19-year-olds halved according to student demographics (1)

†No data was reported during winter vacation and early 2021 school closures

Region 1

Data was provided from the sub-team for schools, kindergartens and communal accommodations at the local health authority. The data was filtered for cases of students and staff working at schools only. Teachers and other school staff were not differentiable. Cases were marked as to age and the type of school they belonged to. Student and staff data was analysable both by age and type of school. Contacts and secondary cases were marked as to whether they occurred in school, at home, or "other".
The data suffered from being entered differently by different staff and over time and in part field markings were not standardized. In close conversation with the health authority data entering errors and discrepancies could be resolved and relevant were fields standardized.

Denominators for student and staff numbers at Cologne schools were sourced from:

Ministerium für Schule und Bildung des Landes Nordrhein-Westfalen. (2020). Das Schulwesen in Nordrhein-Westfalen aus quantitativer Sicht 2019/20

Stadt Köln, Amt für Stadtentwicklung und Statistik, Amt für Presse- und Öffentlichkeitsarbeit. (2020). Statistisches Jahrbuch 2019, 96. Jahrgang. Kölner Statistische Nachrichten 1/2020

Statistisches Landesamt Nordrhein-Westfalen. (2018). Regionalisierte Schüler-Modellrechnungen in Nordrhein-Westfalen – Schülerinnen und Schüler sowie Schulabgängerinnen und Schulabgänger Schuljahr 2018/19 bis 2033/34

Region 2

This data set was the only one provided by an educational authority. It included cases among students and staff notified by schools, not the health authority but was managed in close interaction with the health authority to follow implemented interventions. Data was recorded for students and teaching as well as other staff separately. Cases' age was not recorded, only the type of school was marked and for students the school year group. No contacts or secondary cases were recorded.

Denominators for student and teaching staff were sourced from:

Behörde für Schule und Berufsbildung. (2021). BQ12 Systemanalysen und Berichterstattung: Das Schuljahr 2020/21 in Zahlen
https://www.hamburg.de/schuljahr-in-zahlen/ last accessed 09.08.2021
The Behörde für Schule und Berufsbildung provided data on numbers of staffs and students from all types of schools in 11/2021. No breakdown of non-teaching staff by school type was available, thus they were included in the total only. No breakdown of private school teaching staff was available, thus they were included in the total only.

Region 3

The data were provided by the local health authority as an export of the pandemic management tool SORMAS. The data did not include a marking of cases as students or school staff, however, outbreaks and clusters at schools were recorded. Data was thus analysed by age and affiliation. Student cases were filtered by age from 6 to 19; to determine an approximation of the case count in the student population, cases among 6- and 19-year-olds were each halved in line with their proportional share of students in the age group. Contacts and secondary cases were collated from index cases by age and as listed in outbreaks/clusters in schools. Contact areas were not recorded.

Denominators for students were sourced from:
Thüringer Landesamt für Statistik. (2021). Bevölkerung der Gemeinden, erfüllenden Gemeinden und Verwaltungsgemeinschaften nach Altersgruppen in Thüringen. https://www.statistik.thueringen.de/datenbank/TabAnzeige.asp?tabelle=gg000103%7C%7C last accessed 08.09.2021

Region 4
The data were provided by the local health authority as an extract of cases marked as students in their total case database. Data included students only by age, not the type of school attended. Information on the number of contacts and secondary cases recorded for each case was provided, however with no information on contact characteristics or contact areas.

Denominators for students were sourced from:

Statistisches Landesamt Baden-Württemberg. (2020). Bevölkerungspyramiden: Bevölkerung nach Alter und Geschlecht (absolut) 2020 Konstanz, LKR *and* Schüler und Schulen seit 1987/88 nach Schularten Konstanz, LKR
https://www.statistik-bw.de/Bevpyramiden/ last accessed 08.09.2021

Region 5
The data were provided by the local health authority as an export of their pandemic management software. Data was thus analysed by age and affiliation. Student cases were filtered by age from 6 to 19; to determine an approximation of the case count in the student population, cases among 6- and 19-year-olds were each halved in line with their proportional share of students in the age group. Contacts and secondary cases were collated from index cases by age. Contact areas were not recorded.

Denominators for students were sourced from:

Statistisches Landesamt Baden-Württemberg. (2020). Bevölkerung nach Altersgruppen und Geschlecht (absolut) Stuttgart, SK *and* Schüler und Schulen seit 1987/88 nach Schularten Stuttgart, SK
https://www.statistik-bw.de/Bevpyramiden/ last accessed 08.09.2021

#### S2 Table. Overview of Regional Data Collection

Texts were provided by the local authorities on request and translated into English when necessary. The texts are a self-representation of the regions themselves.

| Region 1 |
| --- |
| All patients with detection of SARS-CoV-2 were contacted and interviewed by the local health department. The patients and their close contacts were registered in DIKOMA (Digitales Kontaktmanagement; digital contact management—a database developed by the Department of Information Processing by the city of Cologne (2).<br>The definition of a first grade contact person follows the recommendations of the RKI (e.g. >10 min close contact of under 1.5m distance) which also developed through the pandemic. Particularly the mandatory use of masks and the different safety standards changed the definition sometimes.<br><br>Interviews took place in a standardized form. However certain questions were added during the course of the pandemic (e.g. vaccinations). In addition, in times of high case numbers not every contact person could be interviewed via telephone but had to fill in an online database. The periods in which the contact tracing leaked on accuracy are overlapping with the covid waves.<br>The quality of the interviews depends on the experiences of the person in charge.<br>Data is analyzed retrospectively.<br>Contact tracing in schools took place in a differentiated matter as previously described (2). |
| Region 2 |
| a) What were the conditions for a case to be included on this list?<br>- The condition for a case was the student or teacher self-reporting a positive SARS-CoV-2 PCR test to the school. The list of cases includes all infections reported by the school to the education authority.<br>b) Are all school types (general education schools, special schools, vocational schools) included?<br>- All types of schools are included<br>c) Are all cases of students included, or only those who were at school during the infectious period?<br>- All cases of students and teachers reported to schools and the education agency.<br>d) How were they identified as belonging to students? (Notification by schools, entry in telephone call for contact tracing, ...) and did this process change over the period?<br>Only cases of students or teaching staff were included. No contacts outside schools identified.<br>- Completeness of the data<br>a) In your opinion, is the infection data complete, or what could be the reasons for completeness?<br>Underestimation of cases in comparison to cases reported to the public health agency is possible.<br>b) In your opinion, is the contact tracing data complete, or what could be the reasons for completeness? (e.g. at times of high incidence due to lack of capacity).<br>See above |
| Region 3 |

| |
|---|
| The data of this county was collected by the using the SORMAS and SORMAS-X software continuously from March 3, 2020 until end of the observation period. During this time there was a dramatic increase in the number of infections starting in December 2020, so that staff had to be recruited and trained. These changes in staff may have affected the quality of data in particular in the most dramatic months December 2020 and January 2021. In this county there are 21 primary schools, 12 secondary schools, and 5 high schools, as well as 3 special schools. |
| Region 4 |
| a) What were the conditions for a case to be included on this list?<br>- The condition for a case was a positive PCR test result.<br>b) Are all school types (general education schools, special schools, vocational schools) included?<br>- All types of schools are included<br>c) Are all cases of students included, or only those who were at school during the infectious period?<br>- All cases of students<br>d) How were they identified as belonging to students? (Notification by schools, entry in telephone call for contact tracing, ...) and did this process change over the period?<br>- Individuals were marked as belonging to pupils during contact tracing. The process remained unchanged over the period.<br><br>- Completeness of the data<br>a) In your opinion, is the infection data complete, or what could be the reasons for completeness?<br>- The infection data is complete<br>b) In your opinion, is the contact tracing data complete, or what could be the reasons for completeness? (e.g. at times of high incidence due to lack of capacity).<br>- Contact tracing data was not complete at the beginning of the second wave due to lack of staff capacity. By the end of 2020, the data were complete again.<br><br>After the infection incidence in the age cohort under investigation was at a manageable level due to early school closures in spring 2020, a clear increase can be seen at the end of the summer holidays. The increase in new infections is particularly noticeable at the end of September and the beginning of October 2020. It must be noted that due to the data structure stringent statistical analysis cannot evidence this.<br>Following this, a continuous increase in the number of infections occurred, whereby the rate of new infections in the analysed age cohort was not conspicuous in relation to the overall population mean. However, the increase became less steep after the introduction of alternate teaching from grade eight and the subsequent announced school closure.<br>A renewed increase in the incidence of infection is seen after 19 January 2021.<br>With the onset of the third wave in spring 2021, there was a rapid increase after steady growth until the late weeks of February. It can be assumed that this was due to the displacement of the SARS-CoV-2 wild type by variant of concern B.1.1.7. |
| Region 5 |
| a) What were the conditions for a case to be included on this list? |

- The condition for a case was a positive PCR test result reported to the public health agency.
b) Are all school types (general education schools, special schools, vocational schools) included?
- Yes, all types of schools are included
c) Are all cases of students included, or only those who were at school during the infectious period?
- All cases of students are included.
d) How were they identified as belonging to students? (Notification by schools, entry in telephone call for contact tracing, ...) and did this process change over the period?
- Individuals were marked as students during contact tracing. The process remained unchanged.

- Completeness of the data
a) In your opinion, is the infection data complete, or what could be the reasons for completeness?
- The infection data is complete
b) In your opinion, is the contact tracing data complete, or what could be the reasons for completeness? (e.g. at times of high incidence due to a lack of capacity).
- Contact tracing data is not complete from the second wave onwards due to lack of staff capacity.

On 4 March 2020, the Stuttgart Health Department received the first report of a person testing positive for SARS-CoV-2. By the end of the month, there were already over 700 confirmed cases, most of them were returning travelers from ski resorts in South Tyrol and Austria. The first days of the pandemic were marked by organisational challenges. Already on 12 March, an individually programmed database was launched, which made it possible to manage the rapidly increasing number of cases in a structured way.
On 13 March, the city banned the operation of facilities and catering establishments with immediate effect to protect the population; thus, public life was practically paralysed. On 9 March, a last football match of VfB Stuttgart had still taken place in the packed Mercedes-Benz Arena.
Schools and day-care centres were closed from 17 March 2020, and home schooling started in Baden-Württemberg. This first wave ebbed significantly after 2 months. First loosening of restrictions occurred in early May, e.g., schools opened for graduating classes.
The second wave (October 2020 to mid-February 2021) was characterised by outbreaks in nursing homes and other institutions, most deaths and a renewed state-wide school and day-care centre closure from 16 December. In this wave, in addition to the elderly and those in need of care, it was mainly vocational and high school students who were affected. At the end of February, the first teachers and educational staff were vaccinated against SARS-CoV-2 and mobile testing was offered in day-care centres and schools.
In the third wave (March to June 2021), the virus variant Alpha prevailed. For the first time, many infants and primary school children were affected. In May, the 7-day incidence for Stuttgart peaked at 224.
With the end of the summer holidays in Baden-Württemberg, the fourth wave began in Stuttgart at the beginning of September, dominated by the highly contagious delta variant. Schoolchildren, adolescents and the middle age group of adults were mainly affected. Among the adolescents, however, it is more the lower grades and fewer adolescents from higher grades. We assume that this group is now protected by vaccination.

#### S3 Text. Additional Methodology

##### 3.1 Aims

The primary aim of this study is a better understanding of the risk of infection, the transmission of SARS-CoV-2 in schools, and the contribution of in-school contacts to overall cases in the population over different phases of the pandemic in Germany. Moreover, we assess the reduction of risks of infection and transmission associated with specific measures taken in schools.

Specific aims of the study are an analysis of the

1) risk of infection in students and staff in schools,
2) risk of notified infection for a contact person (*secondary attack rate*) of a notified student and staff index cases, and
3) effect of measures taken in schools to reduce the transmission of SARS-CoV-2.

Whereas health and education agency data were used for Aims 1) and 2), data of the Standing Conference of Ministers of Education and Cultural Affairs in Germany (*KMK*) at NUTS-3 level was used for aim 3).

##### 3.2 Health/education agency data

Whereas the national Robert Koch Institute receives core notification data from local health authorities, more detailed data are often collected locally to support public health management. Of around 400 local health agencies in Germany, 20% were contacted since March 2021, of which 7 agreed to provide their school-specific data, of which 5 were provided in time for analysis. These are numbered Region 1-5 (R1-5) according to their order of receipt. Supplementary S1 and S2 Tables summarise locations, and data items provided. Data was anonymised prior to transfer. Items included for analysis were the date of case notification, age of the index case, contacts and secondary cases (R1, R3-5), contact age (R1, R3, R5) and contact area (R1). Data was requested from February 2020 to March 2021 and was provided as available at the time of data sharing. The data was cleaned, and discrepancies and questions cleared with the providing agency.

Population infections and incidences were sourced from Robert Koch Institute (RKI) survstat (3) and described as incidence per 100.000 inhabitants. Student and staff infection risk and incidences were calculated with school population data from regional or federal state authorities, analysed by age and school form as percentages where possible (Supplementary Table 1) and described.

Secondary attack rates (SARs) as percentages were calculated from data where possible.

Data was analysed per calendar week (W) as the main time unit in German state agency publications and avoids the weekly seasonality of data reporting (4) (no school on weekends, some local health agencies in Germany do not report data on weekends or Sunday). The pandemic phases were adapted from the RKI (5), focusing on phases 2 (W21-39, 2020; p2), 3 (W40, 2020 – W8, 2021; p3) and 4 (W9-23, 2021; p4). Additionally, we subdivided phase 3 into 3a (W40-51, 2020; p3a) and 3b (W52, 2020 – W07, 2021; p3b) to better subcategorise phases by school public health policy intervals and adapted the phase 3/4 transition to school reopenings (W07/08 instead of W08/09, all 2021).

Confidence intervals for binomial values were calculated using the Agresti-Coull approach (6). Data were analysed in R Studio version 1.3.959.

3.3 Data of the KMK

We received weekly data on known infections among students and teachers on the NUTS-3 level (*Kreise* or *Districts*) by the KMK for 14 of the 16 German federal state (except Bavaria and Hesse; Berlin and Hamburg are both counties and federal states) for calendar weeks 9-25, 2021 (7, 8). Moreover, we collected data on school-related non-pharmaceutical interventions (NPIs) during that period (see Section 2.4). To control for the overall infection activity during that period, we downloaded weekly incidences per 100000 inhabitants for the districts for the total population of said districts (3). To account for socioeconomic effects, we used an index derived by Kroll (9). The current values this index for the year 2015 were provided by Kroll (10). Moreover, we accounted for potential geographical effects of the districts, i.e. we may witness higher ceteris paribus (c.p.) infection risks for students who are living in highly rural areas and need to commute to school using public transports, which could increase their risk of infection. To test these effects on infection risks of students and teachers, we discriminated all districts according to the so-called RegioStar7 categorisation by the Federal Ministry of Transport and Digital Infrastructure, which defines 7 different categories of LAU regions (11).

3.4 Infection control measures in schools

In Germany, federal states maintain sovereignty over education policy resulting in 16 different approaches to school infection control at any one time. School-specific public health interventions were sourced from federal and local state and educational authority websites between 23 February and 11 August 2021 (infection control policies; laws; decrees; frameworks for hygiene measures; information to schools, parents and students; press releases; other forms of publicly available documents). Only state-issued publications were used; press reports or third-party sources were not included, as were non-publicly available materials. Measures affecting attendance (presence, rotating, distance class), masks, tests and technical aids (air filters, $CO_2$-sensors) were included and coded for regression analysis (S5 Tables, S3 Text Table 1, S11 Text). Measures describe policy-level interventions and not school-level application thereof. Sources for infection control measures are listed in the Supplement References.

*Table 1: Coding of school infection control policies for regression analysis*

| | |
|---|---|
| Category I | Attendance in schools |
| a - baseline | Open schools, normal presence teaching |
| b | School vacation |
| c | School closures |
| d | Reduced presence in schools, alternative class organization e.g. rotating classes and distance learning |
| Category II | Mask mandates |
| a - baseline | No mask mandate incl. voluntary masking |
| b | Partial mask mandate in some or all schools |
| c | Mandatory mask wearing in all school classes, including primary schools |
| Category III | Testing at schools |
| a - baseline | No testing in schools |
| b | Voluntary testing in schools |
| c | Mandatory testing in schools |
| Vaccination | Percentage of completely vaccinated population in corresponding federal state |
| Urbanity | RegioStar Index† |
| a - baseline | Rural |
| b | Urban (RegioStar71/72) |
| Deprivation | German Index of Social Deprivation* |
| a - baseline | Deprivation Index 0-0.5 |
| b | Deprivation Index 0.51-1 |

Attendance in schools: Open Schools = 81-100% of students in presence, School vacation = 0%, School closures 1-20%, Reduced presence in schools = 21-80%

Partial mask mandate in some or all schools: mask mandates only for secondary schools, for all schools but not in class, or in class only for secondary schools

*Deprivation Index: RKI German Index of Social Deprivation (9,10)

†Urbanity Index by Federal Ministry of Transport and Infrastructure (11)

S4 Figure. Overview of Incidences across RKI Phases of the Pandemic

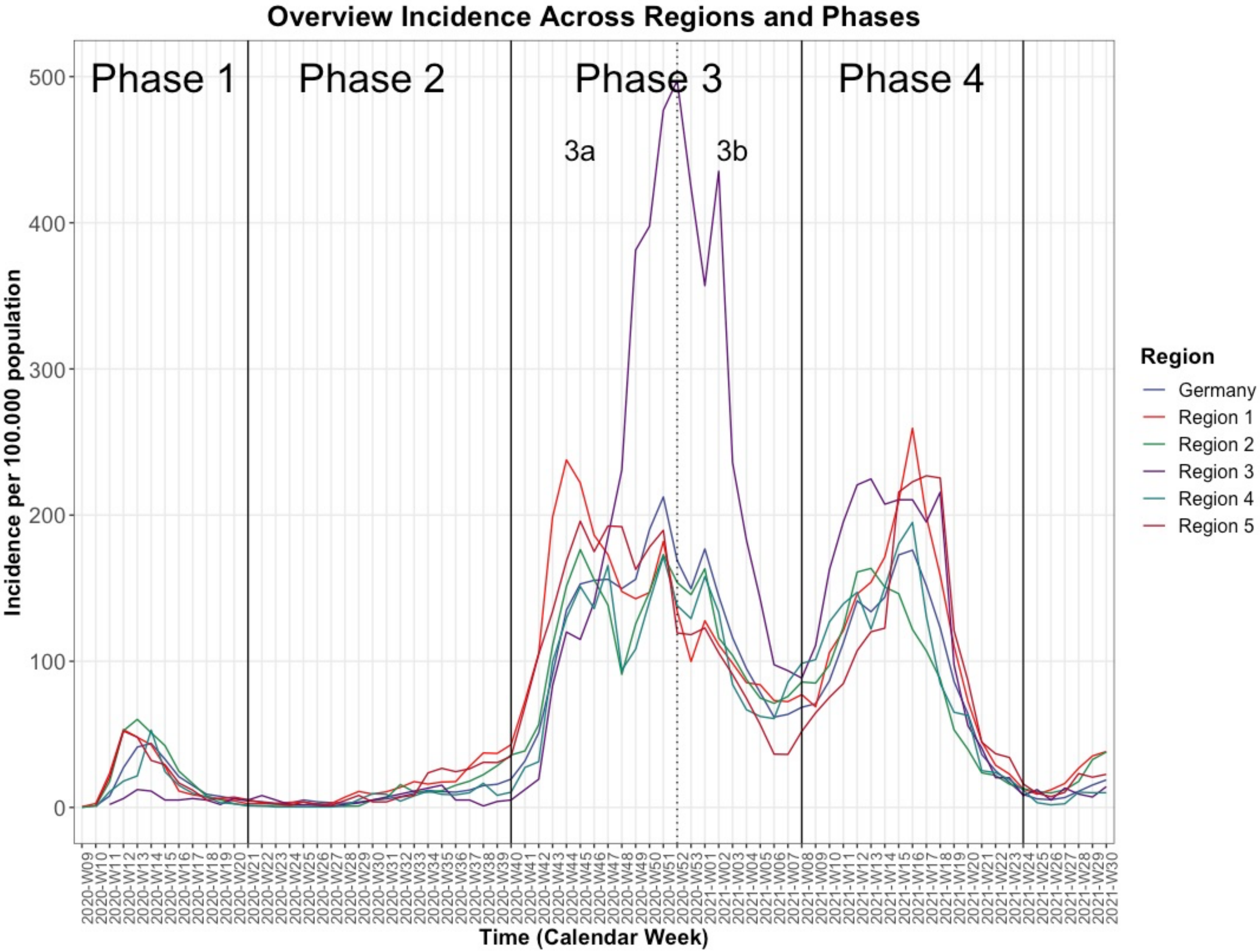

Based on Schilling et al (5).
Data sourced from RKI (3)

#### S5 Tables. Infection Control Measures at the School-Level

##### Table 1: Infection Control Measures for Schools in Region 1

| Calendar Week | Incidence Population | Incidence Students | Primary Schools (PY) | Secodnary Schools (SS) | Final Years (FY) | Measures |
|---|---|---|---|---|---|---|
| 2020 09 | 0.49 | 0.00 | | | | |
| 10 | 3.32 | 0.67 | | | | |
| 11 | 23.51 | 0.67 | | | | |
| 12 | 53.84 | 0.00 | | | | School closure |
| 13 | 47.89 | 0.00 | | | | School closure |
| 14 | 43.7 | 0.67 | | | | School closure |
| 15 | 29.85 | 0.67 | | | | School closure |
| 16 | 17.17 | 0.67 | | | | School closure |
| 17 | 8.68 | 1.34 | | | | School closure |
| 18 | 7.12 | 0.67 | | | | |
| 19 | 5.75 | 0.00 | | | | - Cloth mask recommended in all schools (not in class) |
| 20 | 4.49 | 2.01 | | | | |
| 21 | 2.54 | 0.00 | | | | |
| 22 | 2.54 | 2.01 | | | | |
| 23 | 2.63 | 3.35 | | | | |
| 24 | 3.51 | 2.68 | | | | |
| 25 | 3.8 | 1.34 | | | | |
| 26 | 1.85 | 0.00 | | | | |
| 27 | 2.63 | 0.67 | | | | Holidays |
| 28 | 7.02 | 2.01 | | | | Holidays |
| 29 | 10.92 | 7.36 | | | | Holidays |
| 30 | 9.17 | 6.69 | | | | Holidays |
| 31 | 10.73 | 10.04 | | | | Holidays |
| 32 | 13.75 | 13.38 | | | | Holidays |
| 33 | 17.65 | 17.40 | | | | - Cloth mask mandatory in all schools<br>- Cloth mask mandatory in class for Year 5+ |
| 34 | 16 | 12.72 | | | | |
| 35 | 17.36 | 8.03 | | | | |
| 36 | 17.65 | 8.70 | | | | - Cloth mask not mandatory in class anymore |
| 37 | 28.19 | 24.09 | | | | |
| 38 | 37.36 | 22.08 | | | | |
| 39 | 36.97 | 32.79 | | | | |
| 40 | 43.01 | 28.78 | | | | |
| 41 | 72.57 | 76.29 | | | | |
| 42 | 105.63 | 59.56 | | | | Holidays |
| 43 | 198.59 | 95.70 | | | | Holidays |
| 44 | 237.6 | 154.59 | | | | - Cloth mask mandatory in class for Year 5+<br>- $CO_2$ monitors available |
| 45 | 222 | 170.65 | | | | |
| 46 | 186.1 | 137.19 | | | | |
| 47 | 172.74 | 151.91 | | | | |

| | | | | | | |
|---|---|---|---|---|---|---|
| 48 | 147.48 | 107.07 | | | | |
| 49 | 142.6 | 107.07 | | | | |
| 50 | 147.09 | 107.74 | | | | |
| 51 | 181.91 | 125.81 | | | | - Distance learning Year 8+, no mandatory attendance for lower years |
| 52 | 134.41 | 63.58 | | | | Holidays |
| 53 | 100.07 | 46.18 | | | | Holidays |
| 2021<br>01 | 128.07 | 62.91 | | | | Holidays |
| 02 | 111.97 | 57.55 | | | | School closure |
| 03 | 98.81 | 44.84 | | | | School closure |
| 04 | 85.64 | 41.49 | | | | School closure |
| 05 | 85.15 | 32.12 | | | | School closure |
| 06 | 73.35 | 38.15 | | | | School closure |
| 07 | 72.47 | 38.81 | | | | School closure |
| 08 | 77.25 | 41.49 | | | | - Surgical mask mandatory from Year 9+<br>- Cloth mask mandatory Year 1-8, surgical mask recommended<br>- Masks mandatory in class for all schools |
| 09 | 70.72 | 56.21 | | | | |
| 10 | 108.27 | 91.01 | | | | |
| 11 | 122.51 | 99.04 | | | | |
| 12 | 147.38 | 165.97 | | | | |
| 13 | 155.38 | 131.17 | | | | - Voluntary testing 1x per week for all schools |
| 14 | 173.42 | 139.87 | | | | Holidays |
| 15 | 211.07 | 232.22 | | | | Holidays |
| 16 | 262.28 | 353.35 | | | | - Mandatory testing 2x per week for all schools |
| 17 | 199.95 | 174.67 | | | | - *Bundesnotbremse*: School closure at incidence >165 |
| 18 | 158.86 | | | | | |
| 19 | 110.36 | | | | | |
| 20 | 72.79 | | | | | |
| 21 | 44.98 | | | | | |
| 22 | 28.88 | | | | | |
| 23 | 22.83 | | | | | |
| 24 | 12.69 | | | | | |
| 25 | 9.07 | | | | | - Masks not mandatory outside of school buildings |
| 26 | 12.10 | | | | | |
| 27 | 16.39 | | | | | |

Legend: Green= class in presence, Yellow= Rotating class, Orange = Remote class, Red = school closure (emergency care for children of key workers Year 1-6 and special needs schools), Grey= Holidays. Sources in Supplement References

Table 2: Infection Control Measures for Schools in Region 2

| Calendar Week | Incidence Population per 100.000 | Incidence Students per 100.000 | Primary Schools (PS) | Secondary Schools (SS) | Final Years (FY) | Measures |
|---|---|---|---|---|---|---|
| 26 | 1.85 | 0.00 | | | | Holidays |
| 27 | 2.63 | 0.67 | | | | Holidays |
| 28 | 7.02 | 2.01 | | | | Holidays |
| 29 | 10.92 | 7.36 | | | | Holidays |
| 30 | 9.17 | 6.69 | | | | Holidays |
| 31 | 10.73 | 10.04 | | | | Holidays |
| 32 | 15.73 | 6.33 | | | | - Cloth masks mandatory for SS (not in class) |
| 33 | 10.67 | 12.27 | | | | |
| 34 | 10.39 | 7.91 | | | | |
| 35 | 11.4 | 5.94 | | | | |
| 36 | 15.11 | 9.89 | | | | |
| 37 | 18.03 | 23.74 | | | | |
| 38 | 22.42 | 19.78 | | | | |
| 39 | 28.71 | 30.07 | | | | |
| 40 | 35.9 | 30.86 | | | | |
| 41 | 38.71 | 14.64 | | | | Holidays |
| 42 | 56.69 | 7.12 | | | | Holidays |
| 43 | 110.85 | 66.08 | | | | - Cloth mask mandatory in class for Year 11+ and Vocational schools |
| 44 | 151.47 | 116.72 | | | | |
| 45 | 176.63 | 175.28 | | | | - Cloth mask mandatory in class for SS<br>- Funds provided for $CO_2$ monitors |
| 46 | 155.96 | 155.90 | | | | |
| 47 | 138.71 | 125.82 | | | | |
| 48 | 91.07 | 75.57 | | | | |
| 49 | 125.57 | 83.88 | | | | |
| 50 | 147.53 | 102.87 | | | | |
| 51 | 173.21 | 85.86 | | | | Schools stopped presence classes on 16.12.2020 |
| 52 | 153.99 | 15.83 | | | | Holidays |
| 53 | 145.79 | 3.96 | | | | Holidays |
| 2021 01 | 163.71 | 24.93 | | | | Holidays |
| 02 | 116.3 | 23.74 | | | | School closure |
| 03 | 104.2 | | | | | School closure |
| 04 | 87.67 | | | | | School closure |
| 05 | 74.81 | | | | | School closure |
| 06 | 71.27 | | | | | School closure |
| 07 | 75.94 | | | | | School closure |
| 08 | 85.82 | | | | | School closure |
| 09 | 85.09 | | | | | Holidays |
| 10 | 97.39 | | | | | Holidays |

| | | | | | | |
|---|---|---|---|---|---|---|
| 11 | 123.4 | | | | | - Surgical masks mandatory from age 14+<br>- Voluntary testing for school staff |
| 12 | 161.03 | | | | | - Voluntary testing for students x1 per week and staff x3 per week |
| 13 | 163.55 | | | | | - Voluntary testing for students x2 per week and staff x3 per week |
| 14 | 150.75 | | | | | - Mandatory testing for students x2 per week |
| 15 | 146.26 | | | | | |
| 16 | 121.77 | | | | | |
| 17 | 107.11 | | | | | - Vaccinations available for teachers |
| 18 | 87.84 | | | | | |
| 19 | 1.85 | | | | | Holidays |
| 20 | 2.63 | | | | | |
| 21 | 7.02 | | | | | |
| 22 | 10.92 | | | | | |
| 23 | 9.17 | | | | | |
| 24 | 10.73 | | | | | |
| 25 | 15.73 | | | | | |
| 26 | 10.67 | | | | | Holidays |

Legend: Green= class in presence, Yellow= Rotating class, Orange = Remote class, Red = school closure (emergency care for students independent of their parents' occupation), Grey= Holidays (optional learning holidays in school for all students since W09/21). Sources in Supplement References

Table 3: Infection Control Measures for Schools in Region 3

| Calendar Week | Incidence Population per 100.000 | Incidence Students per 100.000 | Primary Schools (PS) | Secondary Schools (SS) | Final Years (FY) | Measures |
|---|---|---|---|---|---|---|
| 2020 09 | 0.00 | 0.00 | | | | |
| 10 | 0.00 | 0.00 | | | | |
| 11 | 2.03 | 0.00 | | | | |
| 12 | 6.1 | 9.95 | | | | School closure |
| 13 | 12.19 | 9.95 | | | | School closure |
| 14 | 11.18 | 0.00 | | | | School closure |
| 15 | 5.08 | 0.00 | | | | School closure |
| 16 | 5.08 | 9.95 | | | | School closure |
| 17 | 6.1 | 19.91 | | | | School closure |
| 18 | 5.08 | 0.00 | | | 1 | - Cloth mask mandatory in all schools (not in class)<br>- Daily temperature check<br>- 1 – only German Abitur FY in presence |
| 19 | 2.03 | 0.00 | | | | - All FY and special needs schools in presence |
| 20 | 6.1 | 0.00 | 2 | | 2 | 2 – Year 4 and 11 in presence |
| 21 | 5.08 | 0.00 | 2 | | 2 | |
| 22 | 8.13 | 0.00 | 2 | | 2 | |
| 23 | 0.00 | 0.00 | 2 | | 2 | |
| 24 | 2.03 | 0.00 | 2 | | 2 | |
| 25 | 0.00 | 0.00 | | | | |
| 26 | 0.00 | 0.00 | | | | |
| 27 | 1.02 | 0.00 | | | | |
| 28 | 0.00 | 0.00 | | | | |
| 29 | 3.05 | 0.00 | | | | |
| 30 | 0.00 | 0.00 | | | | Holidays |
| 31 | 0.00 | 0.00 | | | | Holidays |
| 32 | 0.00 | 0.00 | | | | Holidays |
| 33 | 0.00 | 0.00 | | | | Holidays |
| 34 | 0.00 | 0.00 | | | | Holidays |
| 35 | 15.24 | 79.63 | | | | Holidays |
| 36 | 5.08 | 29.86 | | | | Phased Plan „Kindertagesbetreuung und Schule unter Pandemiebedingungen für das Kita- und Schuljahr 2020/21 (27.07.2020“)[1] - Phase GREEN |
| 37 | 5.08 | 0.00 | | | | |
| 38 | 1.02 | 0.00 | | | | |
| 39 | 4.06 | 0.00 | | | | |
| 40 | 5.08 | 9.95 | | | | |
| 41 | 11.18 | 39.82 | | | | |
| 42 | 19.3 | 29.86 | | | | |
| 43 | 81.28 | 39.82 | | | | Holidays |

| | | | | | | |
|---|---|---|---|---|---|---|
| 44 | 116.83 | 0.00 | | | | Holidays |
| 45 | 111.75 | 149.31 | | | | |
| 46 | 140.2 | 89.59 | | | | Phase YELLOW[2] for major city in county |
| 47 | 180.84 | 99.54 | | | | |
| 48 | 231.64 | 89.59 | | | | |
| 49 | 379.96 | 228.95 | | | | Phase YELLOW[2] |
| 50 | 395.2 | 278.72 | | | | |
| 51 | 469.37 | 328.49 | | | | |
| 52 | 493.75 | 288.67 | | | | Holidays |
| 53 | 426.7 | 228.95 | | | | Holidays |
| 2021<br>01 | 355.58 | 288.67 | | | | Holidays |
| 02 | 433.81 | 248.86 | | | | School closure ; |
| 03 | 235.7 | 169.22 | | | | School closure |
| 04 | 181.85 | 139.36 | | | | School closure |
| 05 | 140.2 | 79.63 | | | | School closure |
| 06 | 97.53 | 69.68 | | | | School closure |
| 07 | 93.47 | 99.54 | | | | School closure |
| 08 | 88.39 | 109.50 | | | | - Cloth masks mandatory for Year 1-6, not in class<br>- Surgical masks mandatory for Years 7+, not in class |
| 09 | 114.8 | 238.90 | | | | |
| 10 | 160.52 | 179.18 | | | | |
| 11 | 194.05 | 288.67 | | | | |
| 12 | 216.4 | 318.53 | | | | - Vaccinations available for teachers |
| 13 | 224.52 | 179.18 | | | | Holidays |
| 14 | 206.24 | 129.40 | | | | Holidays |
| 15 | 206.24 | 228.95 | | | | - Voluntary testing 2x per week for all schools<br>- Masks mandatory in class |
| 16 | 210.3 | 258.81 | | | | |
| 17 | 197.09 | 238.90 | | | | - *Bundesnotbremse*: School closure at incidence >165, mandatory testing 2x per week for all schools |
| 18 | 214.37 | 328.49 | | | | |
| 19 | 97.53 | 159.27 | | | | |
| 20 | 55.88 | 59.73 | | | | |
| 21 | 39.62 | 69.68 | | | | |
| 22 | 20.32 | 99.54 | | | | - Masks mandatory in class only for Year 7+ |
| 23 | 20.32 | 69.68 | | | | |
| 24 | 8.14 | | | | | |
| 25 | 12.20 | | | | | |
| 26 | 5.08 | | | | | |
| 27 | 13.22 | | | | | - Testing now voluntary 2x per week |
| 28 | 9.15 | | | | | |
| 29 | 7.12 | | | | | |
| 30 | 14.24 | | | | | Holidays |

Legend: Green= class in presence, Yellow= Rotating class, Orange = Remote class, Red = school closure (emergency care for children of key workers Year 1-6 and special needs schools), Grey= Holidays

Table 4: Infection Control Measures for Schools in Region 4

| Calendar Week | Incidence Population per 100.000 | Incidence Students per 100.000 | Primary Schools (PS) | Secondary Schools (SS) | Final Years (FY) | Measures |
|---|---|---|---|---|---|---|
| 2020 09 | 0.00 | 0.00 | | | | |
| 10 | 0.36 | 0.00 | | | | |
| 11 | 10.8 | 0.00 | | | | |
| 12 | 19.07 | 9.95 | | | | School closure |
| 13 | 26.27 | 9.95 | | | | School closure |
| 14 | 54.34 | 0.00 | | | | School closure |
| 15 | 26.63 | 3.55 | | | | School closure |
| 16 | 15.11 | 0.00 | | | | School closure |
| 17 | 10.8 | 0.00 | | | | School closure |
| 18 | 7.2 | 3.55 | | | | |
| 19 | 3.24 | 3.55 | | | | |
| 20 | 2.52 | 0.00 | | | | |
| 21 | 1.08 | 0.00 | 1 | | | 1 – only Year 4 in presence |
| 22 | 1.08 | 0.00 | 1 | | | |
| 23 | 0.36 | 0.00 | | | | Holidays |
| 24 | 0.36 | 0.00 | | | | Holidays |
| 25 | 0 | 0.00 | | | | |
| 26 | 0 | 0.00 | | | | |
| 27 | 0.36 | 0.00 | | | | |
| 28 | 1.08 | 0.00 | | | | |
| 29 | 5.4 | 3.55 | | | | |
| 30 | 9.36 | 7.11 | | | | |
| 31 | 9 | 7.11 | | | | |
| 32 | 4.32 | 3.55 | | | | Holidays |
| 33 | 7.92 | 3.55 | | | | Holidays |
| 34 | 11.16 | 14.22 | | | | Holidays |
| 35 | 9 | 7.11 | | | | Holidays |
| 36 | 8.64 | 0.00 | | | | Holidays |
| 37 | 10.44 | 14.22 | | | | Holidays |
| 38 | 16.55 | 28.44 | | | | |
| 39 | 9 | 7.11 | | | | |
| 40 | 10.44 | 7.11 | | | | |
| 41 | 27.35 | 24.88 | | | | |
| 42 | 31.31 | 0.00 | | | | |
| 43 | 99.68 | 31.99 | | | | - Cloth mask mandatory in all schools<br>- Mask mandatory in class Year 5+ |
| 44 | 129.55 | 28.44 | | | | Holidays |
| 45 | 151.14 | 31.99 | | | | |

| | | | | | | |
|---|---|---|---|---|---|---|
| 46 | 136.03 | 56.87 | | | | |
| 47 | 165.53 | 24.88 | | | | |
| 48 | 93.92 | 24.88 | | | | |
| 49 | 108.32 | 71.09 | | | | |
| 50 | 141.06 | 95.97 | | | | |
| 51 | 171.65 | 53.32 | | | | |
| 52 | 138.18 | 81.75 | | | | Holidays |
| 53 | 129.19 | 49.76 | | | | Holidays |
| 2021 01 | 157.98 | 49.76 | | | | Holidays |
| 02 | 133.51 | 74.65 | | | | School closure |
| 03 | 84.21 | 17.77 | | | | School closure |
| 04 | 66.93 | 39.10 | | | | School closure |
| 05 | 62.61 | 17.77 | | | | School closure |
| 06 | 60.82 | 53.32 | | | | School closure |
| 07 | 85.65 | 46.21 | | | | School closure |
| 08 | 98.96 | 92.42 | | | | |
| 09 | 101.48 | 74.65 | | | | |
| 10 | 127.03 | 85.31 | | | | |
| 11 | 139.26 | 99.53 | | 2 | | 2 – Year 5 and 6 in presence, Year 7+ in distance learning |
| 12 | 147.18 | 181.28 | | 2 | | - Surgical masks mandatory for all schools also in class |
| 13 | 121.99 | 124.41 | | 2 | | |
| 14 | 150.78 | 99.53 | | | | Holidays |
| 15 | 181.01 | 42.65 | | | | - Voluntary testing available |
| 16 | 195.04 | 258.81 | | | | - Mandatory testing for students x2 per week |
| 17 | 130.99 | 238.90 | | | | |
| 18 | 84.91 | | | | | |
| 19 | 65.12 | | | | | |
| 20 | 62.96 | | | | | |
| 21 | 25.18 | | | | | Holidays |
| 22 | 23.75 | | | | | Holidays |
| 23 | 19.07 | | | | | |
| 24 | 11.87 | | | | | |
| 25 | 3.24 | | | | | |
| 26 | 1.8 | | | | | |
| 27 | 2.52 | | | | | |
| 28 | 10.43 | | | | | |
| 29 | 10.07 | | | | | |
| 30 | 10.07 | | | | | |
| 31 | 16.55 | | | | | Holidays |

Legend: Green= class in presence, Yellow= Rotating class, Orange = Remote class, Red = school closure (emergency care for children of key workers Year 1-6 and special needs schools), Grey= Holidays. Sources in Supplement References

Table 5: Infection Control Measures for Schools in Region 5

| Calendar Week | Incidence Population per 100.000 | Incidence Students per 100.000 | Primary Schools (PS) | Secondary Schools (SS) | Final Years (FY) | Measures |
|---|---|---|---|---|---|---|
| 2020 09 | 0.00 | 0.00 | | | | |
| 10 | 1,64 | 0,94 | | | | |
| 11 | 20,69 | 6,60 | | | | |
| 12 | 52,21 | 11,32 | | | | School closure |
| 13 | 48,1 | 7,55 | | | | School closure |
| 14 | 32,18 | 4,72 | | | | School closure |
| 15 | 29,22 | 4,72 | | | | School closure |
| 16 | 16,75 | 1,89 | | | | School closure |
| 17 | 11,82 | 3,77 | | | | School closure |
| 18 | 5,25 | 3,77 | | | | |
| 19 | 6,24 | 5,66 | | | | |
| 20 | 7,06 | 0,00 | | | | |
| 21 | 5,25 | 3,77 | 1 | | | 1 – only Year 4 in presence |
| 22 | 3,45 | 0,00 | 1 | | | |
| 23 | 2,46 | 0,94 | | | | Holidays |
| 24 | 1,48 | 1,89 | | | | Holidays |
| 25 | 3,61 | 4,72 | | | | |
| 26 | 2,3 | 1,89 | | | | |
| 27 | 1,48 | 0,00 | | | | |
| 28 | 4,76 | 3,77 | | | | |
| 29 | 8,21 | 7,55 | | | | |
| 30 | 3,78 | 4,72 | | | | |
| 31 | 3,78 | 1,89 | | | | |
| 32 | 7,06 | 3,77 | | | | Holidays |
| 33 | 8,37 | 1,89 | | | | Holidays |
| 34 | 23,64 | 17,93 | | | | Holidays |
| 35 | 26,76 | 26,42 | | | | Holidays |
| 36 | 24,46 | 26,42 | | | | Holidays |
| 37 | 26,43 | 29,25 | | | | Holidays |
| 38 | 30,86 | 11,32 | | | | |
| 39 | 30,7 | 35,85 | | | | |
| 40 | 34,97 | 25,47 | | | | |
| 41 | 68,95 | 68,88 | | | | |
| 42 | 104,91 | 90,58 | | | | |
| 43 | 134,13 | 109,45 | | | | - Cloth mask mandatory in all schools<br>- Mask mandatory in class Year 5+ |
| 44 | 168,61 | 83,03 | | | | Holidays |
| 45 | 195,86 | 140,58 | | | | |
| 46 | 175,01 | 141,53 | | | | |
| 47 | 192,58 | 142,47 | | | | |
| 48 | 192,08 | 175,49 | | | | |
| 49 | 162,86 | 161,34 | | | | |
| 50 | 178,13 | 170,77 | | | | |
| 51 | 189,62 | 133,03 | | | | |

| | | | | | | |
|---|---|---|---|---|---|---|
| 52 | 119,35 | 77,37 | | | | Holidays |
| 53 | 118,2 | 47,18 | | | | Holidays |
| 2021<br>01 | 122,8 | 45,29 | | | | Holidays |
| 02 | 105,73 | 51,89 | | | | School closure |
| 03 | 90,95 | 47,18 | | | | School closure |
| 04 | 74,7 | 29,25 | | | | School closure |
| 05 | 56,64 | 33,02 | | | | School closure |
| 06 | 36,45 | 17,93 | | | | School closure |
| 07 | 36,28 | 13,21 | | | | School closure |
| 08 | 52,04 | 36,80 | | | | |
| 09 | 64,68 | 55,67 | | | | |
| 10 | 75,36 | 62,27 | | | | - Voluntary testing for staff available |
| 11 | 84,71 | 83,03 | | 2 | | 2 – Year 5 and 6 in presence, Year 7+ in distance learning |
| 12 | 107,21 | 129,26 | | 2 | | - Surgical masks mandatory for all schools also in class |
| 13 | 120,17 | 105,67 | | 2 | | - Voluntary testing available also for students |
| 14 | 122,64 | 101,90 | | | | Holidays |
| 15 | 215,72 | 164,17 | | | | |
| 16 | 222,78 | 188,70 | | | | - Mandatory testing for students x2 per week |
| 17 | 226,89 | 203,80 | | | | |
| 18 | 225,41 | 131,15 | | | | |
| 19 | 121 | | | | | |
| 20 | 86,85 | | | | | |
| 21 | 44,66 | | | | | Holidays |
| 22 | 36,77 | | | | | Holidays |
| 23 | 34,15 | | | | | |
| 24 | 16,09 | | | | | |
| 25 | 9,69 | | | | | - Masks no longer mandatory in class |
| 26 | 7,39 | | | | | |
| 27 | 10,34 | | | | | |
| 28 | 23,15 | | | | | |
| 29 | 20,69 | | | | | |
| 30 | 22,66 | | | | | |
| 31 | 22,98 | | | | | Holidays |

Legend: Green= class in presence, Yellow= Rotating class, Orange = Remote class, Red = school closure (emergency care for children of key workers Year 1-6 and special needs schools), Grey= Holidays. Sources in Supplement References

#### S6 Figures. Incidences of Population, Students and Staff across regions based on local agency data

Figure 1: Incidences of Population, Students and Staff over time in Region 1

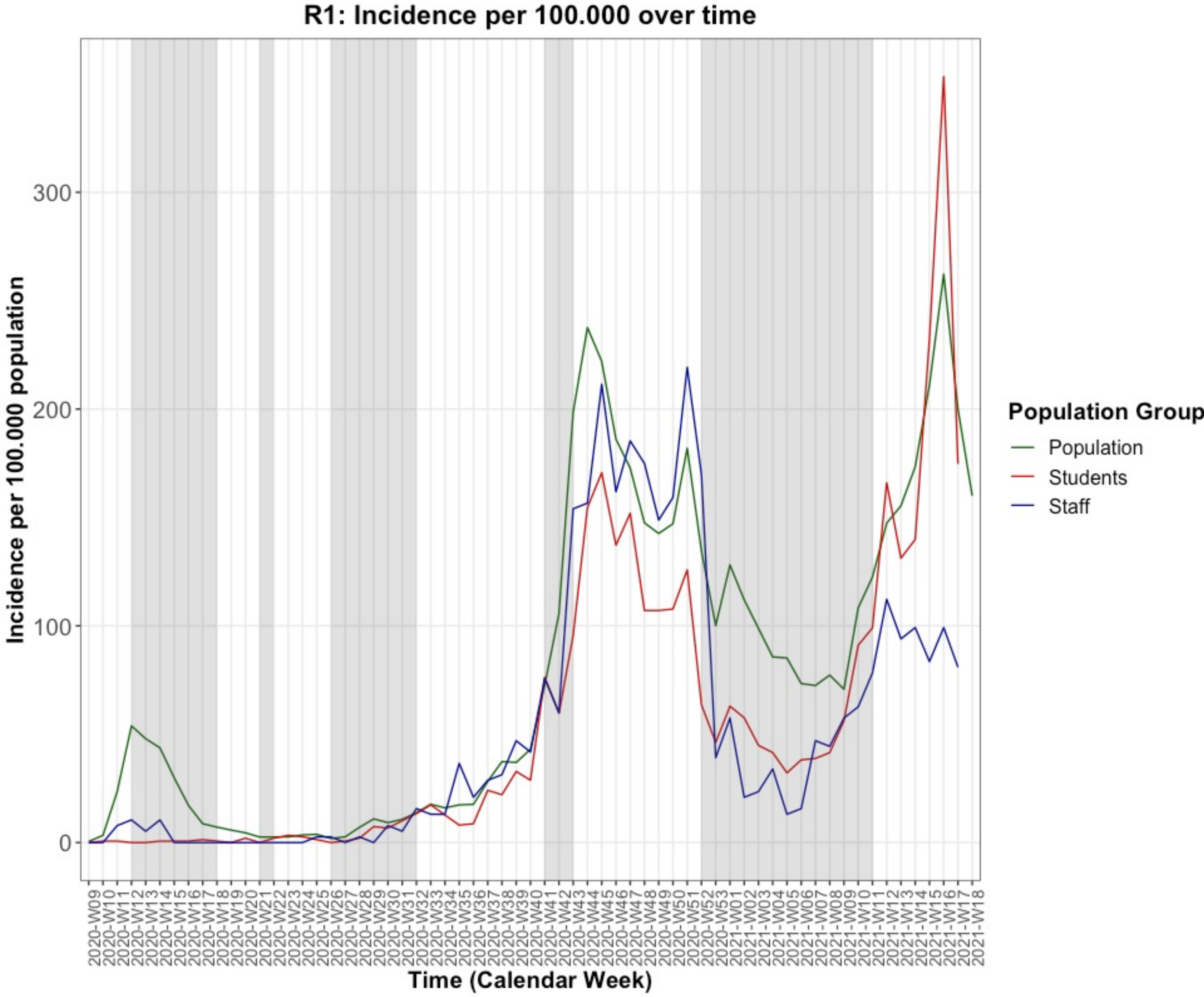

*Grey: areas mark periods of school closure and vacation*
*Population data from RKI (3)*

Figure 2: Incidences of Population, Students and Staff over time in Region 2

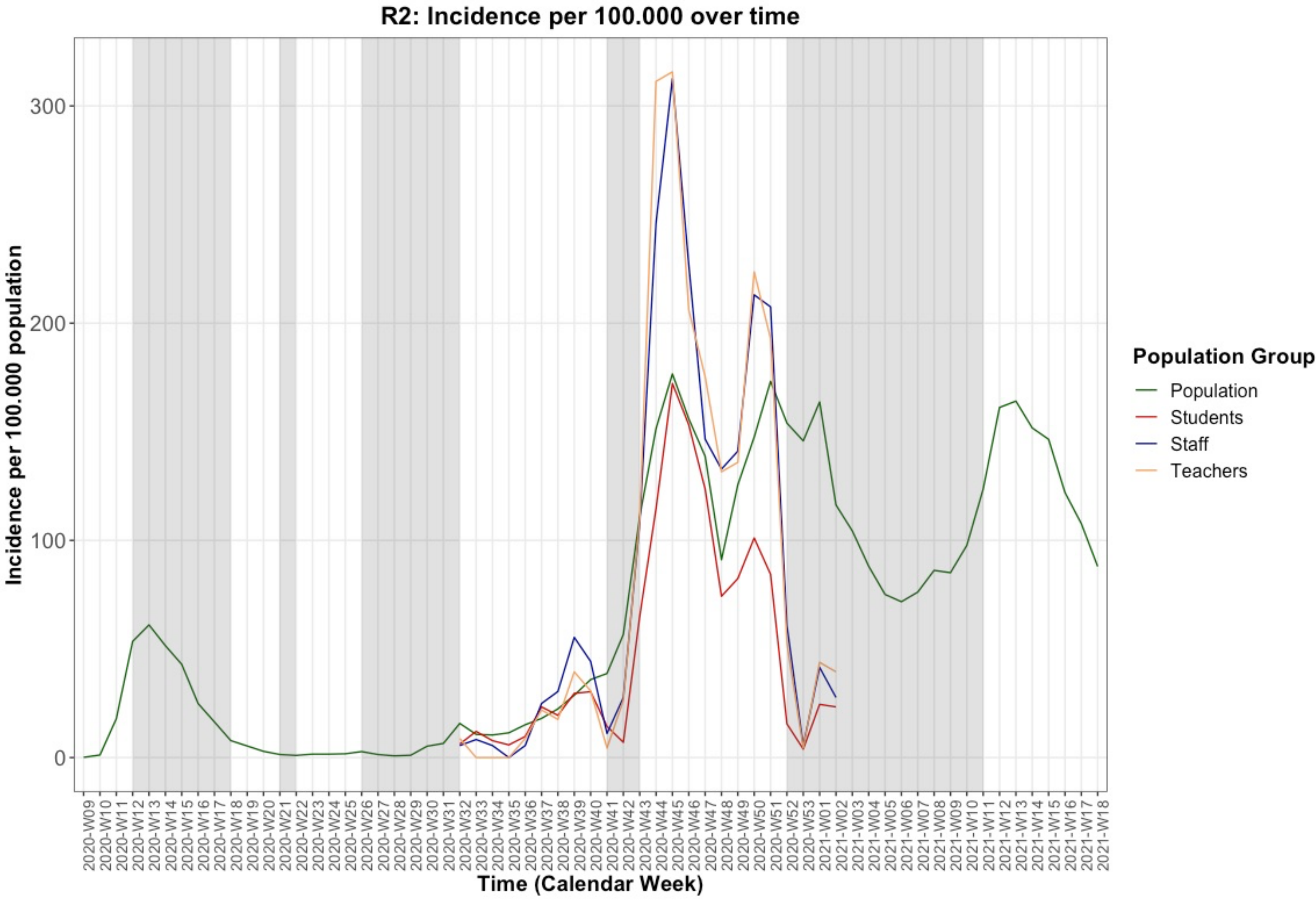

*Grey: areas mark periods of school closure and vacation*
*Population data from RKI (3)*

Figure 3: Incidences of Population and Students over time in Region 3

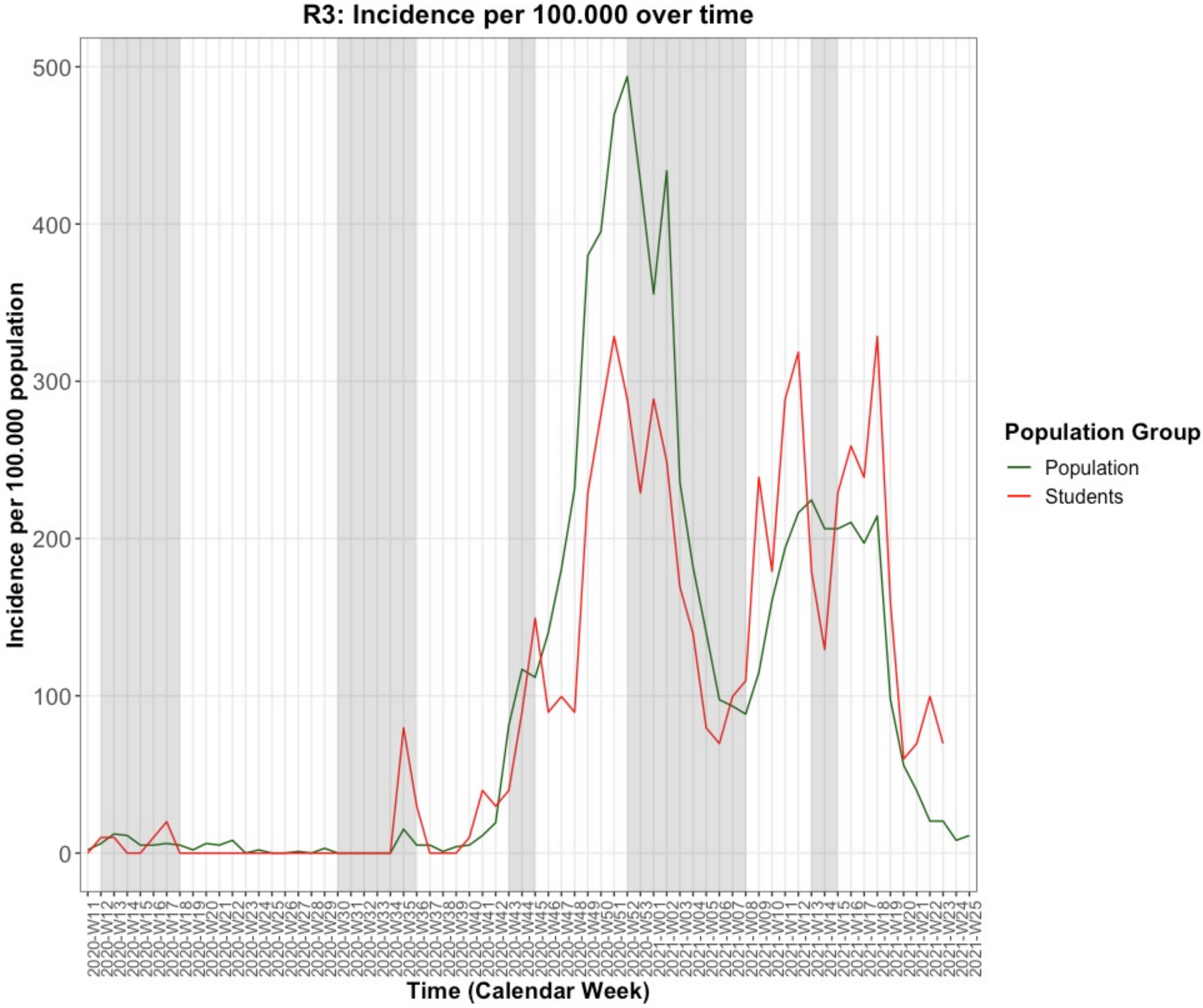

*Grey: areas mark periods of school closure and vacation*
*Population data from RKI (3)*

Figure 4: Incidences of Population and Students over time in Region 4

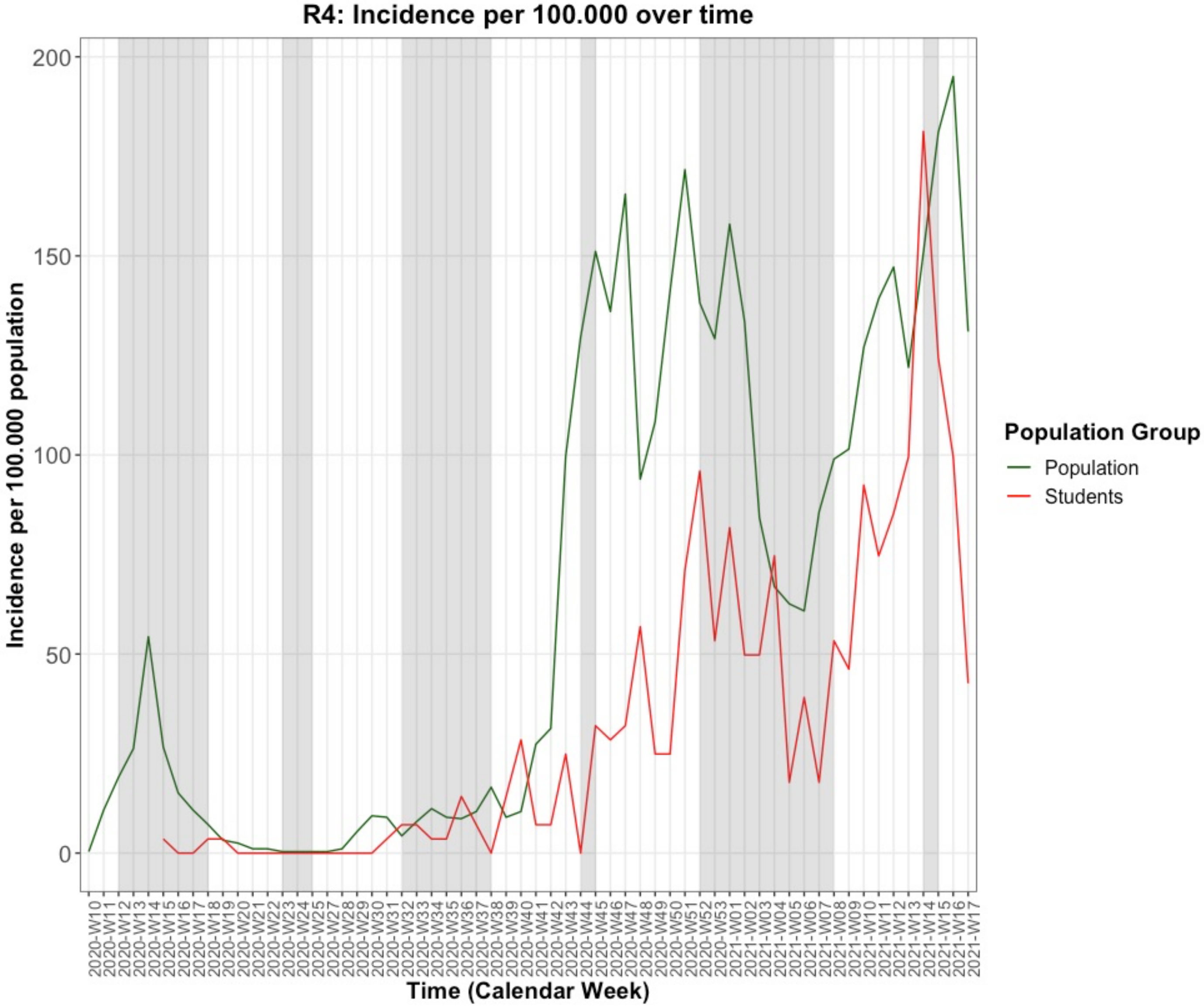

*Grey: areas mark periods of school closure and vacation*
*Population data from RKI (3)*

Figure 5: Incidences of Population and Students over time in Region 5

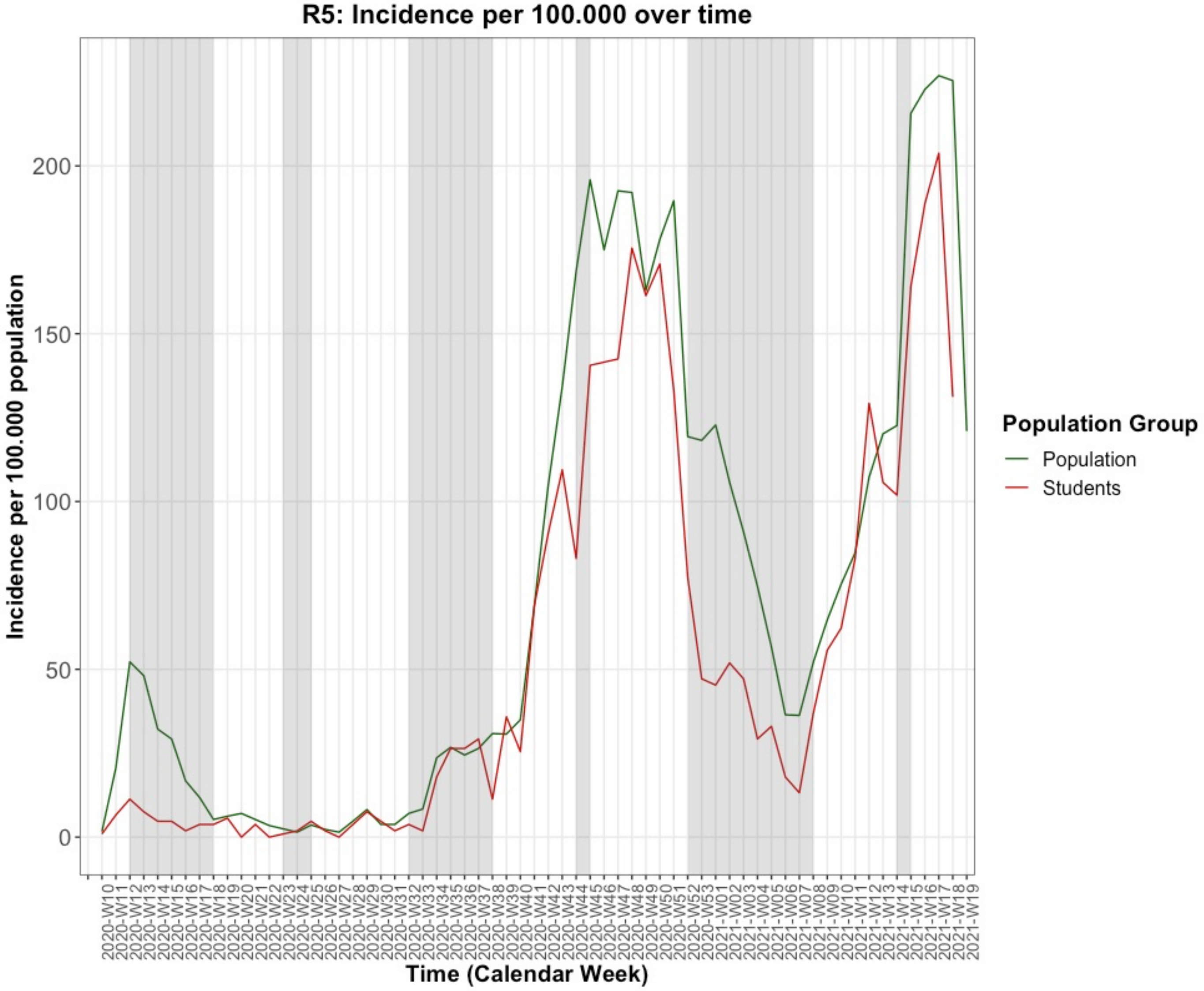

*Grey: areas mark periods of school closure and vacation*
*Population data from RKI (3)*

Figure 6: Incidences of Students (All Regions)

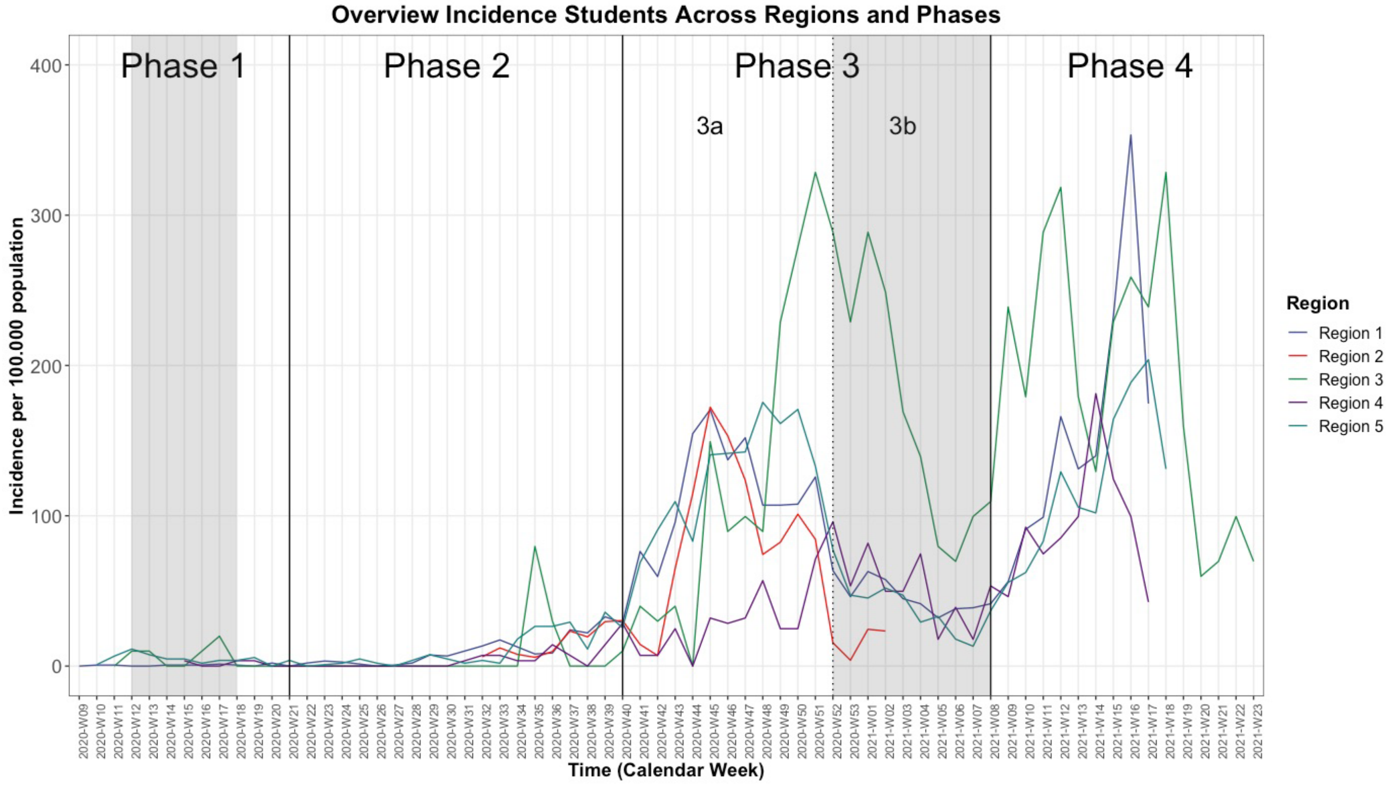

*Grey: Germany-wide school closures*

Figure 7: Incidences of Staff (All Regions)

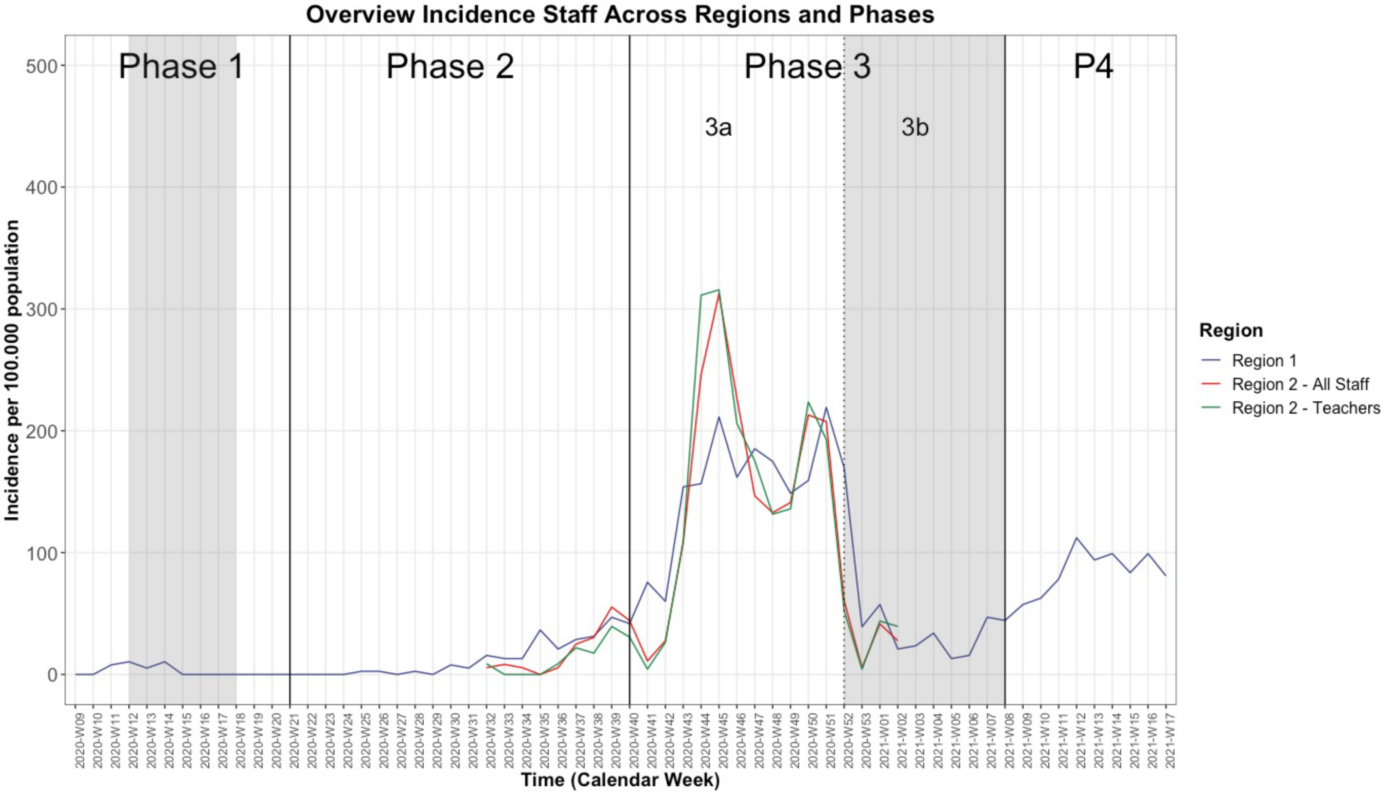

*Grey: Germany-wide school closures*

#### S7 Tables: Infection Risks Across Regions and Subgroups

##### Table 1: Infection Risks, Crude Risk Ratios and Quarantine Risk Across Federal States (KMK Data)

| | Baden-Württemberg | Bavaria | Berlin | Brandenburg | Bremen | Hamburg | Hesse | Mecklenburg Western Pomerania | Lower Saxony | Northrhine-Westphalia | Rhineland Palatinate | Saarland | Saxony | Saxony -Anhalt | Schleswig Holstein | Thuringia |
|---|---|---|---|---|---|---|---|---|---|---|---|---|---|---|---|---|
| W46-50 / 2020 | | | | | | | | | | | | | | | | |
| Population Infection Risk | 0.78% (0.78-0.79%) | 0.96% (0.95-0.96%) | 1.06% (1.06-1.08%) | 0.67% (0.66-0.69%) | 0.67% (0.65-0.69%) | 0.63% (0.62-0.64%) | 0.86% (0.86-0.87%) | 0.28% (0.27-0.29%) | 0.48% (0.48-0.49%) | 0.81% (0.81-0.82%) | 0.72% (0.72-0.73%) | 0.74% (0.73-0.76% | 1.5% (1.49-1.52%) | 0.56% (0.55-0.57%) | 0.28% (0.27-0.28%) | 0.82% (0.81-0.82%) |
| Student Infection Risk | 1.15% (1.13-1.17%) | 1.15% (1.13-1.16%) | 1.65% (1.61-1.69%) | 0.77% (0.74-0.8%) | 1.06% (1-1.13%) | 0.86% (0.83-0.9%) | 0.9% (0.87-0.92%) | 0.08% (0.06-0.09%) | 0.67% (0.65-0.68%) | 1.12% (1.1-1.13%) | 1.06% (1.03-1.09%) | 0.63% (0.59-0.68%) | 0.61% (0.58-0.63%) | 0.57% (0.54-0.6%) | 0.2% (0.19-0.21%) | 0.89% (0.85-0.92%) |
| Student CRR† | 0.2 | 1.16 | 0.15 | 1.14 | 1.58 | 1.36 | 1.04 | 0.29 | 1.39 | 1.38 | 1.46 | 0.84 | 0.41 | 1.02 | 0.72 | 1.08 |
| Teacher Infection Risk | 1.38% (1.32-1.44%) | 1.5% (1.44-1.56%) | 4.78% (4.58-4.98%) | 2.86% (2.66-3.08%) | 1% (0.82-1.21%) | 1.42% (1.27-1.58%) | 1.52% (1.42-1.62%) | 0.22% (0.15-0.31%) | 1.1% (1.03-1.17%) | 2.13% (2.06-2.2%) | 1.33% (1.24-1.44%) | 0.74% (0.59-0.92%) | 2.95% (2.77-3.14%) | 1.82% (1.64-2.02%) | 0.2% (0.16-0.24%) | 4.66% (4.38-4.95%) |
| Teacher CRR | 1.77 | 1.57 | 4.49 | 4.24 | 1.49 | 2.24 | 1.76 | 0.79 | 2.28 | 2.62 | 1.84 | 0.99 | 1.96 | 3.27 | 0.72 | 5.67 |
| Student Quarantine Risk | NA | 16.99% (16.93-17.05%) | 21.89% (21.76-22.02%) | 17.35% (17.22-17.49%) | 18.2% (17.94-18.46%) | 6.36% (6.27-6.46%) | 8.11% (8.05-8.18%) | 9.34% (9.2-9.48%) | NA | 15.33% (15.28-15.38%) | 10.43% (10.35-10.52%) | 9.73% (9.56-9.9%) | 14.64% (14.53-14.74%) | 12.88% (12.75-13.01%) | 0.97% (0.94-0.99%) | 22.17% (22.01-22.34%) |
| Teacher Quarantine Risk | NA | 8.46% (8.33-8.6%) | 9.9% (9.63-10.18%) | 13.53% (13.1-13.96%) | 12.91% (12.28-13.56%) | 3.67% (3.44-3.93%) | 7.17% (6.97-7.38%) | 10.84% (10.32-11.38%) | NA | 11.19% (11.04-11.35%) | 7.82% (7.59-8.06%) | 6.98% (6.51-7.49%) | 5.18% (4.95-5.42%) | 12.81% (12.35-13.29%) | 0.63% (0.56-0.71%) | 37.13% (36.48-37.79%) |
| Phase 4 (W08-23) | | | | | | | | | | | | | | | | |
| Population Infection Risk | 1.69% (1.68-1.7% | 1.65% (1.64-1.65% | 1.44% (1.42-1.45%) | 1.33% (1.31-1.34%) | 1.45% (1.42-1.47%) | 1.43% (1.41-1.44%) | 1.67% (1.66-1.68%) | 1.28% (1.26-1.29%) | 1.25% (1.25-1.26%) | 1.63% (1.62-1.64%) | 1.32% (1.31-1.34%) | 1.33% (1.31-1.35%) | 2.33% (2.31-2.34%) | 1.85% (1.83-1.87%) | 0.78% (0.77-0.79%) | 2.59% (2.57-2.61%) |
| Student Infection Risk | 0.57% (0.56-0.59%) | 1.45% (1.43-1.47%) | 1.83% (1.79-1.87%) | 1.07% (1.03-1.1%) | 1.23% (1.16-1.31%) | 0.92% (0.89-0.96%) | 0.94% (0.92-0.96%) | 0.46% (0.43-0.49%) | 0.79% (0.78-0.81%) | 1.61% (1.59-1.63%) | 2.27% (2.23-2.31%) | 1.51% (1.45-1.59%) | 1.98% (1.93-2.02%) | 2.05% (1.98-2.11%) | 0.17% (0.16-0.19%) | 1.29% (1.25-1.34%) |

| | | | | | | | | | | | | | | | | |
|---|---|---|---|---|---|---|---|---|---|---|---|---|---|---|---|---|
| Student CRR | 0.34 | 0.88 | 1.27 | 0.81 | 0.85 | 0.64 | 0.56 | 0.36 | 0.63 | 0.99 | 1.71 | 1.14 | 0.85 | 1.11 | 0.22 | 0.5 |
| Teacher Infection Risk | 0.66% (0.62-0.71%) | 1.79% (1.73-1.86%) | 2.45% (2.31-2.59%) | 2.9% (2.7-3.12%) | 0.47% (0.36-0.62%) | 0.49% (0.4-0.59%) | 1.06% (0.99-1.15%) | 0.8% (0.66-0.96%) | 0.74% (0.69-0.79%) | 1.82% (1.75-1.89%) | 1.67% (1.56-1.78%) | 0.97% (0.79-1.18%) | 4% (3.78-4.22%) | 3.93% (3.63-4.26%) | 0.12% (0.09-0.15%) | 4.21% (3.95-4.49%) |
| Teacher CRR | 0.39 | 1.09 | 1.71 | 2.19 | 0.32 | 0.34 | 0.63 | 0.63 | 0.59 | 1.12 | 1.26 | 0.73 | 1.72 | 2.13 | 0.15 | 1.63 |
| Student Quarantine Risk | 2.58% (2.55-2.6%) | 6.05% (6.01-6.09%) | 6.03% (5.96-6.11%) | 7.16% (7.06-7.25%) | 11.9% (11.69-12.13%) | 2.86% (2.8-2.93%) | 5.16% (5.11-5.21%) | 5.28% (5.17-5.39%) | NA | 5.91% (5.87-5.94%) | 6.08% (6.02-6.15%) | 5.17% (5.05-5.3%) | 11.51% (11.42-11.61%) | 16.98% (16.81-17.15%) | 2.43% (2.39-2.48%) | 10.23% (10.11-10.35%) |
| Teacher Quarantine Risk | 2.08% (2-2.16%) | 4.47% (4.37-4.57%) | 7.05% (6.82-7.28%) | 9.18% (8.82-9.54%) | 10.9% (10.32-11.51%) | 1.56% (1.4-1.73%) | 6.17% (5.98-6.36%) | 6.38% (5.98-6.81%) | NA | 6.03% (5.91-6.15%) | 5.56% (5.36-5.76%) | 4.87% (4.46-5.32%) | 10.23% (9.9-10.57%) | 19.41% (18.77-20.05%) | 1.82% (1.71-1.94%) | 16.06% (15.57-16.57%) |
| Phase 5 (W24+ [-40]*) | | | | | | | | | | | | | | | | |
| Population Infection Risk | 0.8% (0.8-0.81%) | 0.8% (0.79-0.8%) | 0.87% (0.86-0.88%) | 0.38% (0.37-0.39%) | 0.97% (0.94-0.99%) | 0.89% (0.88-0.91%) | 0.79% (0.78-0.8%) | 0.37% (0.36-0.38%) | 0.57% (0.56-0.57%) | 0.9% (0.88-0.91%) | 0.73% (0.72-0.74%) | 0.69% (0.68-0.71%) | 0.43% (0.42-0.44%) | 0.3% (0.3-0.31%) | 0.47% (0.47-0.48%) | 0.46% (0.45-0.47%) |
| Student Infection Risk | 0.43% (0.42-0.44%) | 0.99% (0.97-1%) | 1.85% (1.81-1.89%) | 0.99% (0.96-1.03%) | 1.33% (1.25-1.41%) | 1.17% (1.13-1.21%) | 0.19% (0.18-0.19%) | 0.45% (0.42-0.49%) | 0.41% (0.4-0.42%) | 3.8% (3.76-3.83%) | 1.83% (1.8-1.87%) | 0.88% (0.83-0.94%) | 0.78% (0.75-0.81%) | 0.83% (0.79-0.87%) | 0.52% (0.5-0.54%) | 0.35% (0.33-0.38%) |
| Student CRR | 0.54 | 1.25 | 2.13 | 2.61 | 1.38 | 1.31 | 0.24 | 1.21 | 0.72 | 4.24 | 2.52 | 1.27 | 1.82 | 2.73 | 1.1 | 0.76 |
| Teacher Infection Risk | 0.18% (0.16-0.2%) | 0.56% (0.53-0.6%) | 1.05% (0.96-1.14%) | 0.87% (0.76-0.99%) | 0.08% (0.04-0.15%) | 0.31% (0.24-0.39%) | 0.07% (0.05-0.1%) | 0.18% (0.12-0.27%) | 0.15% (0.13-0.18%) | 1.07% (1-1.14%) | 0.76% (0.69-0.84%) | 0.35% (0.25-0.49%) | 0.64% (0.55-0.73%) | 0.65% (0.52-0.8%) | 0.07% (0.05-0.1%) | 0.46% (0.38-0.56%) |
| Teacher CRR | 0.22 | 0.71 | 1.21 | 2.29 | 0.08 | 0.35 | 0.09 | 0.48 | 0.27 | 1.2 | 1.04 | 0.51 | 1.49 | 2.14 | 0.15 | 0.99 |
| Student Quarantine Risk | 0.91% (0.9-0.93%) | 2.39% (2.36-2.41%) | 4.55% (4.49-4.61%) | 6.44% (6.35-6.52%) | 3.93% (3.8-4.06%) | 4.05% (3.97-4.12%) | 1.58% (1.55-1.61%) | 2.37% (2.29-2.44%) | NA | 12.44% (12.38-12.5%) | 5.87% (5.8-5.93%) | 2.84% (2.75-2.94%) | 1.79% (1.75-1.83%) | 3.28% (3.2-3.37%) | 1.38% (1.34-1.41%) | 3.17% (3.1-3.23%) |
| Teacher Quarantine Risk | 0.26% (0.23-0.28%) | 0.86% (0.82-0.91%) | 1.08% (0.99-1.17%) | 1.98% (1.81-2.16%) | 3.18% (2.86-3.54%) | 0.17% (0.12-0.24%) | 0.61% (0.55-0.68%) | 0.81% (0.67-0.98%) | NA | 2.51% (2.41-2.62%) | 1.47% (1.37-1.58%) | 0.57% (0.43-0.74%) | 1.11% (1-1.23%) | 0.65% (0.52-0.8%) | 0.19% (0.15-0.23%) | 2.36% (2.17-2.58%) |

*Phase 5 end not yet defined by RKI. W40 represents the latest week of available data at time of submission

†Crude risk ratio of infection

Population Infection Risk: Infection data sourced from RKI survstat (3) and population data sourced from Statistisches Bundesamt (12)

Table 2: Infection Risk Across Regions and Subgroups (Local Agency Regional Data)

| Infection Risk (95% CI) | Phase 2 (W21-39) | | | | | Phase 3a (W40-51) | | | | | Phase 3b (W52-07) | | | | | Phase 4 (W08-23) | | | | |
|---|---|---|---|---|---|---|---|---|---|---|---|---|---|---|---|---|---|---|---|---|
| | R1 | R2* | R3 | R4 | R5 | R1 | R2 | R3 | R4 | R5 | R1 | R2* | R3 | R4 | R5 | R1* | R2 | R3 | R4* | R5 |
| **POPULATION** | | | | | | | | | | | | | | | | | | | | |
| | 0.23% (0.22-0.24%) | 0.11% (0.11-0.12%) | 0.06% (0.04-0.07%) | 0.1% (0.09-0.11%) | 0.21% (0.2-0.22%) | 1.76% (1.73-1.78%) | 1.35% (1.33-1.36%) | 2.39% (2.29-2.49%) | 1.23% (1.19-1.27%) | 1.74% (1.71-1.77%) | 0.84% (0.83-0.86%) | NA | 2.74% (2.63-2.85%) | 0.89% (0.86-0.92%) | 0.74% (0.71-0.76%) | 1.44% (1.42-1.47%) | NA | 2.03% (1.94-2.12%) | 1.22% (1.18-1.26%) | 1.78% (1.75-1.81%) |
| **STUDENTS** | | | | | | | | | | | | | | | | | | | | |
| All Students | 0.18% (0.16-0.2%) | 0.11% (0.1-0.13%) | 0.11% (0.06-0.2%) | 0.08% (0.05-0.11%) | 0.21% (0.18-0.24%) | 1.32% (1.27-1.38%) | 1.02% (0.98%-1.1%) | 1.38% (1.17-1.63%) | 0.37% (0.31-0.44%) | 1.62% (1.54-1.7%) | 0.43% (0.39-0.46%) | NA | 1.61% (1.38-1.88%) | 0.35% (0.29-0.42%) | 0.41% (0.37-0.45%) | 1.53% (1.47-1.6%) | NA | 2.96% (2.64-3.31%) | 0.65% (0.57-0.75%) | 1.74% (1.66-1.83%) |
| Age <10 | 0.11% (0.08-0.15%) | NA | 0.03% (0-0.18%) | 0.08% (0.03-0.17%) | 0.23% (0.17-0.3%) | 0.74% (0.66-0.83%) | NA | 0.72% (0.48-1.06%) | 0.27% (0.18-0.4%) | 1.24% (1.1-1.4%) | 0.31% (0.26-0.38%) | NA | 0.98% (0.69-1.36%) | 0.32% (0.22-0.46%) | 0.36% (0.29-0.46%) | 1.31% (1.2-1.43%) | NA | 2.87% (2.36-3.48%) | 0.87% (0.69-1.08%) | 2.16% (1.97-2.37%) |
| Age 10 to 14 | 0.16% (0.13-0.2%) | NA | 0.09% (0.02-0.26%) | 0.13% (0.08-0.21%) | 0.28% (0.22-0.35%) | 1.08% (0.99-1.18%) | NA | 1.31% (0.98-1.75%) | 0.58% (0.46-0.73%) | 1.97% (1.81-2.14%) | 0.49% (0.43-0.56%) | NA | 1.2% (0.88-1.62%) | 0.29% (0.21-0.4%) | 0.44% (0.37-0.53%) | 1.69% (1.58-1.82%) | NA | 2.65% (2.17-3.24%) | 0.76% (0.62-0.92%) | 2.12% (1.96-2.3%) |
| Age 15 to 19 | 0.25% (0.2-0.3%) | NA | 0.23% (0.1-0.48%) | 0.03% (0.01-0.08%) | 0.28% (0.23-0.35%) | 1.92% (1.8-2.06%) | NA | 2.52% (2.02-3.15%) | 0.27% (0.2-0.38%) | 2.86% (2.67-3.07%) | 0.56% (0.49-0.63%) | NA | 2.82% (2.29-3.47%) | 0.95% (0.79-1.13%) | 0.73% (0.64-0.84%) | 1.82% (1.7-1.95%) | NA | 3.34% (2.76-4.04%) | 0.41% (0.31-0.53%) | 2.58% (2.4-2.78%) |
| Primary school | 0.14% (0.11-0.19%) | 0.05% (0.04-0.07%) | NA | NA | NA | 0.92% (0.82-1.02%) | 0.63% (0.58-0.69%) | NA | NA | NA | 0.35% (0.29-0.41%) | NA | NA | NA | NA | 1.61% (1.49-1.74%) | NA | NA | NA | NA |
| Secondary School | 0.19% (0.16-0.23%) | 0.15% (0.13-0.17%) | NA | NA | NA | 1.69% (1.59-1.79%) | 1.2% (1.14-1.27%) | NA | NA | NA | 0.57% (0.52-0.64%) | NA | NA | NA | NA | 1.86% (1.75-1.97%) | NA | NA | NA | NA |
| Vocational School | 0.19% (0.15-0.24%) | 0.12% (0.1-0.16%) | NA | NA | NA | 1.21% (1.11-1.32%) | 1.16% (1.07-1.25%) | NA | NA | NA | 0.26% (0.22-0.32%) | NA | NA | NA | NA | 0.83% (0.75-0.93%) | NA | NA | NA | NA |
| Special Needs School | 0.2% (0.1-0.38%) | 0.09% (0.03-0.23%) | NA | NA | NA | 1.28% (0.99-1.66%) | 1.36% (1.06-1.74%) | NA | NA | NA | 0.51% (0.34-0.77%) | NA | NA | NA | NA | 1.82% (1.46-2.25%) | NA | NA | NA | NA |
| **STAFF** | | | | | | | | | | | | | | | | | | | | |
| All Staff | 0.23% (0.18-0.28%) | 0.14% (0.1-0.18%) | NA | NA | NA | 1.75% (1.62-1.88%) | 1.82% (1.68-1.96%) | NA | NA | NA | 0.42% (0.36-0.49%) | NA | NA | NA | NA | 0.81% (0.73-0.91%) | NA | NA | NA | NA |
| All Teachers | NA | 0.1% (0.06-0.15%) | NA | NA | NA | NA | 1.86% (1.7-2.05%) | NA | NA | NA | NA | NA | NA | NA | NA | NA | NA | NA | NA | NA |

| | | | | | | | | | | | | | | | | | | | | |
|---|---|---|---|---|---|---|---|---|---|---|---|---|---|---|---|---|---|---|---|---|
| Age <30 | 0.18% (0.01-0.33%) | NA | NA | NA | NA | 2.51% (2.14-2.93%) | NA | NA | NA | NA | 0.41% (0.27-0.61%) | NA | NA | NA | NA | 1% (0.77-1.28%) | NA | NA | NA | NA |
| Age 30 to 34 | 0.36% (0.24-0.55%) | NA | NA | NA | NA | 1.77% (1.46-2.13%) | NA | NA | NA | NA | 0.5% (0.34-0.71%) | NA | NA | NA | NA | 0.74% (0.55-1%) | NA | NA | NA | NA |
| Age 35 to 39 | 0.27% (0.16-0.44%) | NA | NA | NA | NA | 1.47% (1.19-1.82%) | NA | NA | NA | NA | 0.39% (0.25-0.59%) | NA | NA | NA | NA | 0.8% (0.6-1.07%) | NA | NA | NA | NA |
| Age 40 to 44 | 0.23% (0.13-0.41%) | NA | NA | NA | NA | 1.68% (1.36-2.07%) | NA | NA | NA | NA | 0.43% (0.28-0.65%) | NA | NA | NA | NA | 0.89% (0.66-1.19%) | NA | NA | NA | NA |
| Age 45 to 49 | 0.26% (0.15-0.44%) | NA | NA | NA | NA | 1.33% (1.06-1.68%) | NA | NA | NA | NA | 0.31% (0.19-0.51%) | NA | NA | NA | NA | 0.87% (0.65-1.16%) | NA | NA | NA | NA |
| Age 50 to 54 | 0.12% (0.04-0.28%) | NA | NA | NA | NA | 1.76% (1.41-2.21%) | NA | NA | NA | NA | 0.49% (0.32-0.76%) | NA | NA | NA | NA | 0.71% (0.49-1.01%) | NA | NA | NA | NA |
| Age 55 to 59 | 0.07% (0.01-0.21%) | NA | NA | NA | NA | 1.25% (0.96-1.62%) | NA | NA | NA | NA | 0.34% (0.2-0.57%) | NA | NA | NA | NA | 0.52% (0.34-0.79%) | NA | NA | NA | NA |
| Age >60 | 0.11% (0.03-0.29%) | NA | NA | NA | NA | 1.04% (0.76-1.43%) | NA | NA | NA | NA | 0.25% (0.12-0.48%) | NA | NA | NA | NA | 0.36% (0.2-0.62%) | NA | NA | NA | NA |
| Primary school [Teachers] | 0.29% (0.2-0.41%) | [0.13% (0.06-0.26%)] | NA | NA | NA | 2.49% (2.21-2.8%) | [2.01% (1.68-2.39%)] | NA | NA | NA | 0.4% (0.3-0.54%) | NA | NA | NA | NA | 0.99% (0.82-1.2%) | NA | NA | NA | NA |
| Secondary School [Teachers] | 0.17% (0.12-0.25%) | [0.08% (0.04-0.15%)] | NA | NA | NA | 1.32% (1.17-1.49%) | [2.22% (1.97-2.51%)] | NA | NA | NA | 0.4% (0.32-0.5%) | NA | NA | NA | NA | 0.66% (0.55-0.79%) | NA | NA | NA | NA |
| Vocational School [Teachers] | 0.25% (0.14-0.44%) | [0.04% (0-0.23%)] | NA | NA | NA | 1.39% (1.1-1.77%) | [0.84% (0.56-1.27%)] | NA | NA | NA | 0.27% (0.15-0.47%) | NA | NA | NA | NA | 0.64% (0.45-0.92%) | NA | NA | NA | NA |
| Special Needs School [Teachers] | 0.25% (0.12-0.48%) | [0.43% (0.13-1.15%)] | NA | NA | NA | 2.26% (1.82-2.8%) | [2.92% (2-4.23%)] | NA | NA | NA | 0.61% (0.4-0.93%) | NA | NA | NA | NA | 1% (0.72-1.39%) | NA | NA | NA | NA |

*Data for region not available for whole duration of the phase, the infection risk is given for the time available:

- Phase 2: R2 only W33-39
- Phase 3b: R2 only W52-02. Data incomplete due to holiday and school closure periods
- Phase 4: R1 only W08-17; R4 only W08-15

#### S8 Text. Additional Results 1: Infection Risk

##### Student infection risk and age

10-to-14-year-olds show a higher infection risk than under 10-year-olds in phase 3a in R1, R4 and R5, in phase 3b and phase 4 in R1. 10-to-14-year-olds have a lower risk of infection than 15-to-19-year-olds in phase 3a in R1, R3 and R5 (higher in R4) and in phase 3b in R3-5. In phase 4, R5 shows a lower risk than 15-to-19-year-olds and R4 a higher one. 15-to-19-year-olds show a higher infection risk than younger students in phase 3a in R1, R3 and R5 (lower than 10-to-14-year-olds in R4), phase 3b in R3-5 and are only higher than <10-year-olds in R1. For phase 4, the infection risk is either higher than all younger students (R5), just than the under 10-year-olds (R1) or lower than all younger students (R4).

##### Student infection risk and school form

Data was available from R1 and R2 (phases 2 and 3a). Primary schools show a lower risk of infection than secondary schools in phase 2 in R2, phase 3a in R1-2 and phases 3b and 4 in R1. Vocational schools show a lower risk of infection than secondary schools in phases 3 and 4 in R1, as well as a higher risk of infection than primary schools in phase 3a in R2, yet a lower risk in phase 4 in R1.

##### Staff infection risk and school form

Data was available from R1 and R2 (phases 2 and 3a). Infection risk was higher in primary schools than secondary schools in phases 3a and 4 in R1, an effect not seen for teachers in R2 (phase 3a), and higher than in vocational schools in phase 3a in both R1-2. In special needs schools, the risk of infection was higher than secondary and vocational schools in phase 3a in R1. The infection risk of secondary school staff was lower than in primary and special needs schools in phase 3a in R1, an effect not seen for teachers in R2. Among vocational school staff, the risk of infection is lower than in primary and special needs schools in phase 3a in both R1-2, and for teachers also lower than in secondary schools in R2.

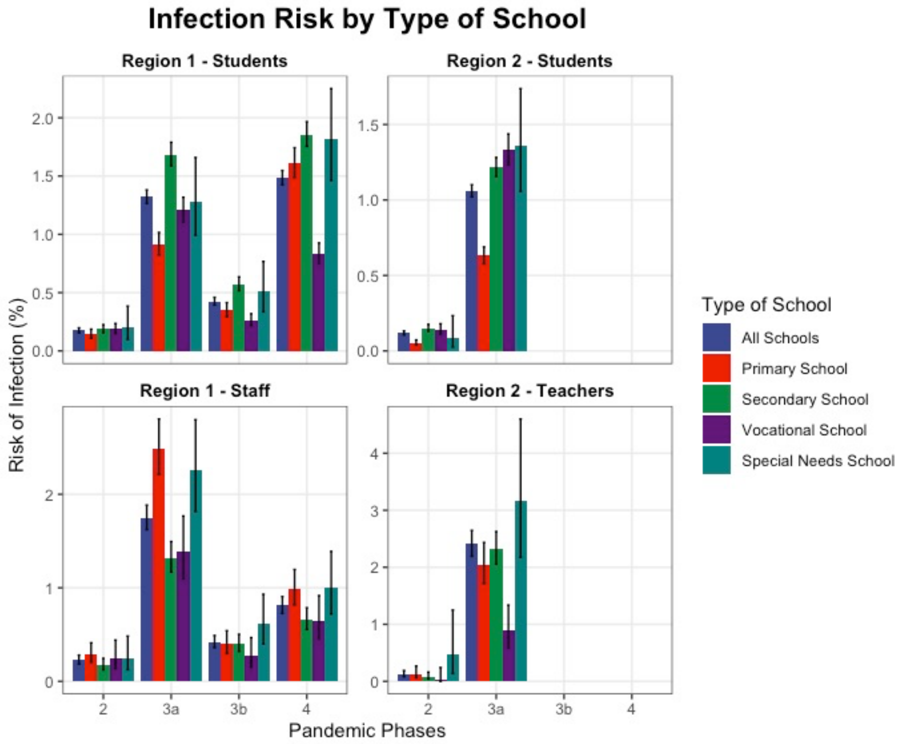

*Figure 1 Infection risks of by school form in two regions*

#### S9 Tables. Secondary Attack Rates

##### Table 1: Secondary Attack Rates in Students by Region, Phase and Age (Local Agency Regional Data)

| | Region 1 | | | | Region 3 | | | | Region 4 | Region 5 | | | |
|---|---|---|---|---|---|---|---|---|---|---|---|---|---|
| **All contact areas** | **Contacts** | | | | **Contacts** | | | | **Contacts** | **Contacts** | | | |
| | Total | Age <10 | Age 10-18 | Age >18 | Total | Age <10 | Age 10-18 | Age >18 | Total | Total | Age <10 | Age 10-18 | Age >18 |
| **All Phases - All Students – All school types. all ages** | | | | | | | | | | | | | |
| Cases | 5108 | | | | 581 | | | | 511 | 3847 | | | |
| Cases with contacts | 2780 | | | | 314 | | | | 213 | 3242 | | | |
| Contacts | 21615 | 5701 | 8200 | 6307 | 2326 | 704 | 824 | 793 | 2256 | 16549 | 4027 | 5348 | 6505 |
| Secondary cases | 1771 | 202 | 372 | 1079 | 172 | 26 | 29 | 117 | 104 | 2118 | 368 | 558 | 1179 |
| Secondary attack rate | 8.2% (7.8-8.6%) | 3.5% (3.1-4.1%) | 4.5% (4.1-5%) | 17.1% (16.2-18.1%) | 7.4% (6.4-8.5%) | 3.7% (2.5-5.4%) | 3.5% (2.4-5%) | 14.8% (12.4-17.4%) | 4.6% (3.8-5.6%) | 12.8% (12.3-13.3%) | 9.1% (8.3-10.1%) | 10.4% (9.6-11.3%) | 18.1% (17.2-19.1%) |
| Secondary cases/ cases | 0.64 | | | | 0.55 | | | | 0.49 | 0.65 | | | |
| **All Phases – Students - Student Age < 10** | | | | | | | | | | | | | |
| Cases | 947 | | | | 162 | | | | 136 | 816 | | | |
| Cases with contacts | 490 | | | | 83 | | | | 66 | 718 | | | |
| Contacts | 5072 | 3677 | 225 | 979 | 862 | 505 | 143 | 214 | 907 | 5098 | 2834 | 481 | 1459 |
| Secondary cases | 303 | 77 | 33 | 172 | 48 | 9 | 5 | 34 | 53 | 535 | 179 | 88 | 257 |
| Secondary attack rate | 6% (5.4-6.7%) | 2.1% (1.7-2.6%) | 14.7% (10.6-19.9%) | 17.6% (15.3-20.1%) | 5.7% (4.2-7.6%) | 8.5% (5.3-13.3%) | 6.8% (2.6-15.2%) | 15.3% (10.9-21.2%) | 5.8% (4.5-7.6%) | 10.5% (9.7-11.4%) | 6.3% (5.5-7.3%) | 18.3% (15.1-22%) | 17.6% (15.7-19.7%) |
| Secondary cases/ cases | 0.62 | | | | 0.58 | | | | 0.80 | 0.75 | | | |
| **All Phases - Students – Student Age 10 to 14** | | | | | | | | | | | | | |
| Cases | 1667 | | | | 185 | | | | 246 | 1280 | | | |
| Cases with contacts | 753 | | | | 78 | | | | 95 | 1104 | | | |
| Contacts | 5701 | 1449 | 2827 | 1169 | 730 | 93 | 401 | 231 | 1047 | 5224 | 732 | 2451 | 1993 |
| Secondary cases | 433 | 64 | 97 | 242 | 42 | 6 | 11 | 25 | 35 | 757 | 126 | 222 | 409 |
| Secondary attack rate | 7.6% (6.9-8.3%) | 4.4% (3.5-5.6%) | 3.4% (2.8-4.2%) | 20.7% (18.5-23.1%) | 5.8% (4.3-7.7%) | 6.5% (2.7-13.6%) | 2.7% (1.5-4.9%) | 10.8% (7.4-15.5%) | 3.3% (2.4-4.6%) | 14.5% (13.6-15.5%) | 17.2% (14.6-20.1%) | 9.1% (8-10.3%) | 20.5% (18.8-22.4%) |
| Secondary cases/ cases | 0.58 | | | | 0.54 | | | | 0.37 | 0.69 | | | |
| **All Phases - Students – Student Age 15 to 19** | | | | | | | | | | | | | |
| Cases | 1929 | | | | 234 | | | | 129 | 1751 | | | |
| Cases with contacts | 1145 | | | | 153 | | | | 52 | 1420 | | | |
| Contacts | 8529 | 319 | 4870 | 2646 | 734 | 106 | 280 | 348 | 302 | 6227 | 461 | 2416 | 3053 |
| Secondary cases | 780 | 50 | 216 | 468 | 82 | 11 | 13 | 58 | 16 | 826 | 63 | 248 | 513 |
| Secondary attack rate | 9.1% (8.6-9.8%) | 15.7% (12.1- | 4.4% (3.9-5.1%) | 17.7% (16.3-19.2%) | 11.2% (9.1-13.7%) | 10.4% (5.7- | 4.6% (2.7-7.9%) | 16.7% (13.1-21%) | 5.3% (3.2-8.5%) | 13.3% (12.4-14.1%) | 13.7% (10.8-17.1%) | 10.3% (9.1-11.5%) | 16.8% (15.5-18.2%) |

| | | | | | | | | | | | | | |
|---|---|---|---|---|---|---|---|---|---|---|---|---|---|
| | | 20.1%) | | | | 17.8%) | | | | | | | |
| Secondary cases/ cases | 0.68 | | | | 0.54 | | | | 0.80 | 0.58 | | | |
| **Phase 2 – W21-39** | | | | | | | | | | | | | |
| Cases | 262 | | | | 11 | | | | 27 | 195 | | | |
| Cases with contacts | 158 | | | | 1 | | | | 17 | 187 | | | |
| Contacts | 2089 | 429 | 913 | 612 | 6 | 2 | 3 | 1 | 195 | 1407 | 123 | 243 | 326 |
| Secondary cases | 85 | 6 | 15 | 50 | 1 | 0 | 1 | 0 | 8 | 165 | 33 | 59 | 71 |
| Secondary attack rate | 4.1% (3.3-5%) | 1.4% (0.6-3.1%) | 1.6% (1-2.7%) | 8.2% (6.2-10.6%) | 16.7% (1.1-58.2%) | 0% (0-71%) | 33.3% (5.6-79.8%) | 0% (0-83.3%) | 4.1% (2-8%) | 11.7% (10.1-13.5%) | 26.8% (19.8-35.3%) | 24.3% (19.3-30.1%) | 21.8% (17.6-26.6%) |
| Secondary cases/ cases | 0.54 | | | | 1 | | | | 0.47 | 0.88 | | | |
| **Phase 2 – Students – Student Age < 10** | | | | | | | | | | | | | |
| Cases | 42 | | | | 1 | | | | 7 | 46 | | | |
| Cases with contacts | 22 | | | | 0 | | | | 5 | 46 | | | |
| Contacts | 365 | 268 | 11 | 63 | 0 | | | | 47 | 467 | 75 | 27 | 82 |
| Secondary cases | 7 | 2 | 1 | 1 | 0 | 0 | 0 | 0 | 3 | 43 | 14 | 9 | 18 |
| Secondary attack rate | 1.9% (0.9-4%) | 0.7% (0-2.9%) | 9.1% (-0.5-39.9%) | 1.6% (-0.5-9.3%) | 0 | 0 | 0 | 0 | 6.4% (1.6-17.8%) | 9.2% (6.9-12.2%) | 18.7% (11.3-29.1%) | 33.3% (18.5-52.3%) | 22% (14.3-32.1%) |
| Secondary cases/ cases | 0.32 | | | | | | | | 0.67 | 0.94 | | | |
| **Phase 2 – Students – Student Age 10 to 14** | | | | | | | | | | | | | |
| Cases | 75 | | | | 3 | | | | 7 | 74 | | | |
| Cases with contacts | 38 | | | | 0 | | | | 5 | 71 | | | |
| Contacts | 567 | 135 | 327 | 86 | 0 | | | | 47 | 422 | 28 | 105 | 117 |
| Secondary cases | 12 | 3 | 2 | 6 | 0 | 0 | 0 | 0 | 3 | 69 | 10 | 34 | 25 |
| Secondary attack rate | 2.1% (1.2-3.7%) | 2.2% (0.5-6.6%) | 0.6% (0-2.4%) | 7% (3-14.7%) | 0 | 0 | 0 | 0 | 2.8% (0.9-7.3%) | 16.4% (13.1-20.2%) | 35.7% (20.6-54.2%) | 32.4% (24.2-41.8%) | 21.4% (14.9-29.7%) |
| Secondary cases/ cases | 0.32 | | | | | | | | 0.6 | 0.97 | | | |
| **Phase 2 – Students – Student Age 15 to 19** | | | | | | | | | | | | | |
| Cases | 104 | | | | 7 | | | | 4 | 75 | | | |
| Cases with contacts | 69 | | | | 1 | | | | 3 | 70 | | | |
| Contacts | 931 | 19 | 539 | 298 | 6 | 2 | 3 | 1 | 7 | 518 | 20 | 111 | 127 |
| Secondary cases | 49 | 1 | 11 | 27 | 1 | 0 | 1 | 0 | 1 | 53 | 9 | 16 | 28 |
| Secondary attack rate | 5.3% (4-6.9%) | 5.3% (-0.9-26.5%) | 2% (1.1-3.7%) | 9.1% (6.3-12.9%) | 16.7% (1.1-58.2%) | 0% (0-71%) | 33.3% (5.6-79.8%) | 0% (0-83.3%) | 14.3% (0.5-53.3%) | 10.2% (7.9-13.2%) | 45% (25.8-65.8%) | 14.4% (9-22.2%) | 22% (15.7-30.1%) |
| Secondary cases/ cases | 0.71 | | | | 1 | | | | 0.33 | 0.76 | | | |
| **Phase 3a – W40-51** | | | | | | | | | | | | | |
| Cases | 1976 | | | | 148 | | | | 133 | 1522 | | | |
| Cases with contacts | 1341 | | | | 69 | | | | 70 | 1326 | | | |
| Contacts | 13644 | 3352 | 6113 | 3466 | 598 | 125 | 263 | 210 | 886 | 8757 | 2189 | 3769 | 2802 |

| | | | | | | | | | | | | | |
|---|---|---|---|---|---|---|---|---|---|---|---|---|---|
| Secondary cases | 667 | 65 | 170 | 410 | 47 | 3 | 10 | 34 | 33 | 796 | 117 | 254 | 418 |
| Secondary attack rate | 4.9% (4.5-5.3%) | 1.9% (1.5-2.5%) | 2.8% (2.4-3.2%) | 11.8% (10.8-12.9%) | 7.9% (5.9-10.3%) | 2.4% (0.5-7.1%) | 3.8% (2-6.9%) | 16.2% (11.8-21.8%) | 3.7% (2.6-5.2%) | 9.1% (8.5-9.7%) | 5.3% (4.5-6.4%) | 6.7% (6-7.6%) | 14.9% (13.6-16.3%) |
| Secondary cases/ cases | 0.5 | | | | 0.68 | | | | 0.47 | 0.6 | | | |
| **Phase 3a – Students – Student Age < 10** | | | | | | | | | | | | | |
| Cases | 286 | | | | 25 | | | | 24 | 250 | | | |
| Cases with contacts | 169 | | | | 11 | | | | 12 | 222 | | | |
| Contacts | 2722 | 2151 | 101 | 427 | 104 | 65 | 12 | 27 | 188 | 2257 | 1474 | 227 | 490 |
| Secondary cases | 83 | 26 | 10 | 46 | 1 | 0 | 0 | 1 | 7 | 163 | 52 | 29 | 77 |
| Secondary attack rate | 3% (2.5-3.8%) | 1.2% (0.8-1.8%) | 9.9% (5.3-17.4%) | 10.8% (8.2-14.1%) | 1% (0-5.8%) | 0% (0-6.7%) | 0% (0-28.2%) | 3.7% (0-19.8%) | 3.7% (1.7-7.6%) | 7.2% (6.2-8.4%) | 3.5% (2.7-4.6%) | 12.8% (9-17.8%) | 15.7% (12.7-19.2%) |
| Secondary cases/ cases | 0.49 | | | | 0.09 | | | | 0.58 | 0.73 | | | |
| **Phase 3a – Students – Student Age 10 to 14** | | | | | | | | | | | | | |
| Cases | 580 | | | | 46 | | | | 74 | 515 | | | |
| Cases with contacts | 353 | | | | 17 | | | | 37 | 467 | | | |
| Contacts | 3653 | 862 | 2132 | 525 | 200 | 24 | 111 | 65 | 503 | 3088 | 450 | 1838 | 886 |
| Secondary cases | 137 | 18 | 38 | 79 | 18 | 1 | 6 | 11 | 16 | 297 | 46 | 97 | 155 |
| Secondary attack rate | 3.8% (3.2-4.4%) | 2.1% (1.3-3.3%) | 1.8% (1.3-2.4%) | 15% (12.2-18.4%) | 9% (5.7-13.8%) | 4.2% (0-21.9%) | 5.4% (2.3-11.5%) | 16.9% (9.5-28%) | 3.2% (1.9-5.1%) | 9.6% (8.6-10.7%) | 10.2% (7.7-13.4%) | 5.3% (4.3-6.4%) | 17.5% (15.1-20.1%) |
| Secondary cases/ cases | 0.39 | | | | 1.06 | | | | 0.43 | 0.64 | | | |
| **Phase 3a – Students – Student Age 15 to 19** | | | | | | | | | | | | | |
| Cases | 814 | | | | 77 | | | | 35 | 757 | | | |
| Cases with contacts | 590 | | | | 41 | | | | 21 | 637 | | | |
| Contacts | 5656 | 162 | 3690 | 1433 | 294 | 36 | 140 | 118 | 195 | 3412 | 265 | 1704 | 1426 |
| Secondary cases | 326 | 16 | 111 | 189 | 28 | 2 | 4 | 22 | 10 | 336 | 19 | 128 | 186 |
| Secondary attack rate | 5.8% (5.2-6.4%) | 9.9% (6.1-15.5%) | 3% (2.5-3.6%) | 13.2% (11.5-15%) | 9.5% (6.6-13.5%) | 5.6% (0.6-19.1%) | 2.9% (0.9-7.4%) | 18.6% (12.6-26.7%) | 5.1% (2.7-9.3%) | 9.8% (8.9-10.9%) | 7.2% (4.6-11%) | 7.5% (6.3-8.9%) | 13% (11.4-14.9%) |
| Secondary cases/ cases | 0.55 | | | | 0.68 | | | | 0.48 | 0.53 | | | |
| **Phase 3b – W52-07** | | | | | | | | | | | | | |
| Cases | 640 | | | | 162 | | | | 122 | 377 | | | |
| Cases with contacts | 185 | | | | 87 | | | | 30 | 295 | | | |
| Contacts | 684 | 224 | 112 | 314 | 247 | 67 | 34 | 146 | 131 | 839 | 102 | 159 | 574 |
| Secondary cases | 106 | 12 | 16 | 75 | 29 | 4 | 1 | 24 | 7 | 163 | 19 | 29 | 115 |
| Secondary attack rate | 15.5% (13-18.4%) | 5.4% (3-9.2%) | 14.3% (8.9-22.1%) | 23.9% (19.5-28.9%) | 11.7% (8.3-16.4%) | 6% (1.9-14.8%) | 2.9% (0-16.2%) | 16.4% (11.2-23.4%) | 5.3% (2.4-10.8%) | 19.4% (16.9-22.2%) | 18.6% (12.2-27.4%) | 18.2% (13-25%) | 20% (17-23.5%) |
| Secondary cases/ cases | 0.57 | | | | 0.33 | | | | 0.23 | 0.55 | | | |

| | | | | | | | | | | | | | |
|---|---|---|---|---|---|---|---|---|---|---|---|---|---|
| **Phase 3b – Students – Student Age < 10** | | | | | | | | | | | | | |
| Cases | 124 | | | | 34 | | | | 28 | 74 | | | |
| Cases with contacts | 38 | | | | 17 | | | | 9 | 60 | | | |
| Contacts | 205 | 132 | 13 | 56 | 57 | 31 | 4 | 22 | 38 | 200 | 47 | 26 | 120 |
| Secondary cases | 9 | 3 | 2 | 3 | 5 | 1 | 0 | 4 | 3 | 42 | 9 | 8 | 25 |
| Secondary attack rate | 4.4% (2.2-8.2%) | 2.3% (0.5-6.8%) | 15.4% (3.1-43.5% ) | 5.4% (1.3-15.2%) | 8.8% (3.4-19.4%) | 3.2% (0-17.6% ) | 0% (0-54.6%) | 18.2% (6.7-39.1%) | 7.9% (2-21.5%) | 21% (15.9-27.2%) | 19.1% (10.2-32.8%) | 30.8% (16.3-50.1%) | 20.8% (14.5-29%) |
| Secondary cases/ cases | 0.24 | | | | 0.29 | | | | 0.33 | 0.7 | | | |
| **Phase 3b – Students – Student Age 10 to 14** | | | | | | | | | | | | | |
| Cases | 226 | | | | 42 | | | | 57 | 114 | | | |
| Cases with contacts | 51 | | | | 16 | | | | 10 | 87 | | | |
| Contacts | 160 | 39 | 44 | 74 | 42 | 4 | 7 | 31 | 39 | 220 | 29 | 47 | 148 |
| Secondary cases | 32 | 4 | 3 | 25 | 7 | 1 | 0 | 6 | 2 | 46 | 7 | 8 | 31 |
| Secondary attack rate | 20% (14.5-26.9%) | 10.3% (3.5-24.2% ) | 6.8% (1.7-18.9% ) | 33.8% (24-45.2%) | 16.7% (8-30.9%) | 25% (3.4-71.1% ) | 0% (0-40.4%) | 19.4% (8.8-36.7%) | 5.1% (0.5-17.8%) | 20.9% (16-26.8%) | 24.1% (12-42.4%) | 17% (8.6-30.4%) | 20.9% (15.1-28.2%) |
| Secondary cases/ cases | 0.63 | | | | 0.44 | | | | 0.2 | 0.53 | | | |
| **Phase 3b – Students – Student Age 15 to 19** | | | | | | | | | | | | | |
| Cases | 286 | | | | 86 | | | | 37 | 189 | | | |
| Cases with contacts | 70 | | | | 54 | | | | 11 | 148 | | | |
| Contacts | 213 | 23 | 45 | 125 | 148 | 32 | 23 | 93 | 54 | 419 | 26 | 86 | 306 |
| Secondary cases | 38 | 3 | 8 | 26 | 17 | 2 | 1 | 14 | 2 | 75 | 3 | 13 | 59 |
| Secondary attack rate | 17.8% (13.3-23.6%) | 13% (3.7-33%) | 17.8% (9-31.6% ) | 20.8% (14.6-28.8%) | 11.5% (7.2-17.7%) | 6.3% (0.7-21.2% ) | 4.3% (0-22.7%) | 15.1% (9.1-23.8%) | 3.7% (0.3-13.3%) | 17.9% (14.5-21.9%) | 11.5% (3.2-29.8%) | 15.1% (8.9-24.3%) | 19.3% (15.2-24.1%) |
| Secondary cases/ cases | 0.54 | | | | 0.32 | | | | 0.18 | 0.51 | | | |
| **Phase 4 – W08-23** | | | | | | | | | | | | | |
| Cases | 2219 | | | | 299 | | | | 225 | 1642 | | | |
| Cases with contacts | 1089 | | | | 151 | | | | 95 | 1334 | | | |
| Contacts | 5163 | 1689 | 1053 | 1902 | 1365 | 497 | 455 | 408 | 1042 | 5018 | 1533 | 946 | 2592 |
| Secondary cases | 911 | 119 | 170 | 543 | 88 | 19 | 17 | 52 | 56 | 952 | 195 | 202 | 551 |
| Secondary attack rate | 17.6% (16.6-18.7%) | 7% (5.9-8.4%) | 16.1% (14-18.5% ) | 28.5% (26.6-30.6%) | 6.4% (5.3-7.9%) | 3.8% (2.4-5.9%) | 3.7% (2.3-5.9%) | 12.7% (9.8-16.4%) | 5.4% (4.2-6.9%) | 19% (17.9-20.1%) | 12.7% (11.1-14.5%) | 21.4% (18.9-24.1%) | 21.3% (19.7-22.9%) |
| Secondary cases/ cases | 0.84 | | | | 0.58 | | | | 0.59 | 0.71 | | | |
| **Phase 4 – Students – Student Age < 10** | | | | | | | | | | | | | |
| Cases | 498 | | | | 100 | | | | 77 | 431 | | | |
| Cases with contacts | 263 | | | | 51 | | | | 40 | 376 | | | |
| Contacts | 1806 | 1147 | 99 | 438 | 597 | 399 | 58 | 140 | 634 | 2080 | 1172 | 194 | 750 |
| Secondary cases | 203 | 46 | 19 | 122 | 37 | 8 | 5 | 24 | 40 | 283 | 104 | 40 | 135 |

| | | | | | | | | | | | | | |
|---|---|---|---|---|---|---|---|---|---|---|---|---|---|
| Secondary attack rate | 11.2% (9.9-12.8%) | 4% (3-5.3%) | 19.2% (12.6-28.1%) | 27.9% (23.9-32.2%) | 6.2% (4.5-8.4%) | 2% (0.9-4%) | 8.6% (3.3-19%) | 17.1% (11.7-24.3%) | 6.3% (4.6-8.5%) | 13.6% (12.2-15.1%) | 8.9% (7.4-10.6%) | 20.6% (15.5-26.9%) | 18% (15.4-20.9%) |
| Secondary cases/ cases | 0.77 | | | | 0.73 | | | | 1 | 0.75 | | | |
| **Phase 4 – Students – Student Age 10 to 14** | | | | | | | | | | | | | |
| Cases | 783 | | | | 93 | | | | 96 | 543 | | | |
| Cases with contacts | 308 | | | | 45 | | | | 39 | 446 | | | |
| Contacts | 1299 | 408 | 317 | 478 | 488 | 65 | 283 | 135 | 364 | 1343 | 214 | 350 | 782 |
| Secondary cases | 252 | 39 | 54 | 132 | 17 | 4 | 5 | 8 | 13 | 326 | 60 | 78 | 187 |
| Secondary attack rate | 19.4% (17.3-21.6%) | 9.6% (7-12.8%) | 17% (13.3-21.6%) | 27.6% (23.8-31.8%) | 3.5% (2.1-5.5%) | 6.2% (2-15.2%) | 1.8% (0.6-4.2%) | 5.9% (2.9-11.4%) | 3.6% (2-6.1%) | 24.3% (22.1-26.6%) | 28% (22.4-34.4%) | 22.3% (18.2-26.9%) | 23.9% (21.1-27%) |
| Secondary cases/ cases | 0.82 | | | | 0.38 | | | | 0.33 | 0.73 | | | |
| **Phase 4 – Students – Student Age 15 to 19** | | | | | | | | | | | | | |
| Cases | 771 | | | | 102 | | | | 52 | 668 | | | |
| Cases with contacts | 415 | | | | 55 | | | | 16 | 512 | | | |
| Contacts | 1725 | 115 | 595 | 788 | 280 | 33 | 114 | 133 | 44 | 1595 | 147 | 402 | 1060 |
| Secondary cases | 367 | 30 | 86 | 226 | 34 | 7 | 7 | 20 | 3 | 343 | 31 | 84 | 229 |
| Secondary attack rate | 21.3% (19.4-23.3%) | 26.1% (18.9-34.8%) | 14.5% (11.8-17.5%) | 28.7% (25.6-31.9%) | 12.1% (8.8-16.5%) | 21.2% (10.4-38%) | 6.1% (2.8-12.3%) | 15% (9.9-22.2%) | 6.8% (1.7-18.9%) | 21.5% (19.6-23.6%) | 21.1% (15.2-28.4%) | 20.9% (17.2-25.1%) | 21.6% (19.2-24.2%) |
| Secondary cases/ cases | 0.88 | | | | 0.62 | | | | 0.19 | 0.67 | | | |

#### S10 Text. Additional Results 2: Secondary Attack Rates

##### Secondary attack rates for students increasing with contact and index age

For all phases, 15-to-19-year-old student index cases had a higher SAR than under <10-year-olds and in several regions than 10-to-14-year-olds. 10-to-14-year-olds show a higher SAR than <10-year-olds in W21-39 (p2) and W40-51 (p3a) in R5, W52-07 (p3b) in R1 and W08-23 (p4) in R1 and R5, and higher than 15-to-19-year-olds only in W21-39 (p2) in R1. The share of index cases that are 15-to-19-year-old is the highest overall and for W21-07 (p2 to 3b), only in W08-23 (p4) is the share of contacts of <10-year-olds highest (Fig. 1).

SARs increased with contact age (S9 Tables). Data was available from R1, R3, R5. For all phases, >18-year-old contacts showed a higher risk of infection than <10-year-olds and also than 10-to-18-year-olds in W21-39 (p2) in R1, W40-51 (p3a) and W08-23 (p4) both in R1 and R3. The share of >18-year-old contacts was 33.6% overall, at 26.8%, 28.2% during W21-51 (p2 and p3a) with open schools and increased to 58.4% and 42.3% during W52-23 (p3b and p4) with part school closures and a higher share of household contacts. For <10-year-old contacts, 64% of index cases were <10-years-old. For 10-to-18 and >18-year-old contacts, most student index cases were 15-to-19 years old, followed by 10-to-14-year-olds, in a similar proportion for both contact age groups (for contact age 10-to-18 45.7% and 39.5% and contact age >18, 30% and 25% respectively).

##### Secondary attack rates of students by school type

Data on contacts and cases by index case school type was available from one region. SARs for the school forms across the observation period showed the lowest risk in primary schools (ps) at 5.8% (95% CI, 5.3-6.4%), lower than the total student SAR of 8.2% (95% CI, 7.8-8.6%), with secondary schools (ss) at 8.2% (95% CI, 7.7-8.7%), and vocational schools (vs) showing the highest SAR at 11.3% (95% CI, 10.3-12.3%).

For staff, for primary schools SAR was 7% (95% CI, 6.1-8%), secondary schools 8.6% (95% CI, 7.4-10%), vocational schools 15.9% (95% CI, 12.4-20.1%) and special needs schools 6.8% (95% CI, 5.2-8.9%); for the phases 2, 3a, 3b and 4 respectively 3.8% (95% CI, 2.5-5.1%), 5.8% (95% CI, 5.1-6.5%), 16% (95% CI, 12.4-20.5%) and 22.4% (95% CI, 19.5-25.7%).

Comparing age groups, >30-year-old staff show a lower SAR in the school setting than the age group from 45-49, 50-54 years, with 0.6% (95% CI, 0.2-1.4) compared to 3.1% (95% CI, 1.7-5.4%) and 3.48% (95% CI, 1.6-7.2%).

Special needs school (sns) showed a higher SAR than primary schools and the average at 10.6% (95% CI, 8.7-12.8%).
Whereas vocational schools have a higher SAR for contacts in schools at 2.1% (95% CI, 1.5-2.8%) than primary schools (0.9%; 95% CI 0.7-1.2%) and the average (1.2%; 95% CI, 1.1-1.4%), special needs schools have a higher SAR in the household (5.4%; 95% CI, 29.5-41.8%) than all other school forms.
Vocational schools show the highest SAR overall in W21-51 (p2 and p3a – except for special needs schools) with open schools, with primary schools showing the lowest in W40-51 (p3a). Special needs schools showed a higher household SAR (35.4%; 29.5-41.8%) than the average (23.2%; 22.2-24.2%), secondary schools (21.8%; 20.5-23.3%) and vocational schools (24.6%; 22.5-26.8%). During school closures in phase 3b, all schools, except primary schools due to a

significant proportion of emergency care in schools, show their highest SAR yet (ss: 25.7%; 20.4-31.7%, vs: 23.3%; 16.8-31.3%, sns: 60%; 22.9-88.4%) as the proportion of household contacts and household secondary cases rises. Further, household SAR in W08-23 (p4) is highest in primary schools compared to other phases (27.5%; 24.4-30.9) and secondary and vocational household SAR are similar to school closures (p3b) (26.4%; 24.1-28.7% and 33.5%; 29.6-27.7%) despite mixed class models and teaching in presence.
Among household contacts across all school forms the majority are over 18 years old (ps: 58%, ws: 60%, vs: 77%, fs: 56%), whereas school contact proportion change with student index age (S9 Tables).

Staff SAR

Comparing different school setting, overall staff SAR differences are based on the relative proportion of school and household cases in the relative school settings and observation intervals. This is evident in the jump in overall SAR from W21-51 (p2 and p3a) to W52-23 (p3b and p4), with W21-39 (p2) and W40-51 (p3a) showing a majority of school contacts (70.5% and 62.9%) and W52-07 (p3b) (with only one secondary case reported for school staff during school closures) and W08-34 (p4) a majority of household contacts (64.2%; 53.8% with 26.4% school and remaining “other”).

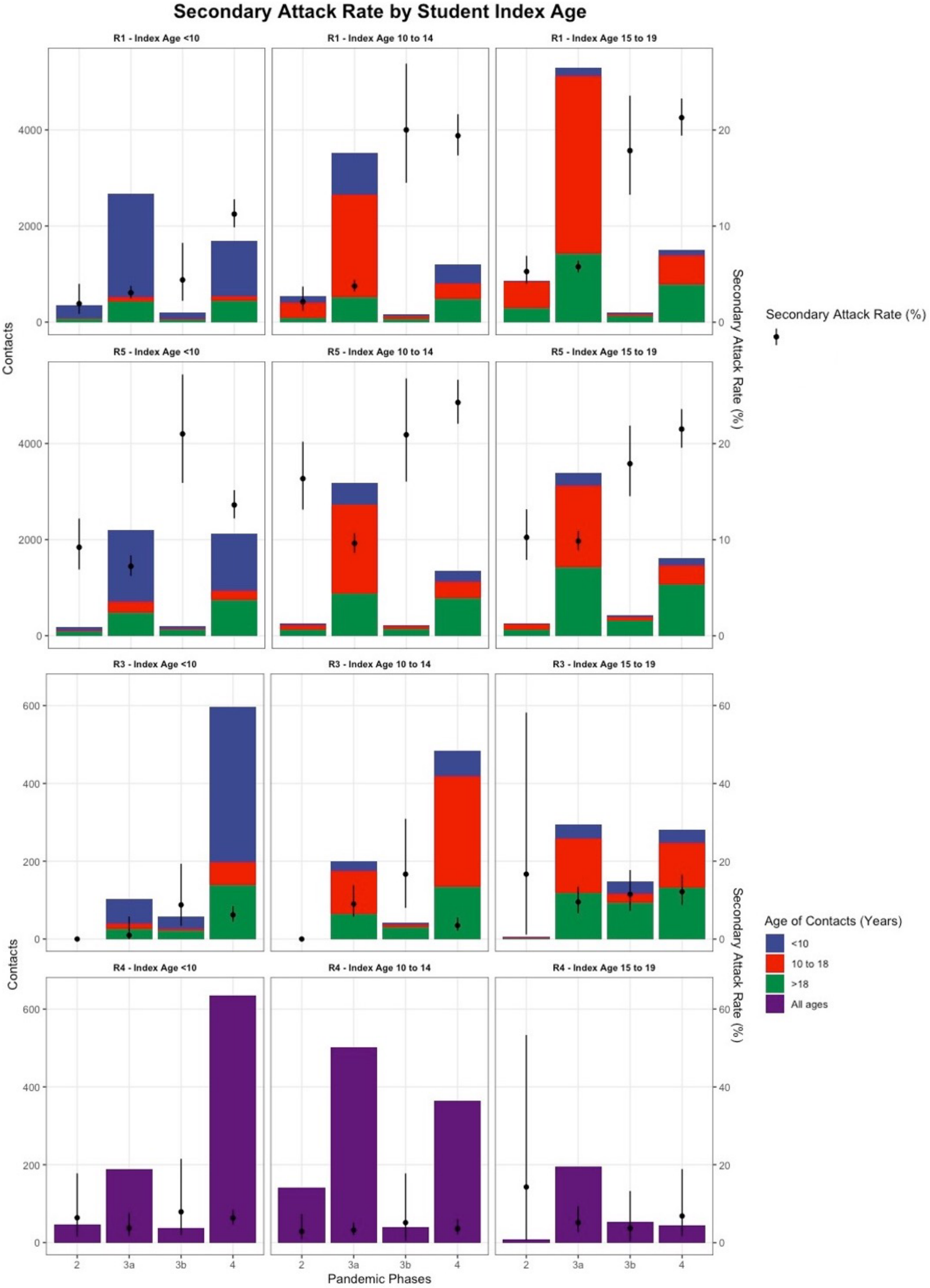

*Figure 1 Risk of infection after contact with an infected student by age of the infected student and overall distribution of number of contacts*

#### S11 Text. Additional Results 3: Effects of NPIs in Schools

Table 2 (Main Text) shows the result for our first regression approach that investigates the impact of school-specific NPIs on infection activities among teachers and students. For the students, we run an OLS model of the official active cases per 100k students as reported by the KMK on the two-week-lags of the earlier estimated attendance rates different types of school, different degrees of mask duties and testing while controlling for the two-week incidence per 100k inhabitants of the corresponding county, the percentage of fully vaccinated persons in the corresponding federal state, alongside the socioeconomic status and geography of the county. For the teachers, we run a similar model, yet not testing the school-type specific but the overall attendance rate as an explanatory variables since earlier tests did give plausible results and we may assume that the children's age does not influence the infection risk of the teachers strongly given the current hygiene measures.

Whereas higher attendance rates among students are associated with higher c.p. infection risks of teachers, the effect appears to depend very much on the childrens' age for the students. On average, c.p., a one percentage point (p.p.) higher attendance in school is associated with 0.22 more cases per 100000 (or 2.2 per 1 million) teachers per week. Among students, higher attendance rates are associated with lower incidence rates among primary school children, whereas the effects for secondary school children and those in graduate classes are positive. For instance, a 1 p.p. higher attendance among secondary school children is associated with on average, c.p., 5.5 more cases per 1 million students.
Testing in schools significantly affects the infections notified to authorities among students and teachers. For instance, controlling for the other factors included in the model, the offer to subsidized testing of school employees in our study period was associated with an average 1.4 p.p. increase in the case numbers among the teachers, which was further amplified by testing among the students and by making tests mandatory instead of voluntary.
Assessment of air filters and $CO_2$ sensors did not yield statistically significant results.
The level of vaccination coverage among the population affects case numbers among students and teachers highly significantly. For instance, a 1 p.p. higher share of fully vaccinated persons is associated with a decrease in the weekly incidence per 100000 of 4.1 for students and 5.5 for teachers.
A lower socioeconomic status, represented by a higher deprivation index, is associated with c.p. higher infection risks among students and teachers, with a stronger effect on the teachers.
The impact of geographical factors on infection risks is inverted between students and teachers. Higher urbanity is associated with higher c.p. infection rates among students when controlling for deprivation. For teachers, however, there are only statistically significant differences in c.p. infection risks for metropolises and regiopolises in comparison to small villages in rural areas. For instance, according to our estimates, we would, c.p., expect 15 per 100000 more cases among students in a metropolis in comparison to a village in a rural area, whereas for the teachers, we would expect 60 per 100000 cases less under the same circumstances.

##### Relative model

Attendance rates in schools appear to not affect infections among teachers to a different degree than the average population. However, there appear to be differences between the students and the population as well as among different groups of students. Whereas the model could not detect a different sensitivity of infection rates in primary school students and

the population towards school attendance, secondary school children appear to respond slightly weaker than the total population, graduate students, however, appear to overproportionately pick up infections at higher physical presence in school. Subsidized testing of school employees is associated with overproportionately many cases in students in teachers in comparison to the total population. However, this difference relativizes somewhat for mandatory instead of voluntary testing.
Supplementary Results 3, Table 1 shows the results of the investigation on how the school-related NPIs affect infection activity in schools relative to the overall population, represented by the 14-day incidence.

*Table 1. Regression Results for Ratio of Active Cases to 14-day incidences for Students or Teachers relative to the overall Population, respectively, on the two-week-lags of school-specific NPIs*

| **Variable** | **Students (95% CI)** | **Teachers (95% CI)** |
|---|---|---|
| 2-week incidence per 100k inhabitants | - 0.0003 (- 0.0004; - 0.0002) | - 0.0004 (- 0.0008; - 0.0001) |
| Attendance in schools | | |
| Open schools | baseline | baseline |
| School vacation | 0.09 (0.03; 0.2) | - 0.1 (- 0.3; 0.1) |
| School closures | 0.01 (- 0.1; 0.1) | - 0.1 (- 0.3; 0.1) |
| Reduced presence in schools | 0.12 (0.05; 0.18) | – 0.1 (- 0.2; - 0.1) |
| Mask mandates | | |
| No mask mandate or voluntary masking | baseline | baseline |
| Partial mask mandate in some or all schools | - 0.11 (- 0.16; - 0.06) | 0.05 (- 0.06; 0.16) |
| Mandatory masks in all school classes | - 0.3 (- 0.35; - 0.25) | - 0.24 (- 0.36; - 0.13) |
| Testing | | |
| No testing in schools | baseline | baseline |
| Voluntary testing in schools | 0.06 (0.01; 0.1) | 0.03 (-0.1; 0.1) |
| Mandatory testing in schools | - 0.01 (- 0.07; 0.04) | - 0.1 (- 0.2; 0.04) |
| Percentage of completely vaccinated population in corresponding federal state [cont.] | 0.02 (0.01; 0.03) | - 0.02 (- 0.03; - 0.003) |
| Urbanity† | | |
| Rural | baseline | baseline |
| Urban (RegioStar71/72) | 0.09 (0.03; 0.15) | - 0.19 (- 0.31; - 0.06) |
| Deprivation Index * | | |
| Deprivation Index 0-0.5 | baseline | baseline |
| Deprivation Index 0.51-1 | -0.01 (- 0.07; 0.04) | 0.42 (0.31; 0.54) |
| $R^2$ [as %] (sample size) | 16.6 (3809) | 6.04 (3979) |

Attendance in schools: Open Schools = 81-100% of students in presence, School vacation = 0%, School closures 1-20%, Reduced presence in schools = 21-80%
Partial mask mandate in some or all schools: mask mandates only for secondary schools, for all schools but not in class, or in class only for secondary schools
*Deprivation Index: RKI German Index of Social Deprivation (9,10)
†Urbanity Index by Federal Ministry of Transportation and Infrastructure; 71= metropolis, urban centre defined by infrastructure and service availability in the region; 72 = regiopolis, regional urban centre defined by infrastructure and service availability in region (11)

Sensitivity Analysis

S11 Text, Table 2 and 3 show the results of a sensitivity analysis splitting the observation period into rising infections during the 4th wave (W08-16) and waning infections during the 4th wave (W17-23).

S11 Text, Table 4 and 5 show previous analyses using the variable coding in Table 6. The original (full) model is presented next to the final model. The original selection of variables yielded some insensible and insignificant estimates due to multicollinearity (Table 7), thus some variables were excluded or rearranged for further analysis in the final model and the new variable grouping as per S3 Text, Table 1.

*Table 2: Sensitivity Analysis by background population infection trends in students*

| Variable | Students W08-16 Estimate | Students W17-23 Estimate |
|---|---|---|
| 2-week incidence per 100k inhabitants | - 0.1 (- 0.8; - 1.6) | 0.28 (0.25; 0.3) |
| Attendance in schools | | |
| Open schools | baseline | baseline |
| School vacation | 29.4 (- 25.5; 84.3) | - 40.1 (- 49.8; - 30.3) |
| School closures | - 23.8 (- 79.7; 32) | - 85.1 (- 98; - 72.2) |
| Reduced presence in schools | 6.4 (- 48.3; 61.2) | - 33.8 (- 43.4; - 24.2) |
| Mask mandates | | |
| No mask mandate or voluntary masking | baseline | baseline |
| Partial mask mandate in some or all schools | - 22.8 (- 33.9; - 11.6) | - 24.4 (- 35.4; - 13.5) |
| Mandatory masks in all school classes | - 47.8 (- 59.4; - 36.2) | - 69.7 (- 81.2; - 58.2) |
| Testing | | |
| No testing in schools | baseline | baseline |
| Voluntary testing in schools | 7.6 (- 4.1; 19.2) | 67.7 (32.6; 102.9) |
| Mandatory testing in schools | 25 (- 7.8; 57.8) | 46.6 (11.6; 81.7) |
| Percentage of completely vaccinated population in corresponding federal state [cont.] | - 2.8 (- 5.7; -0.0009) | - 5.6 (- 6.5; - 4.6) |
| Urbanity† | | |
| Rural | baseline | baseline |
| Urban (RegioStar71/72) | 1 (- 14.1; 16.1) | 31.8 (20.3; 43.4) |
| Deprivation Index * | | |
| Deprivation Index 0-0.5 | baseline | baseline |
| Deprivation Index 0.51-1 | 23 (9.7; 36.3) | -15.9 (- 26.1; - 5.6) |
| $R^2$ [as %] (sample size) | 0.2 (1393) | 0.5 (2146) |

Attendance in schools: Open Schools = 81-100% of students in presence, School vacation = 0%, School closures 1-20%, Reduced presence in schools = 21-80%

Partial mask mandate in some or all schools: mask mandates only for secondary schools, for all schools but not in class, or in class only for secondary schools

*Deprivation Index: RKI German Index of Social Deprivation (9,10)

†Urbanity Index by Federal Ministry of Transportation and Infrastructure; 71= metropolis, urban centre defined by infrastructure and service availability in the region; 72 = regiopolis, regional urban centre defined by infrastructure and service availability in region (11)

*Table 3: Sensitivity Analysis by background population infection trends in teachers*

| **Variable** | **Teachers W08-16 Estimate** | **Teachers W17-23 Estimate** |
|---|---|---|
| 2-week incidence per 100k inhabitants | 0.4 (0.3; 0.5) | 0.6 (0.5; 0.63) |
| Attendance in schools | | |
| Open schools | baseline | baseline |
| School vacation | -155 (- 297.9; - 12.2) | - 21.2 (- 39.3; - 3.3) |
| School closures | - 159.7 (- 304.3; - 15.2) | - 75.1 (- 98.7; - 51.5) |
| Reduced presence in schools | - 134.7 (- 276.7; 7.3) | - 22.9 (- 40.4; - 5.4) |
| Mask mandates | | |
| No mask mandate or voluntary masking | baseline | baseline |
| Partial mask mandate in some or all schools | - 22.8 (- 48.9; 3.3) | 1.2 (- 15.5; 17.9) |
| Mandatory masks in all school classes | - 106.6 (- 135.9; - 77.3) | - 54.2 (- 72; - 36.4) |
| Testing | | |
| No testing in schools | baseline | baseline |
| Voluntary testing in schools | 20.2 (- 5.9; 46.4) | 26.9 (- 35.4; 89.3) |
| Mandatory testing in schools | 38.6 (- 45.4; 122.5) | 16.4 (- 45.8; 78.5) |
| Percentage of completely vaccinated population in corresponding federal state [cont.] | - 4.4 (- 11.3; 2.5) | - 1.4 (- 3.1; 0.3) |
| Urbanity† | | |
| Rural | baseline | baseline |
| Urban (RegioStar71/72) | - 91.4 (- 125; - 57.9) | - 21.1 (- 38.7; - 3.5) |
| Deprivation Index * | | |
| Deprivation Index 0-0.5 | baseline | baseline |
| Deprivation Index 0.51-1 | 149.6 (119.8; 179.4) | 36.4 (20.8; 51.2) |
| $R^2$ [as %] (sample size) | 0.3 (1557) | 0.4 (2152) |

Attendance in schools: Open Schools = 81-100% of students in presence, School vacation = 0%, School closures 1-20%, Reduced presence in schools = 21-80%

Partial mask mandate in some or all schools: mask mandates only for secondary schools, for all schools but not in class, or in class only for secondary schools

*Deprivation Index: RKI German Index of Social Deprivation (9,10)

†Urbanity Index by Federal Ministry of Transportation and Infrastructure; 71= metropolis, urban centre defined by infrastructure and service availability in the region; 72 = regiopolis, regional urban centre defined by infrastructure and service availability in region (11)

*Table 4: Regression Results for Active Cases per 100k Students or Teachers, respectively, on the two-week-lags of school-specific NPIs*

| **Variable** | **Students Estimate of change in notified infections / 100.000 persons/14 days (95%-CI)** | | **Teacher Estimate of change in notified infections / 100.000 persons/14 days (95%-CI)** | |
|---|---|---|---|---|
| | **Final Model** | **Full Modell** | **Final Modell** | **Full Model** |
| 2-week incidence per 100k inhabitants | 0.29 (0.27; 0.3) | 0.27 (0.26; 0.3) | 0.52 (0.49; 0.56) | 0.51 (0.48; 0.55) |
| Attendance rate in schools [as %] | - | - | 0.2 (0.05; 0.4) | 0.2 (0.03; 0.4) |
| Attendance rate in primary schools [as %] | - 0.3 (- 0.4; - 0.2) | - 0.3 (- 0.4; - 0.2) | - | - |
| Attendance rate in secondary schools [as %] | 0.6 (0.4; 0.7) | 0.6 (0.5; 0.8) | - | - |
| Attendance rate in graduation classes [as %] | 0.1 (0.02; 0.2) | 0.05 (- 0.04; 0.15) | - | - |
| Mandatory protective face covering *for all* types of schools [bin.] | - 43.6 (- 48.4; - 38.8) | - 44.7 (- 49.7; - 39.8) | - 45.5 (- 54.5; - 36.6) | - 46.5 (- 55.6; - 37.4) |
| Mandatory protective face covering in primary school classes [bin.] | - | 8.5 (2.8; 14.1) | - 20 (- 30.4; - 9.7) | - 19.2 (- 29.6; - 8.8) |
| Mandatory protective face covering in secondary school classes [bin.] | - | 21.1 (13.6; 28.6) | - 82.2 (- 95.3; - 69.2) | - 85.5 (- 99.4; - 71.7) |
| Voluntary testing of employees in schools [bin.] | 78.5 (63.7; 93.2) | 76.3 (61.6; 91) | 140.6 (114.1; 167) | 139.9 (112.7; 167) |
| Voluntary testing of students and employees in schools [bin.] | 41.3 (33.5; 49) | 38.2 (30.4; 46) | 12.8 (1; 24.6) | 11.2 (- 2.5; 24.8) |
| Mandatory testing of employees and secondary school students [bin.] | 109.7 (83.5; 135.8) | 129.9 (103.4; 156.5) | 49.7 (1; 98.4) | 45.6 (- 3.8; 95) |
| Mandatory testing of employees and students for all schools [bin.] | 18.9 (9.3; 28.5) | 15.3 (5.8; 24.9) | - | - 5.1 (- 21.1; 10.9) |
| Percentage of completely vaccinated population in corresponding federal state [as %] | - 2.5 (- 3.2; -1.8) | - 3.1 (- 3.9; - 2.4) | - 2.4 (- 3.3; - 1.4) | - 2.2 (- 3.5; - 0.9) |
| Deprivation Index* [cont.] | 64.7 (52; 77.5) | 55.9 (41.9; 70) | 264.5 (240; 288.9) | 262.1 (236.6; 287.5) |
| Metropolis (RegioStar 71)† [bin.] | 15.2 (5.5; 24.9) | 12.6 (1.5; 23.7) | - 60.3 (- 77.4; - 43.3) | - 51.3 (- 71.7; - 30.8) |
| Regiopolis and large city (RegioStaR 72)† [bin.] | 15.9 (8.3; 23.5) | 8.2 (- 0.9; 17.4) | - 38.3 (- 51.5; - 25.1) | - 30.5 (- 47.1; - 14) |
| Medium-sized city in urban region (RegioStaR 73) [bin.] | 19 (13; 25) | 10.1 (2.5; 17.6) | - | 9.9 (- 3.9; 23.7) |
| Small town or village in urban region (RegioStaR 74) [bin.] | 11 (3.9; 18.1) | 5.8 (- 2.6; 14.3) | - | 5.3 (- 10.4; 21) |
| Central city in rural region (RegioStaR 75) [bin.] | - | - 1.9 (- 12.1; 8.2) | - | 1.8 (- 17; 20.7) |
| Medium city in rural region (RegioStaR 76) [bin.] | - | - 4.1 (- 11.9; 3.7) | - | 15.2 (0.8; 29.5) |
| Small town or village in rural region (RegioStaR 77) [ref.] | - | - | - | - |
| $R^2$ [as %] (sample size) | 64.4 (4 034) | 65.1 (4 034) | 55.5 (4 204) | 55.6 (4 204) |

*Attendance calculated as follows: Assuming 8% of students in each year based on 12-year standard education. For primary schools 33% of all students, secondary schools (without Final Years) 55%, Final Years 12%. Rotating classes = 50% attendance of affected year groups. School closure emergency care (10% of Years 1-6 and special needs schools) = 5%

*Deprivation Index: RKI German Index of Social Deprivation (9,10)

†Urbanity Index by Federal Ministry of Transportation and Infrastructure; 71= metropolis, urban centre defined by infrastructure and service availability in the region; 72 = regiopolis, regional urban centre defined by infrastructure and service availability in region (11)

*Table 5: Regression Results for Ratio of Active Cases to 14-day incidences for Students or Teachers relative to the overall Population, respectively, on the two-week-lags of school-specific NPIs based on variables in Table 6*

| Variable | Student Model Estimate (p-value) | | Teacher Model Estimate (p-value) | |
|---|---|---|---|---|
| | **Final Model** | **Full Modell** | **Final Modell** | **Full Model** |
| Attendance rate in schools [as %] | - | - | - | 0 (- 0.002; 0.001) |
| Attendance rate in primary schools [as %] | - | 0 (- 0.001; 0.001) | - | - |
| Attendance rate in secondary schools [as %] | - 0.001 (- 0.002; 0) | 0 (- 0.001; 0.001) | - | - |
| Attendance rate in graduation classes [as %] | 0.001 (0.001; 0.002) | 0.001 (0; 0.002) | - | - |
| Mandatory protective face covering *for all* types of schools [bin.] | - 0.25 (- 0.28; - 0.21) | - 0.26 (- 0.3; - 0.22) | - 0.19 (- 0.25; - 0.12) | - 0.2 (- 0.3; - 0.1) |
| Mandatory protective face covering in primary school classes [bin.] | - | 0.03 [- 0.01; 0.07] | - | - 0.03 (- 0.11; 0.06) |
| Mandatory protective face covering in secondary school classes [bin.] | - | 0.23 (0.17; 0.29] | - | - 0.06 (- 0.17; 0.05) |
| Voluntary testing of employees in schools [bin.] | 0.3 (0.1; 0.4) | 0.2 (0.1; 0.3) | 0.5 (0.3; 0.7) | 0.5 (0.3; 0.7) |
| Voluntary testing of students and employees in schools [bin.] | - | - 0.02 (- 0.08; 0.04) | - 0.14 (- 0.24; - 0.04) | - 0.11 (- 0.22; - 0.01) |
| Mandatory testing of employees and secondary school students [bin.] | - | 0.3 (0.1; 0.5) | - | - 0.1 (- 0.5; 0.3) |
| Mandatory testing of employees and students for all schools [bin.] | - 0.08 (- 0.14; - 0.02) | - 0.12 (- 0.19; - 0.04) | - 0.23 (- 0.31; - 0.16) | - 0.2 (- 0.3; - 0.06) |
| Percentage of completely vaccinated population in corresponding federal state [as %] | 0.031 (0.025; 0.036) | 0.026 (0.021; 0.032) | - | - 0.002 (- 0.012; 0.008) |
| Deprivation Index* [cont.] | 0.67 (0.61; 0.74) | 0.5 (0.4; 0.6) | 1.7 (1.6; 1.8) | 1.7 (1.5; 1.9) |
| Metropolis (RegioStaR 71)† [bin.] | - | - 0.03 (- 0.1; 0.06) | - 0.3 (- 0.5; - 0.2) | - 0.3 (- 0.4; - 0.1) |
| Regiopolis and large city (RegioStaR 72)† [bin.] | - | - 0.08 (- 0.1; - 0.005) | - 0.2 (- 0.3; - 0.1) | - 0.2 (- 0.3; - 0.03) |
| Medium-sized city in urban region (RegioStaR 73) [bin.] | 0.15 [0.1; 0.19] | 0.03 (- 0.03; 0.09) | - | 0.1 (- 0.01; 0.2) |
| Small town or village in urban region (RegioStaR 74) [bin.] | 0.1 (0.05; 0.2] | 0.02 (- 0.05; 0.08) | - | 0.03 (- 0.09; 0.16) |
| Central city in rural region (RegioStaR 75) [bin.] | - | - 0.1 (- 0.2; - 0.02) | - | 0.1 (- 0.03; 0.3) |
| Medium city in rural region (RegioStaR 76) [bin.] | - | - 0.09 (- 0.15; - 0.04) | - | 0.1 (- 0.004; 0.2) |
| Small town or village in rural region (RegioStaR 77) [ref.] | - | - | - | - |
| $R^2$ [as %] (sample size) | 47.4 (4 034) | 48.7 (4 034) | 27.3 (4 204) | 27.4 (4 204) |

*Attendance calculated as follows: Assuming 8% of students in each year based on 12-year standard education. For primary schools 33% of all students, secondary schools (without Final Years) 55%, Final Years 12%. Rotating classes = 50% attendance of affected year groups. School closure emergency care (10% of Years 1-6 and special needs schools) = 5%

*Deprivation Index: RKI German Index of Social Deprivation (9,10)

†Urbanity Index by Federal Ministry of Transportation and Infrastructure; 71= metropolis, urban centre defined by infrastructure and service availability in the region; 72 = regiopolis, regional urban centre defined by infrastructure and service availability in region (11)

*Table 6: Sensitivity Analysis by background population infection trends in teachers*

| | |
|---|---|
| Category I | Class Organisation |
| k1_att | Students in attendance (%) |
| k1_gsatt | Primary school students in attendance (%) |
| k1_msatt | Secondary school students in attendance (%) |
| k1_akatt | Final Year students in attendance (%) |
| k1_gsp | Primary schools in presence classes (bin) |
| k1_gsw | Primary schools in rotating classes (bin) |
| k1_gsf | Primary schools in distance learning (bin) |
| k1_msp | Secondary schools in presence classes (bin) |
| k1_msw | Secondary schools in rotating classes (bin) |
| k1_msf | Secondary schools in distance learning (bin) |
| k1_akp | Final Years in presence classes (bin) |
| k1_akw | Final Years in rotating classes (bin) |
| k1_akf | Final Years in distance learning (bin) |
| k1_ferien | Vacation (bin) |
| Category II | Mask mandates |
| a | Surgical masks mandatory (bin) |
| b | Mask mandate applies to all school forms (including primary schools) (bin) |
| c | Masks mandated during class in primary schools (bin) |
| d | Masks mandated during class in secondary schools (bin) |
| Category III | Air quality aids |
| A | $CO_2$-monitors (bin) |
| B | Air filters (bin) |
| Category IV | Testing at schools |
| a | Tests available at schools (bin) |
| b | Voluntary testing x1/week only staff (bin) |
| c | Voluntary testing x1/week for all students and staff (bin) |
| d | Mandatory testing x1/week (x2 W16 Saarland only) only secondary schools and staff (bin) |
| e | Mandatory testing x2/week for all students and staff (bin) |
| Vacc | Percentage of completely vaccinated population in corresponding federal state (%) |
| Depr | Deprivation Index* [cont.] |
| Urb | 7-step Urbanity Index by Federal Ministry of Transportation and Infrastructure† (bin) |

Attendance calculated as follows: Assuming 8% of students in each year based on 12-year standard education. For primary schools 33% of all students, secondary schools (without Final Years) 55%, Final Years 12%. Rotating classes = 50% attendance of affected year groups. School closure emergency care (10% of Years 1-6 and special needs schools) = 5%

*Deprivation Index: RKI German Index of Social Deprivation (9,10)

†Urbanity Index by Federal Ministry of Transportation and Infrastructure (11)

*Table 7: Correlation matrix for early model variables*

| | **2WIncPop** | **k1_gsatt** | **k1_msatt** | **k1_akatt** | **k2b** | **k2c** | **k2d** | **k4b** | **k4c** | **k4d** | **k4e** | **Vacc** | **Depr** | **R71** | **R72** | **R73** | **R74** | **R75** | **R76** |
|---|---|---|---|---|---|---|---|---|---|---|---|---|---|---|---|---|---|---|---|
| **2WIncPop** | 1,00 | -0,17 | -0,32 | -0,24 | -0,09 | 0,03 | -0,20 | 0,04 | 0,28 | 0,19 | -0,32 | -0,38 | 0,07 | 0,01 | 0,04 | -0,06 | -0,05 | 0,06 | 0,10 |
| **k1_gsatt** | -0,17 | 1,00 | 0,66 | 0,60 | 0,02 | -0,07 | -0,16 | 0,10 | -0,18 | -0,01 | 0,15 | 0,24 | 0,14 | -0,04 | -0,01 | -0,04 | 0,01 | 0,04 | 0,04 |
| **k1_msatt** | -0,32 | 0,66 | 1,00 | 0,65 | -0,01 | 0,11 | 0,00 | 0,00 | -0,21 | -0,01 | 0,58 | 0,66 | 0,10 | -0,03 | 0,02 | -0,02 | 0,01 | 0,01 | 0,00 |
| **k1_akatt** | -0,24 | 0,60 | 0,65 | 1,00 | 0,08 | 0,08 | 0,02 | 0,11 | -0,19 | 0,01 | 0,41 | 0,36 | 0,07 | 0,00 | 0,05 | 0,02 | 0,00 | -0,02 | 0,03 |
| **k2b** | -0,09 | 0,02 | -0,01 | 0,08 | 1,00 | -0,32 | -0,04 | 0,18 | 0,04 | 0,11 | 0,02 | -0,03 | 0,08 | 0,09 | -0,16 | -0,04 | 0,07 | -0,01 | 0,09 |
| **k2c** | 0,03 | -0,07 | 0,11 | 0,08 | -0,32 | 1,00 | 0,33 | -0,15 | -0,01 | -0,10 | 0,16 | 0,17 | -0,33 | -0,13 | 0,17 | 0,10 | -0,12 | -0,02 | -0,12 |
| **k2d** | -0,20 | -0,16 | 0,00 | 0,02 | -0,04 | 0,33 | 1,00 | 0,06 | 0,04 | -0,33 | 0,15 | 0,08 | -0,22 | 0,07 | 0,06 | 0,07 | -0,04 | -0,07 | -0,12 |
| **k4b** | 0,04 | 0,10 | 0,00 | 0,11 | 0,18 | -0,15 | 0,06 | 1,00 | -0,09 | -0,02 | -0,16 | -0,14 | 0,22 | 0,18 | -0,02 | -0,10 | 0,01 | -0,02 | 0,02 |
| **k4c** | 0,28 | -0,18 | -0,21 | -0,19 | 0,04 | -0,01 | 0,04 | -0,09 | 1,00 | -0,06 | -0,41 | -0,19 | 0,10 | 0,04 | -0,03 | 0,01 | 0,02 | -0,01 | -0,02 |
| **k4d** | 0,19 | -0,01 | -0,01 | 0,01 | 0,11 | -0,10 | -0,33 | -0,02 | -0,06 | 1,00 | -0,10 | -0,04 | 0,11 | -0,01 | -0,02 | 0,01 | 0,00 | 0,03 | 0,03 |
| **k4e** | -0,32 | 0,15 | 0,58 | 0,41 | 0,02 | 0,16 | 0,15 | -0,16 | -0,41 | -0,10 | 1,00 | 0,78 | 0,02 | 0,02 | 0,01 | 0,01 | -0,01 | 0,00 | 0,00 |
| **Vaccinations** | -0,38 | 0,24 | 0,66 | 0,36 | -0,03 | 0,17 | 0,08 | -0,14 | -0,19 | -0,04 | 0,78 | 1,00 | 0,05 | 0,01 | 0,00 | -0,01 | -0,01 | 0,01 | 0,01 |
| **Deprivation** | 0,07 | 0,14 | 0,10 | 0,07 | 0,08 | -0,33 | -0,22 | 0,22 | 0,10 | 0,11 | 0,02 | 0,05 | 1,00 | 0,36 | 0,18 | -0,27 | -0,02 | 0,08 | -0,08 |
| **RegioStaR71** | 0,01 | -0,04 | -0,03 | 0,00 | 0,09 | -0,13 | 0,07 | 0,18 | 0,04 | -0,01 | 0,02 | 0,01 | 0,36 | 1,00 | -0,12 | -0,16 | -0,12 | -0,09 | -0,15 |
| **RegioStaR72** | 0,04 | -0,01 | 0,02 | 0,05 | -0,16 | 0,17 | 0,06 | -0,02 | -0,03 | -0,02 | 0,01 | 0,00 | 0,18 | -0,12 | 1,00 | -0,21 | -0,16 | -0,12 | -0,21 |
| **RegioStaR73** | -0,06 | -0,04 | -0,02 | 0,02 | -0,04 | 0,10 | 0,07 | -0,10 | 0,01 | 0,01 | 0,01 | -0,01 | -0,27 | -0,16 | -0,21 | 1,00 | -0,21 | -0,15 | -0,27 |
| **RegioStaR74** | -0,05 | 0,01 | 0,01 | 0,00 | 0,07 | -0,12 | -0,04 | 0,01 | 0,02 | 0,00 | -0,01 | -0,01 | -0,02 | -0,12 | -0,16 | -0,21 | 1,00 | -0,12 | -0,20 |
| **RegioStaR75** | 0,06 | 0,04 | 0,01 | -0,02 | -0,01 | -0,02 | -0,07 | -0,02 | -0,01 | 0,03 | 0,00 | 0,01 | 0,08 | -0,09 | -0,12 | -0,15 | -0,12 | 1,00 | -0,15 |
| **RegioStaR76** | 0,10 | 0,04 | 0,00 | 0,03 | 0,09 | -0,12 | -0,12 | 0,02 | -0,02 | 0,03 | 0,00 | 0,01 | -0,08 | -0,15 | -0,21 | -0,27 | -0,20 | -0,15 | 1,00 |

2WIncPop: 14-day incidence in the population (3); all others variables as in Table 6 with 2 week lag

#### S12 Text. Additional Limitations

Limitations to the data are inherent in the process of disease reporting and contact tracing. Underdetection of cases, especially in the younger age group, will affect infection risk. Regional differences in infection environment and NPI adherence affect infection risk and make inter-regional comparison difficult. NPIs affecting domains of life other than schools were not considered and differed by federal state in Germany. R1, R2 and R4 collated data on students separately, with a margin of error in picking up the relevant classification of cases from contact tracing. R3 and R5 identified students based on age, thus older (in particular vocational) students will not be perfectly reflected in the analysis. Furthermore, health agencies collate cases based on registered place of residence. In particular R4 data will be affected by cross-border (country and county) commuters and administrative differentiation of cases by residence, as would be R1, R2 and R5 through peri-urban commuting.
Contact tracing fidelity depends on capacity and capability of health agencies, reporting by index cases and is inherently flawed. This is obvious in that a) the reproduction numbers for regions where full population datasets were provided (R3, R5) is below 1 and thus not compatible with the pandemic progression witnessed and b) on average, only 65% of index cases had any contacts reported at all (R1 54%, R3 54%, R4 42% and R5 84%), leading to a skewed picture of contacts and secondary attack rates as well as biases as to which index cases would have reported contacts and how or whether they would have been identified as secondary cases by authorities in due course. Furthermore, especially for household cases the direction of transmission in instances of multiple household cases cannot always be clearly ascertained, the same holds true for attribution of origin of infected to any one contact, particularly during phases of high transmission. Among contacts, it was not possible to ascertain from the data whether these were still susceptible to infection. However, it can be assumed that biases are similar in each region as they are situated within the same state and timeline of events, and that a reduction in the proportion of the susceptible population through vaccination or infection is assumed relevant from March 2021, where it is included as a parameter in regression analysis. All considered, the SAR parameter describes further transmission after cases have been detected in the school with limitations.
Compared to health agencies, education agencies are not trained in contact tracing and systematic disease reporting, thus a greater margin of error is to be expected in those datasets gathered by education authorities.

A thorough interpretation of our results has to take into account significant limitations associated with our data. The regressions based on the KMK data span only a relatively narrow time window, which spans from calendar week 8-25 of 2021. Therefore, there may be time effects, which should be considered but cannot be included sufficiently in a statistical analysis. We tried to control for this to some extent by including the overall infection level in the local populace. Moreover, our intervention variables are obviously not ideal but rather a collection of either binary variables or approximations to include the differences in stringency of measures and attendance in school and roughly quantify the impacts these measures have on the infection risks of students and teachers as best as possible with limited data. As the NPIs have been introduced as bundles in most cases, we do not have a perfect experimental environment that would be needed for an optimal assessment of the NPIs' impacts on the infection risks. Therefore, the coefficients could be slightly biased in some cases as the Gauß-Markov assumptions, especially independence among the explanatory variables, are not completely fulfilled. We chose the variables in a manner that multicollinearity is minimised as well as possible, yet perfect independence is not given in the data. Another major limitation is the lack of comparability between the KMK and the RKI data. Whereas the RKI reports new cases weekly which can be stratified by age groups, the KMK reports known and current sick cases due to COVID-19. We tried to circumvent this limitation by comparing new cases of the previous two calendar weeks, as reported by the RKI, to the sick numbers reported by the KMK. The assumption behind this is that students or teachers currently excused from school due to an infection have been infected in one of the previous two weeks, which appears a reasonable assumption given the incubation time (13). Furthermore, KMK data do not discriminate by type of school on the district level and demographic data on the students and teachers is not reported. Finally, the KMK data may be vulnerable towards higher underdetection of COVID-19 cases, as students and teachers are not obligated to report their reason of absence correctly, i.e. it is not ensured that someone being excused from school will indeed report the real reason for their absence.

#### S13 Text. Supplement References

1. Statistisches Bundesamt, Deutsches Zentrum für Hochschul- und Wissenschaftsforschung. (2020). GENESIS-Online Datenbank; Fachserie 11 Reihe 1, Fachserie 11 Reihe 2, Fachserie 11 Reihe 4.1. Available at https://www.destatis.de/DE/Service/Bibliothek/_publikationen-fachserienliste-11.html
Last accessed 15.11.21

11. Bundesministerium für Verkehr und Digitale Infrastruktur (BMVDI). Regionalstatistische Raumtypologie. (2020). Available at
https://www.bmvi.de/SharedDocs/DE/Artikel/G/regionalstatistische-raumtypologie.html
Last accessed 18.10.2021

12. Statistisches Bundesamt (2021). Ergebnisse der Bevölkerungsfortschreibung auf Grundlage des Zensus 2011. https://www.destatis.de/DE/Themen/Gesellschaft-Umwelt/Bevoelkerung/Bevoelkerungsstand/Tabellen/bevoelkerung-nichtdeutsch-laender.html Last accessed 18.11.2021

13. McAloon C, Collins Á, Hunt K, Barber A, Byrne AW, Butler F, et al. Incubation period of COVID-19: a rapid systematic review and meta-analysis of observational research. BMJ Open. 2020;10(8):e039652

**Sources for Infection Control Measures**

School-specific public health interventions were sourced from federal and local state and educational authority websites on multiple dates between 23 February and 11 August 2021 (infection control policies; laws; decrees; frameworks for hygiene measures; information to schools, parents and students; press releases; other forms of publicly available documents). Source websites and relevant overview access points are given below ordered by federal state.

Baden-Württemberg
Ministerium für Kultus, Jugend und Sport Baden-Württemberg. (2021).
https://km-bw.de
https://km-bw.de/,Lde/startseite/sonderseiten/corona
https://km-bw.de/,Lde/startseite/sonderseiten/verordnungen-corona
https://km-bw.de/,Lde/startseite/sonderseiten/corona-verordnung-schule
https://km-bw.de/,Lde/startseite/sonderseiten/faq-corona-schule
https://km-bw.de/,Lde/startseite/service/Pressemitteilungen
https://km-bw.de/,Lde/startseite/service/Archiv+Pressemitteilungen+2020

Bavaria
Bayerisches Staatsministerium für Unterricht und Kultus. (2021).
https://www.km.bayern.de/
https://www.km.bayern.de/eltern/meldung/7061/aktualisierter-rahmen-hygieneplan-fuer-bayerische-schulen.html
https://www.km.bayern.de/allgemein/meldung/7047/faq-zum-unterrichtsbetrieb-an-bayerns-schulen.html
https://www.km.bayern.de/pressemitteilungen.html

Berlin
Senatsverwaltung für Bildung, Jugend und Familie Berlin. (2021)
https://www.berlin.de/sen/bjf/
https://www.berlin.de/sen/bjf/corona/
https://www.berlin.de/sen/bjf/corona/schule/
https://www.berlin.de/sen/bjf/corona/briefe-an-schulen/
https://www.berlin.de/sen/bjf/service/presse/pressearchiv-2021/
https://www.berlin.de/sen/bjf/service/presse/pressearchiv-2020/

Brandenburg
Ministerium für Bildung, Jugend und Sport Brandenburg. (2021)
https://mbjs.brandenburg.de/corona-aktuell.html

https://mbjs.brandenburg.de/corona-aktuell/schule-und-unterricht.html
https://mbjs.brandenburg.de/corona-aktuell/chronologie.html
https://mbjs.brandenburg.de/aktuelles/pressemitteilungen.html

Bremen
Die Senatorin für Kinder und Bildung Bremen. (2021)
https://www.bildung.bremen.de/start-251714
https://www.senatspressestelle.bremen.de/ressorts/die-senatorin-fuer-kinder-und-bildung-1612?template=20_pmressort_d
https://www.bildung.bremen.de/detail.php?gsid=bremen117.c.340812.de
https://www.bildung.bremen.de/detail.php?gsid=bremen117.c.340822.de
https://www.bildung.bremen.de/detail.php?gsid=bremen117.c.340818.de
https://www.bildung.bremen.de/detail.php?gsid=bremen117.c.340816.de

Hamburg
Behörde für Schule und Berufsbildung Hamburg. (2021)
https://www.hamburg.de/bsb/
https://www.hamburg.de/bsb/faq/
https://www.hamburg.de/bsb/pressemitteilungen/
https://www.hamburg.de/bsb/publikationen-a-z/

Hesse
Hessisches Kultusministerium. (2021)
https://kultusministerium.hessen.de/
https://kultusministerium.hessen.de/Schulsystem/Corona
https://kultusministerium.hessen.de/Schulsystem/Corona/FAQ-Corona
https://kultusministerium.hessen.de/Schulsystem/Corona/Dokumente-zur-Unterrichtsorganisation
https://kultusministerium.hessen.de/Presse

Mecklenburg- Western Pommerania
Ministerium für Bildung, Wissenschaft und Kultur Mecklenburg-Vorpommern. (2021)
https://www.regierung-mv.de/Landesregierung/bm/
https://www.regierung-mv.de/Landesregierung/bm/Blickpunkte/Coronavirus/Coronavirus-%E2%80%93-Informationen-f%C3%BCr-schule/Zu-den-Regelungen-f%C3%BCr-die-Schulorganisation/
https://www.regierung-mv.de/Landesregierung/bm/Presse/Aktuelle-Pressemitteilungen/

Lower Saxony
Niedersächsisches Kultusministerium. (2021)
https://www.mk.niedersachsen.de/startseite/
https://www.mk.niedersachsen.de/startseite/aktuelles/
https://www.mk.niedersachsen.de/startseite/aktuelles/schule-neues-schuljahr-190409.html
https://www.mk.niedersachsen.de/startseite/aktuelles/aktuelle_erlasse_zum_schuljahr_2021_22/corona-erlasse-zum-schuljahr-2021-22-202659.html

https://www.mk.niedersachsen.de/startseite/aktuelles/schule_in_corona_zeiten/faq-schule-196173.html
https://www.mk.niedersachsen.de/startseite/aktuelles/presseinformationen/

North Rhine-Westphalia
Ministerium für Schule und Bildung des Landes Nordrhein-Westfalen. (2021)
https://www.schulministerium.nrw/
https://www.schulministerium.nrw/angepasster-schulbetrieb-corona-zeiten
https://www.schulministerium.nrw/presse/pressemitteilungen

Rhineland- Palatinate
Ministerium für Bildung Rheinland-Pfalz. (2021)
https://corona.rlp.de/de/themen/schulen-kitas/schule/
https://corona.rlp.de/de/themen/schulen-kitas/schule/dokumente-schule/
https://corona.rlp.de/de/themen/schulen-kitas/schule/faqs-schule/
https://corona.rlp.de/de/themen/schulen-kitas/schule/faqs-schule/schulartuebergreifende-hinweise/
https://bm.rlp.de/de/service/pressemitteilungen/

Saarland
Ministerium für Bildung und Kultur Saarland. (2021)
https://www.saarland.de/mbk/DE/home/home_node.html
https://www.saarland.de/mbk/DE/aktuelles/aktuelle-meldungen/aktuelle-meldungen_node.html
https://www.saarland.de/DE/portale/corona/bildung-kultur/bildung/aktuelles-bildung/aktuelles-bildung_node.html
https://www.saarland.de/DE/portale/corona/faq/bildung-kultur/schulen-kitas/schulen-kitas_node.html
https://www.saarland.de/DE/portale/corona/bildung-kultur/bildung/rundschreiben-downloads/rundschreiben-downloads_node.html

Saxony
Sächsischen Staatsministeriums für Soziales und Gesellschaftlichen Zusammenhalt. (2021).
https://www.coronavirus.sachsen.de/faq-infektionsschutz-6050.html
https://www.coronavirus.sachsen.de/amtliche-bekanntmachungen.html?_cp=%7B%7D
https://www.coronavirus.sachsen.de/eltern-lehrkraefte-erzieher-schueler-4144.html?_cp=%7B%22accordion-content-11200%22%3A%7B%220%22%3Atrue%7D%2C%22previousOpen%22%3A%7B%22group%22%3A%22accordion-content-11200%22%2C%22idx%22%3A0%7D%7D
https://www.bildung.sachsen.de/blog/

Saxony-Anhalt
Ministerium für Bildung Sachsen-Anhalt. (2021)
https://mb.sachsen-anhalt.de/
https://mb.sachsen-anhalt.de/themen/schule-und-unterricht/schulbetrieb-in-der-corona-pandemie/

https://mb.sachsen-anhalt.de/presse/pressemitteilungen/archiv-pressemitteilungen/?no_cache=1

Schleswig-Holstein
Ministerium für Bildung, Wissenschaft und Kultur Schleswig-Holstein. (2021)
https://www.schleswig-holstein.de/DE/Landesregierung/III/iii_node.html
https://www.schleswig-holstein.de/DE/Schwerpunkte/Coronavirus/Schulen_Hochschulen/corona_schule.html
https://www.schleswig-holstein.de/DE/Fachinhalte/S/schulrecht/Glossareintraege/C/corona.html
https://www.schleswig-holstein.de/DE/Landesregierung/III/Presse/Pressemitteilungen/pressemitteilungen_node.html

Thuringia
Ministerium für Bildung, Jugend und Sport Thüringen. (2021)
https://bildung.thueringen.de/
https://bildung.thueringen.de/ministerium/coronavirus
https://bildung.thueringen.de/ministerium/coronavirus/schule
https://bildung.thueringen.de/ministerium/medienservice/nachrichtenarchiv
